# “The effect of dietary fiber based on fermentability and viscosity on the gut microbial metabolites in chronic kidney disease: a systematic review and meta-analysis of experimental and clinical trials.”

**DOI:** 10.64898/2026.07.09.26357677

**Authors:** Seyedeh Nooshan Mirmohammadali, Claudia Carrillo, Jason B. Reed, Brandon M. Kistler, Hannah E Wilson, Bruce Hamaker, Sharon Moe, Annabel Biruete

**Author notes:** Corresponding author: Annabel Biruete, PhD, RD, LD, 700 Mitch Daniels Blvd., Stone Hall Room 207 West Lafayette, IN 47907.

## Abstract

**Background:** Chronic kidney disease (CKD) is associated with alterations in the gut microbiome that promote the accumulation of gut-derived uremic solutes and contribute to systemic inflammation, vascular dysfunction, and disease progression. Dietary fiber has emerged as a promising modulator of gut microbial metabolism, yet the influence of fiber physicochemical properties, particularly fermentability and viscosity, on uremic metabolite production in CKD remains poorly understood.

**Objective:** To systematically evaluate the effects of isolated dietary fiber interventions, classified by fermentability and viscosity, on gut microbial metabolites in CKD across experimental rodent models and randomized clinical trials, and to determine whether these fiber properties modify microbial metabolites.

**Methods:** A systematic search of PubMed, Embase, CINAHL, and Cochrane Library (through June 2026) identified randomized controlled trials and controlled rodent studies assessing isolated dietary fiber in CKD. Eligible studies reported at least one gut-derived metabolite (i.e., indoxyl sulfate (IS), p-cresyl sulfate (PCS), trimethylamine-N-oxide (TMAO), tryptophan-derived indoles, or short-chain fatty acids (SCFAs)). Random-effects models were used for pooled estimates using weighted mean differences (WMD) for human studies and standardized mean differences (SMD) for animal studies. Subgroup analyses evaluated fiber fermentability, viscosity, intervention dose, duration, and CKD stage. Risk of bias was assessed with ROB-2 and SYRCLE, and evidence certainty with GRADE.

**Results:** Twenty-eight studies (13 human, 15 animal) met eligibility criteria, comprising 511 participants and 312 animals with CKD. Isolated fiber supplementation, primarily fermentable and non-viscous fibers, reduced IS (human: −0.13 mg/dL; 95% CI: −0.25, −0.01; p = 0.03; animal: −1.99; 95% CI: −3.06, −0.92; p < 0.0001) and pCS (human: −0.23 mg/dL; 95% CI: −0.46, 0.001; p = 0.051; animal: −1.56; 95% CI: −2.08, −1.03; p < 0.0001). SCFAs increased in animal studies, including cecal acetate (2.00, 95% CI: 0.78 to 3.22; p = 0.001) and circulating propionate (1.51, 95% CI: 0.054 to 2.96; p=0.04). There were no dose-dependent effects, but longer interventions (>8 weeks) tended to lower pCS (−0.26 mg/dL, 95% CI: −0.55 to 0.02; p=0.06). Some heterogeneity and low-to-moderate certainty were observed.

**Conclusion:** Isolated dietary fiber reduces major gut-derived uremic solutes in CKD, with fermentability influencing metabolic responsiveness, but with minimal studies on viscous fibers. Larger, longer-duration trials with standardized reporting of total fiber intake and clinical endpoints are needed to guide evidence-based dietary recommendations in CKD.

**Statement of Significance:** This is the first systematic review and meta-analysis examining the effects of isolated dietary fiber on gut-derived metabolites comprising human and rodent models in CKD.

This trial was registered at PROSPERO as CRD42023483468.

## Introduction

Chronic kidney disease (CKD) is a progressive condition defined by a sustained reduction in glomerular filtration rate (GFR < 60 mL/min/1.73 m²) or proteinuria for at least 3 months (1). Declining kidney function leads to impaired excretion of metabolic waste products, disturbances in mineral and electrolyte homeostasis, abnormalities in acid base balance, and defects in bone and mineral metabolism due to reduced vitamin D activation (2). These alterations contribute to metabolic acidosis, chronic kidney disease-mineral and bone disorder (CKD-MBD), and the accumulation of uremic solutes, all of which reinforce a persistent proinflammatory and oxidative milieu (3). As CKD progresses, these interconnected disturbances extend beyond the kidneys to impact multiple organ systems, including the cardiovascular, endocrine, and gastrointestinal systems, with marked effects on the gut microbiome (4). Among the nontraditional complications of CKD, perturbation of the gut microbiome has become increasingly recognized (5). CKD associated alterations in the colonic environment, intestinal transit, luminal pH, and dietary patterns shift the gut microbial community toward a more proteolytic and less saccharolytic profile (5,6). This altered microbial state promotes enhanced production and systemic accumulation of gut-derived uremic toxins such as indoxyl sulfate (IS), p-cresyl sulfate (pCS), and trimethylamine-N-oxide (TMAO), which contribute to inflammation and oxidative stress, leading to endothelial dysfunction and the promotion and progression of cardiovascular disease and CKD (7). Ultimately, these gut-derived uremic toxins have been associated with higher mortality in CKD, making them a target for intervention to improve outcomes in people with CKD (8–10).

Gut microbial metabolism produces a wide spectrum of bioactive compounds through distinct biochemical pathways, many of which become clinically relevant in CKD. Tryptophan metabolism via bacterial tryptophanases generates indole, which is subsequently absorbed and hepatically converted through sulfation or oxidation pathways to produce uremic solutes such as IS (11). Tyrosine and phenylalanine fermentation through bacterial dehydroxylation and decarboxylation pathways yield p-cresol, which undergoes hepatic conjugation to form pCS and p-cresyl glucuronide (pCG), respectively (12). Another clinically relevant metabolite is trimethylamine (TMA), which can be obtained directly from fish or produced by the gut microbiota from dietary choline and carnitine. This TMA is subsequently oxidized in the liver by flavin-containing monooxygenase 3 (FMO3) to form TMAO (13). Together, these metabolites accumulate disproportionately in CKD due to impaired renal clearance, amplifying inflammation, oxidative stress, endothelial injury, and uremia (7).

In contrast, saccharolytic fermentation of dietary fibers produces short-chain fatty acids (SCFAs), including acetate, propionate, and butyrate, which strengthen epithelial barrier function, regulate immune signaling, modulate blood pressure, and exert anti-inflammatory effects (14,15). The perturbed balance between these saccharolytic and proteolytic metabolic products shaped by diet, gut microbial composition, and host physiology in CKD contribute to vascular dysfunction, oxidative stress, inflammation, and accelerated cardiovascular and CKD progression (7). Therefore, the disproportionate accumulation of gut-derived uremic toxins and the decline in SCFAs highlight the importance of identifying modifiable dietary strategies that can shift gut microbial metabolism toward more favorable pathways, most notably through the use of dietary fiber.

Dietary fiber is broadly defined as nondigestible carbohydrates and associated compounds that escape digestion in the small intestine and undergo partial or complete fermentation in the colon (16). Historically, dietary fibers have been categorized as soluble or insoluble based on their ability or inability to dissolve in water. Some soluble fibers form viscous solutions. Solubility has also often been used to infer fermentability, with soluble fibers generally considered fermentable by the gut microbiota and insoluble fibers considered less fermentable. However, this nomenclature provides limited insight into physiological function, microbial metabolism, or clinical outcomes, as some soluble fibers are not fermentable and many insoluble fibers can undergo partial or full fermentation. Moreover, solubility does not accurately predict effects on luminal viscosity, gastrointestinal transit, substrate availability for the gut microbiota, or the anatomical site and rate of fermentation (17). In contrast, classification based on fermentability and viscosity offers a more physiologically relevant framework for understanding fiber-microbiota interactions. Fermentability reflects the capacity of a fiber to be metabolized by the gut microbiota to generate SCFAs (18), whereas viscosity reflects its ability to form pastes and gels and influence luminal transit, nutrient absorption, and intraluminal mixing (19). These physicochemical properties can determine not only which microbial taxa can utilize the substrate but also the kinetics and anatomical location of fermentation (17). Fermentable fibers such as inulin, resistant starches, and select oligosaccharides promote saccharolytic metabolism, SCFA production, and lowering of colonic pH, whereas minimally fermentable fibers (e.g., cellulose, psyllium husk) primarily exert bulking effects, increase viscosity, or modify intestinal transit (20). A growing body of literature suggests that these fiber-dependent actions may reduce the generation or systemic appearance of uremic toxins by enhancing saccharolytic fermentation, suppressing protein fermentation, fasten transit time, improving epithelial barrier function, and modulating microbial composition and metabolic activity (21). Importantly, these distinctions may be particularly relevant in CKD, where the gut microbial ecosystem is characterized by reduced saccharolytic capacity, increased proteolytic metabolism, impaired barrier integrity, and enhanced production of gut-derived uremic solutes (22). Thus, identifying which fiber types such fermentable, non- or minimally fermentable, viscous, or nonviscous, most effectively modulate uremic metabolites is essential for guiding the development of targeted, mechanism-based nutrition strategies for CKD.

Despite increasing interest, the evidence base remains fragmented. Clinical trials vary widely in fiber type, dose, duration, participant characteristics, and outcome measures, while animal studies differ in CKD models, feeding regimens, microbiome methods, and metabolite quantification. Prior reviews have not integrated findings across fiber classes defined by fermentability and viscosity, nor have they synthesized results from both experimental and clinical interventions. Consequently, key mechanistic uncertainties persist, including which fiber types most effectively lower gut-derived uremic toxins, whether fermentable fibers consistently reduce metabolites such as IS, pCS, or TMAO across CKD stages and models, and the extent to which viscosity contributes independently of fermentability. Therefore, the aim of this systematic review and meta-analysis is to comprehensively evaluate the effects of dietary fiber interventions, classified by fermentability and viscosity, on gut microbial metabolites in CKD across both experimental animal models and human clinical trials. This work seeks to identify fiber-specific effects on uremic toxin concentrations and assess whether fiber physicochemical properties modify treatment response. By integrating evidence across study designs, fiber types, and CKD stages, this review provides a physiologically grounded framework to guide targeted fiber interventions for CKD dietary management and future clinical trial design.

## Methods

This systematic review and meta-analysis was conducted according to the methodological standards described in the Cochrane Handbook for Systematic Reviews of Interventions for human studies and SYRCLE’s guide for systematic reviews of preclinical animal intervention studies (23). The review was reported in accordance with the Preferred Reporting Items for Systematic Reviews and Meta-Analyses (PRISMA) 2020 guidelines (24). The review protocol was prospectively registered with the International Prospective Register of Systematic Reviews (PROSPERO) (No. CRD42023483468). The study was designed based on the PICOS (Population, Intervention, Comparison, Outcomes, and Study design) framework (25), described in **Table 1**.

**Table 1:** PICOS framework used to define eligibility criteria for the systematic review and meta-analysis. PICOS refers to Population, Intervention, Comparison, Outcomes, and Study design. Chronic kidney disease was defined according to study-specific criteria. Dietary fiber interventions were categorized based on fermentability and viscosity. Gut-derived microbial metabolites included uremic solutes and short-chain fatty acids, when reported.

| Parameter | Description |
| --- | --- |
| Population (P) | Adults with chronic kidney disease and experimental animal models of chronic kidney disease |
| Intervention (I) | Dietary fiber supplementation, classified according to fermentability and viscosity |
| Comparison (C) | No dietary fiber intervention or standard/control diet without added fiber |
| Outcomes (O) | Changes in gut-derived microbial metabolites, including uremic solutes (indoxyl sulfate, *p*-cresyl sulfate, trimethylamine *N*-oxide) and short-chain fatty acids, when reported |
| Study Design (S) | Randomized controlled trials and experimental animal studies |

### Eligibility criteria

Studies were eligible for inclusion if they were original randomized controlled trials or experimental animal studies evaluating the isolated effects of dietary fiber interventions in the context of CKD. Eligible populations included adults aged 18 years or older with CKD at any stage, either pre-dialysis or receiving any form of dialysis, as well as established rodent models of CKD. Animal models included Cy/+ rats, adenine-induced CKD models, subtotal, partial, or 5/6 nephrectomy models, and *Col4A3* knockout models. In addition, CKD-relevant comorbid animal models, including type 1 diabetes induced by streptozotocin, hypertension models such as the Dahl salt-sensitive rat, and homozygous mutation on leptin receptor gene model, were eligible. Studies conducted in healthy humans or animals, children or adolescents, or models of acute kidney injury were excluded. Eligible interventions consisted of dietary fiber provided as a single, clearly defined supplement, allowing characterization of fiber fermentability and/or viscosity. Fiber was required to be administered as an isolated supplement rather than incorporated within whole foods, fruits, vegetables, or mixed dietary patterns. Studies using multi-component interventions were excluded when the independent effects of dietary fiber could not be clearly discerned. No restrictions were applied based on geographic region; however, the search was limited to articles published in English.

Eligible studies were required to include a comparator arm, consisting of a standard diet, control diet, or placebo, and studies lacking a comparator were excluded. To be eligible, studies were also required to report at least one outcome related to gut-derived microbial metabolites such as IS, p-CS, or TMAO; other indole derivatives including IAA and IPA; and SCFAs, including acetate, propionate, and butyrate. Outcomes could be measured in urine, serum, plasma, feces, or cecal contents. Only studies reporting sufficient quantitative data, such as means and measures of variability, to allow calculation of effect estimates were included. *In vitro* studies, observational studies, case–control studies, case reports or case series, and review articles were excluded (See **Table 2**).

**Table 2.** Eligibility criteria were defined a priori using the PICOS framework (Population, Intervention, Comparator, Outcomes, and Study design).

| Domain | Inclusion Criteria | Exclusion Criteria |
| --- | --- | --- |
| **Population (P)** | Adults ($\geq 18$ years) with chronic kidney disease (CKD) at any stage, including pre-dialysis or dialysis populations; experimental animal models of CKD, including Cy/+ rats, adenine-induced CKD models, subtotal/partial/5/6 nephrectomy models, and *Col4A3* knockout models; CKD-relevant comorbid animal models including type 1 diabetes (streptozotocin-induced) and hypertension models such as the Dahl salt-sensitive rat | Healthy humans or animals; children or adolescents; models of acute kidney injury |
| **Intervention (I)** | Dietary fiber provided as a single, clearly defined supplement allowing characterization of fermentability and/or viscosity; purified fiber sources (e.g., wheat bran); prebiotic or synbiotic formulations; minimum intervention duration of $\geq 1$ week | Fiber provided primarily through whole foods, fruits, vegetables, cereals, or mixed dietary patterns; multi-component interventions in which the independent effect of dietary fiber could not be discerned |
| **Comparator (C)** | Standard diet, control diet, or placebo comparator | Studies without a comparator group |
| **Outcomes (O)** | Gut-derived microbial metabolites, including uremic solutes (indoxyl sulfate, p-cresyl sulfate, trimethylamine N-oxide), indole derivatives (indole-3-acetic acid, indole-3-propionic acid), and short-chain fatty acids (acetate, propionate, butyrate), measured in urine, serum, plasma, feces, cecal contents, or intestinal samples | Studies reporting only clinical outcomes without gut microbiota or microbial metabolite data |
| **Study Design (S)** | Original randomized controlled trials and experimental animal studies with sufficient quantitative data (e.g., means and measures of variability) to calculate effect estimates | *In vitro* studies; observational studies; case-control studies; case reports or case series; review articles |

### Literature search

A comprehensive literature search was conducted by a research librarian (JBR) and a PhD candidate (SNM) on 15 November 2023 and was updated on 12 June 2026, which resulted in the addition of one RCT and two animal studies. The search was performed across four electronic databases: PubMed, Embase, CINAHL, and the Cochrane Library, including CENTRAL and Cochrane Reviews. The search strategy incorporated controlled vocabulary and keyword terms related to CKD, dietary fiber, gut microbiota, fermentable and nonfermentable fibers, and gut-derived microbial metabolites, including uremic solutes and short-chain fatty acids. Reference lists of included articles and relevant reviews were manually screened to identify additional potentially eligible studies. Citation searching was also conducted to ensure comprehensive coverage of the literature. The full electronic search strategy for all databases is provided in **Supplementary Document**.

### Study selection and data extraction

Search results from each database were imported into Covidence reference management software (26), where duplicate records were removed, and titles and abstracts as well as full texts were screened independently and in duplicate by two reviewers (AB, SNM), with disagreements resolved by discussion or, when necessary, through consultation with a senior reviewer (BK). Data from all eligible studies were collected using a standardized data extraction template developed in Microsoft Excel and completed independently by three reviewers (SNM, CC, and HEW). The search strategy was deliberately broad to capture studies examining both gut microbiota composition and gut-derived microbial metabolites in the context of CKD and dietary fiber interventions. Given the breadth of the search results, two separate systematic reviews were planned: one focusing on gut microbiota composition and the other on microbial metabolites. Accordingly, studies included in the present analysis were manually restricted to those in which gut-derived microbial metabolites were evaluated. Extracted information included key study characteristics such as study identifier, country, study design, population, sex distribution, participant health status, trial duration (weeks), intervention and control sample sizes, mean age of participants in each group, intervention ingredient, fiber type and dosage (g/day), control ingredient and dosage (g/day), and the method used for metabolite assessment. For animal studies, additional details including the animal model and the method used to induce CKD were recorded. When data were missing or unclear, corresponding authors were contacted via email to request further clarification or additional information (**Tables 3a-b**). Studies lacking essential data and for which authors did not respond to inquiries were excluded.

### Subgroups of interest

Subgroup analyses were conducted based on (1) fiber type (a) fructans, including inulin, xylooligosaccharides, oligofructose enriched inulin, galacto oligosaccharides, fructooligosaccharides, and arabinoxylan oligosaccharides; b) gums, including gum acacia, unmodified guar gum, and partially hydrolyzed guar gum; c) hemicellulose, represented by psyllium seed husk; d) resistant starch, represented by resistant starch type 2 (Hi-Maize 260); and e) beta glucan), (2) viscosity (viscous or non-viscous), (3) fermentability (fermentable or non-fermentable), (4) trial dose (≥16 g/d versus <16 g/d for RCTs and ≥10 versus <10 dose (% (w/w)) for preclinical studies), (5) intervention duration (≤8 versus >8 weeks for RCTs and ≤4 versus >4 weeks for preclinical studies) for human and animal studies, and (6) dialysis status (dialysis versus non dialysis) just for human trials. Subgroup analyses were performed only when at least two studies contributed data to the outcome of interest for each subgroup and when the overall meta-analytic effect was statistically significant.

### Quality assessment

The methodological quality of all included human RCTs and preclinical animal studies was independently assessed by three reviewers (SNM, CC, HEW). RCTs were evaluated using the Cochrane Risk of Bias 2 (RoB 2) framework (27) and animal intervention studies were assessed using the SYRCLE risk of bias tool (23) with any disagreements resolved through consultation with a fourth reviewer (AB). RoB 2 evaluates five domains: randomization process, deviations from intended interventions, missing outcome data, outcome measurement, and selection of reported results, with each domain rated as low risk, some concerns, or high risk. The SYRCLE tool assesses ten domains, including sequence generation, baseline characteristics, allocation concealment, random housing, caregiver and outcome assessor blinding, random outcome assessment, completeness of outcome data, selective reporting, and other potential sources of bias, with judgments categorized as yes, no, or unclear marked as low, high, or unclear risk of bias, respectively. Unlike the Cochrane RoB2 tool for RCTs, which generates an overall risk-of-bias judgment, the SYRCLE tool advises against calculating a summary score for preclinical studies. This approach avoids imposing arbitrary weighting across bias domains, since the relative importance of issues such as blinding or allocation concealment varies by outcome and study design. A single aggregated score may also obscure critical methodological weaknesses by combining high-and low-risk elements into one value, thereby limiting differentiation between robust studies and those with insufficient reporting. For these reasons, SYRCLE recommends evaluating each domain independently to maintain transparency and preserve the interpretability of risk-of-bias assessments, and that is why the overall column for animal studies is blank (23). For both animal and human studies, robvis visualization tool was utilized (28).

### Certainty Assessment

The certainty of evidence for each outcome was assessed using the Grading of Recommendations Assessment, Development, and Evaluation (GRADE) framework (29). This approach considers five domains: risk of bias, indirectness, imprecision, inconsistency, and publication bias. Based on these criteria, the certainty of evidence was rated as high, moderate, low, or very low. Assessments were performed independently by two reviewers (SNM and CC), with discrepancies resolved through discussion.

### Statistical analyses

Statistical analyses were conducted using Stata SE version 19.0 (StataCorp, College Station, TX, USA). Statistical significance was defined as a two-sided p-value < 0.05. The statistical analyses for the human and animal studies were conducted separately, reflecting the distinct nature of their study designs, data collection methods, and reporting practices. RCTs were analyzed as follows: Summary estimates were generated as pooled weighted mean differences (WMDs) using random-effects models to account for between-study variability (30). Standardized mean differences (SMDs) were explored initially; however, weighted mean differences were used for the primary analyses because outcomes were measured using uniform units and comparable assays across studies. WMDs retain the original scale of measurement and enhance clinical interpretability. Mean differences were calculated as changes from baseline to post-intervention, and when unavailable, the standard deviation of the change was estimated using a standard correlation-based formula (SD = √[(SD at baseline)² + (SD at the end)² − (2 × r × SD at baseline × SD at the end)] (31), with a correlation coefficient (r) of 0.8. In studies where standard deviations were unavailable, reported standard errors, 95% confidence intervals, or interquartile ranges were converted to standard deviations according to the method of Hozo et al. (32). As long as a study reported the sample size, median, and interquartile range, the corresponding mean and standard deviation were estimated using established statistical conversion methods (33–35). Meta-analysis was performed for outcomes reported by at least two independent studies. Subgroup analyses were conducted to explore potential sources of heterogeneity and to examine whether fiber type, fiber characteristics, dosage, intervention duration, or dialysis status modified the effect. Subgroups were analyzed only when at least two studies contributed data to that subgroup. Heterogeneity was evaluated using Cochrane’s Q test with a significance level of p < 0.10 and the I² statistic, defined as low (0–40%), moderate (30–60%), substantial (50–90%), and considerable (75–100%) (36). Sensitivity analyses were carried out using a leave-one-out approach (37,38). Publication bias was assessed through visual inspection of funnel plots as well as Egger’s statistical tests. Meta-regression was performed for outcomes that included ≥9 studies, to assess the presence of a linear association between dietary fiber exposure and the corresponding metabolite concentration. A dose–response meta-analysis was conducted for outcomes in which three or more studies provided estimates across at least two levels of dietary fiber exposure. This minimum was required to ensure adequate variation in dose and to enable estimation of a linear dose-response slope (39). Animal studies were analyzed in accordance with the guidelines described by Ineichen et al. (40) as follows, summary estimates were generated as SMDs, as the inclusion of both rat and mouse studies requires a common scale to ensure consistent interpretation of changes in each outcome across experiments, and random effects models were applied because heterogeneity is explicitly modelled and accounted for, making this approach the preferred method for preclinical meta-analyses. Data from preclinical studies have been synthesized using this framework to provide estimates that reflect both within study and between study variation. Mean differences were calculated using the change between the final values of the control and intervention groups because baseline measurements are not collected for metabolite outcomes in animal studies. The SD and all other calculations were performed in the same manner as described for the human studies above. All the codes for the analysis performed in Stata SE version 19.0 is provided in supplementary files.

## Results

Through database searches, 8,517 records were identified, all of which were imported for screening, in addition to one study found through citation searching. After removal of 1,666 duplicate records, including five identified manually and 1,661 identified using Covidence, 6,852 unique records remained for title and abstract screening. Of these, 6,732 records were excluded as not relevant to the research question. The remaining 120 articles were retrieved and assessed for full-text eligibility. Following full-text review, 76 studies were excluded for the following reasons: abstract-only publications (n = 35), wrong intervention (n = 28), trial registration records (n = 4), wrong study design (n = 3), wrong patient population (n = 3), protocol papers (n = 1), incomplete data report (n = 2) and wrong outcomes (n = 1). Ultimately, 28 studies (13 human studies (41–53) and 15 animal studies (54–68) met the inclusion criteria and were included in the final systematic review (**Figure 1**).

**Figure 1.**
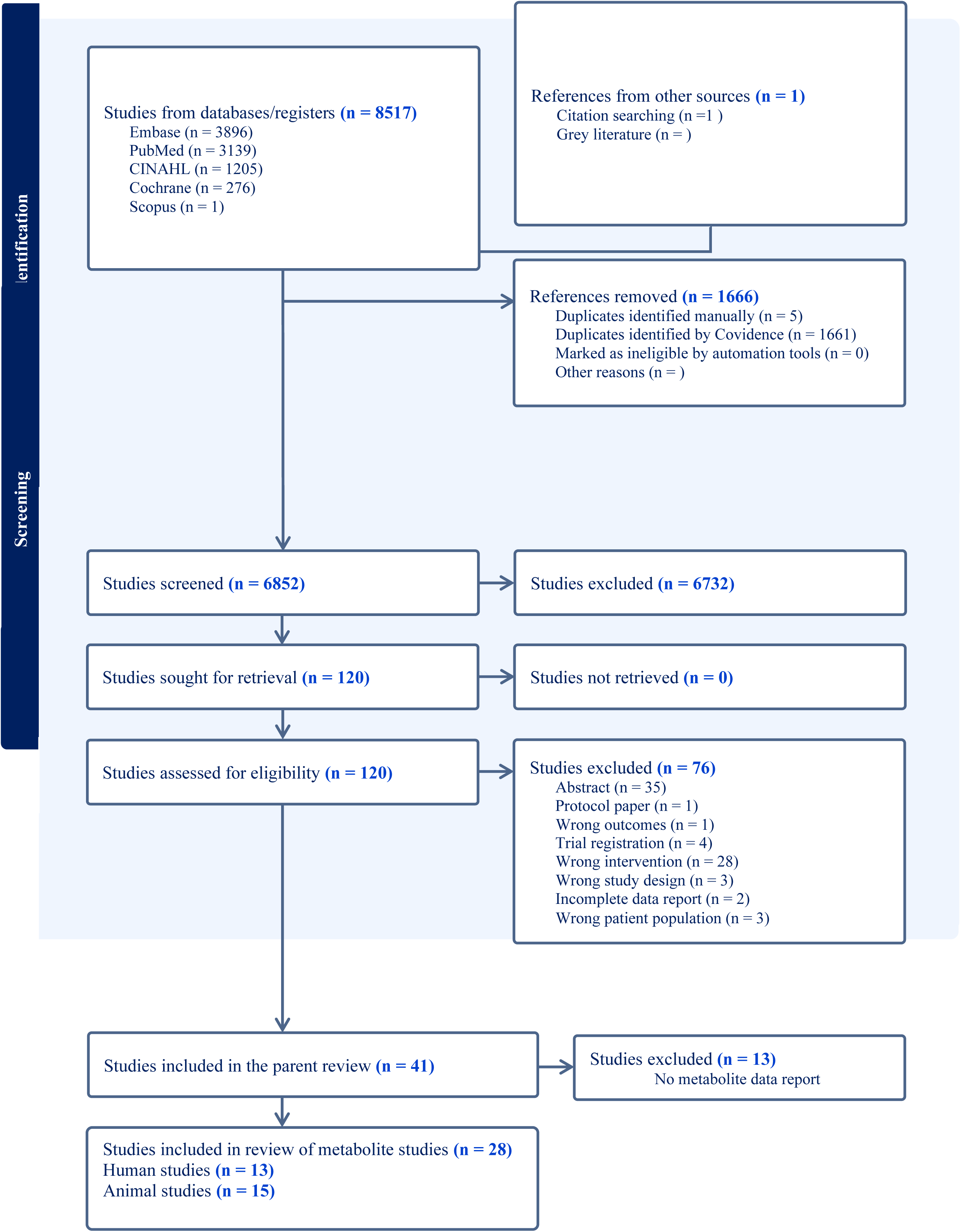
PRISMA flow chart of the study selection process for inclusion studies in the systematic review. 2a)

### Basic characteristics of human studies

The meta-analysis included 14 randomized controlled trials (RCTs) (41–53) encompassing a total of 511 participants, with 256 assigned to intervention groups and 255 to control groups; some participants contributed to both groups due to crossover study designs. Most trials were conducted in Brazil (n = 6) (41,43,47,48,53,69) and the United States (n = 3) (44,45,52) with additional studies carried out in China (n = 2) (46,49), Belgium (n = 1) (50), and South Africa (n = 1) (51). All included studies were randomized and placebo-controlled, employing either parallel or crossover designs. Study populations consisted of individuals with CKD, some undergoing dialysis (42–48,52). Trial durations varied from 4 to 16 weeks, with the majority lasting between 4 and 8 weeks. Mean participant ages ranged from 18 to 77 years, and all studies included both male and female participants. Interventions involved supplementation with various dietary fibers, including inulin, resistant starch type 2, β-glucan, and arabinoxylan oligosaccharides, administered at doses ranging from 3 to 21 g/day. Control interventions included maltodextrin, waxy corn starch (Amioca), and wheat starch combined with a low-protein diet, with doses ranging from 10 to 16 g/day. Two studies did not include a placebo control (44,51) (See **Table 3a**).

**Table 3a.** Characteristic of included human studies in the meta-analysis Abbreviations: DB, double-blinded; SB, single-blinded; RA, randomized; F, Female; M, Male; CKD, Chronic Kidney Disease, HPLC, High Performance Liquid Chromatography; LC/MS, Liquid Chromatography Mass Spectrometry; GC-TOF MS, Gas Chromatography Time of Flight Mass Spectrometry; LCMS, Liquid Chromatography Mass Spectrometry; UPLC-MS/MS, Ultra Performance Liquid Chromatography Tandem Mass Spectrometry; Y, yes; N, no.

| Study | country | Design | sample | Participant health status | Trial duration (week) | Intervention sample size | Control sample size | Intervention mean age | Control mean age | Intervention ingredient [Fermentability, Viscosity] | Fiber type | Dose (g/day) | Control ingredient [Fermentability, Viscosity] | Dose (g/day) | Metabolite assessment method |
| --- | --- | --- | --- | --- | --- | --- | --- | --- | --- | --- | --- | --- | --- | --- | --- |
| Chang et al. 2023 | China | Parallel (RA/DB) | 45<br>22F,23M | CKD 3b–5 non-dialysis | 12 | 23 | 22 | 51.30±13.38 | 51.59±14.64 | Inulin +<br>(Low protein diet)<br>[Y, N] | Fructan | 10 | wheat starch +<br>(Low protein diet)<br>[N, N] | 10 | HPLC |
| Kemp et al. 2022 | Brazil | Parallel (RA/DB) | 25<br>11F,14M | CKD 5 Hemodialysis | 4 | 13 | 12 | 55.6±7.2 | 51.9±11.9 | Resistant starch type 2<br>(Hi-Maize 260)<br>[Y, N] | Resistant Starch | 16 | Manioc/Cassava flour<br>[Y, N] | 16 | LC-MS/MS |
| Ebrahim et al. 2022 | South Africa | Parallel (RA/DB) | 59<br>34F,25M | CKD Stages 3–5 non-dialysis non-diabetic | 14 | 23 | 22 | 40.6±11.4 | 41.3±12.0 | Beta-glucan<br>[Y, Y] | Hemicellulose | 3 | nothing | 0 | UPLC |
| Armani et al. 2021 | Brazil | Parallel (RA/DB) | 46<br>22F,24M | CKD 3b–5 non-dialysis non-diabetic | 12 | 23 | 23 | 61.9±11.4 | 53.4±16.0 | Fructooligosaccharide (FOS)<br>[Y, N] | Fructan | 12 | Maltodextrin<br>[N, N] | 12 | HPLC |
| de Andrade et al. 2021 | Brazil | Crossover (RA/DB) | 26<br>12F,14M | CKD 5 Peritoneal dialysis | 4 | 26 | 26 | 55±12 | 55±12 | Resistant starch type 2<br>(48%)<br>(Unripe Banana Flour)<br>[Y, N] | Resistant Starch | 21 | Waxy corn starch (Amioca)<br>[N, N] | 12 | HPLC |
| Birute 2021 | USA | Crossover (RA/DB) | 12<br>6F,6M | CKD 5 Hemodialysis | 4 | 12 | 12 | 55±10 | 55±10 | Inulin<br>[Y, N] | Fructan | F:10,<br>M:15 | Maltodextrin<br>[N, N] | F:10,<br>M:15 | UPLC-MS/MS |
| Gao et al. 2020 | USA | Crossover (RA/DB) | 8<br>4F,4M | CKD 5 Peritoneal dialysis | 8 | 8 | 8 | 68±14 | 68±14 | p-inulin<br>(oligofructose-enriched p-inulin)<br>[Y, N] | Fructan | 16 | Nothing | 0 | GC-TOF MS |
| Li et al. 2020 | China | Crossover (RA/DB) | 15<br>6F,9M | CKD 5 Continuous ambulatory peritoneal dialysis | 12 | 15 | 15 | 30.88±13.94 | 30.88±13.94 | inulin-type fructans<br>(50:50 mixture of long-chain inulin and oligofructose)<br>[Y, N] | Fructan | 10 | Maltodextrin<br>[N, N] | 10 | UPLC-MS/MS |
| Azevedo et al. 2020 | Brazil | Parallel (RA/DB) | 31<br>13F,18M | CKD 5 Hemodialysis | 4 | 15 | 16 | 56.0±7.5 | 53.5±11.5 | Resistant starch type 2<br>(Hi-Maize 260)<br>[Y, N] | Resistant Starch | 16 | Manioc/Cassava flour<br>[Y, N] | 16 | HPLC |
| Ramos et al. 2018 | Brazil | Parallel (RA/DB) | 46<br>19F,27M | CKD 3b–5 non-dialysis non-diabetic | 12 | 23 | 23 | 62.2±11.3 | 52.8±16.1 | Fructooligosaccharide (FOS)<br>[Y, N] | Fructan | 12 | Maltodextrin<br>[N, N] | 12 | HPLC |
| Esgalhado et al. 2018 | Brazil | Parallel (RA/DB) | 31<br>13F,18M | CKD 5 Hemodialysis | 4 | 15 | 16 | 56±7.5 | 53.5±11.5 | Resistant starch type 2<br>(Hi-Maize 260)<br>[Y, N] | Resistant Starch | 16 | Manioc/Cassava flour<br>[Y, N] | 16 | HPLC |
| Poesen et al. 2016 | Belgium | Crossover (RA/DB) | 40<br>28F,12M | CKD 3b–5 non-dialysis non-diabetic | 4 | 40 | 40 | 70±6 | 70±6 | Arabinoxylan oligosaccharides (AXOS)<br>[Y, N] | Hemicellulose | 20 | Maltodextrin<br>[N, N] | 13.4 | UPLC-MS/MS |
| Sirich et al.<br>2014 | USA | Parallel<br>(RA/SB) | 40<br>24F,16M | CKD 5<br>Hemodialysis | 6 | 20 | 20 | 54±14 | 58±13 | Resistant starch type 2<br>(Hi-Maze 260)<br>[Y, N] | Resistant Starch | 15 | Waxy corn<br>starch (Amioca)<br>[N, N] | 15 | LCMS |

### Basic characteristics of animal studies

The meta-analysis included 15 preclinical animal intervention studies, five of which contained two distinct intervention arms, yielding 20 separate comparisons (54–68). A total of 312 rodents were included, with 153 allocated to intervention and 133 to control groups. Five studies contributed two independent comparisons due to distinct intervention arms: CKD stage-specific groups in Biruete et al. (54); two fiber doses in Yang et al. (55) and Hu et al.(58); two distinct fiber types in Hung and Suzuki et al. (63); and separate CKD induction models (adenine vs STZ) in Alza’abi et al. (57). Most trials were conducted in Oman (n = 5) (56,57,60,62,64) and Japan (n = 3) (59,63,65) with additional studies performed in China (n = 2) (58,68), the United States (n = 2) (54,55), Australia (n= 1) (67) and Turkey (n = 1) (61). Study populations consisted of rat or mouse models of CKD and diabetic kidney disease (DKD). Different mouse and rat strains were used across the studies, including the Cy/+ rat model, Wistar rats, Sprague Dawley rats, mice from the Institution of Cancer Research, and BKS Cg Lepr db mice. CKD was induced using various approaches, including progressive kidney disease models by cyst formation, adenine-induced injury, streptozotocin- induced diabetes, 5/6x nephrectomy-induced renal injury, and a homozygous mutation in the leptin receptor gene. Some CKD models were induced at the same time as the intervention period, while others were established before the start of the intervention. Sample sizes ranged from 8 to 24 animals per study, with intervention and control groups generally balanced. Study durations varied between 3 and 12 weeks. The age of animals ranged from 6 to 22 weeks at baseline, and most studies included just male animals with one study conducted exclusively in female animals (60). Interventions involved supplementation with various dietary fibers, including inulin, xylooligosaccharides (XOS), resistant starch type 2, psyllium husk, guar gum, gum acacia, partially hydrolyzed guar gum, galacto-oligosaccharides, and oligosaccharide-enriched inulin, administered at concentrations ranging from 2% to 15% (w/w). Control groups received no placebo, except for one study that used cellulose (54) (see **Table 3b**).

**Table 3b.** Characteristic of included animal studies in meta-analysis Abbreviations: DB, double-blinded; SB, single-blinded; RA, randomized; F, Female; M, Male; CKD, Chronic Kidney Disease, HPLC, High Performance Liquid Chromatography; LC/MS, Liquid Chromatography Mass Spectrometry; GC-TOF MS, Gas Chromatography Time of Flight Mass Spectrometry; LCMS, Liquid Chromatography Mass Spectrometry; UPLC-MS/MS, Ultra Performance Liquid Chromatography Tandem Mass Spectrometry; quadruple time-of-flight, Q-TOF, Y, yes; N

| Study | Country | Animal Model | CKD Induction Methd | Trial duration (week) | Intervention sample size | Control sample size | Intervention and placebo mean age (week) | Intervention ingredient [Fermentability, Viscosity] | Fiber type | Dos e (% (w/ w)) | Control ingredient [Fermentability, Viscosity] | Metabo lite assess ment method |
| --- | --- | --- | --- | --- | --- | --- | --- | --- | --- | --- | --- | --- |
| He et al. 2024 | China | Institution of Cancer Research mice | streptozotocin-induced (diabetic) | 6 | 24 | 12 | 6 | Inulin [Y, N] | Fructan | 2.5 | Nothing | UPLC-MS/MS |
| Snelson et al. 2024 | Australia | BKS.Cg-Leprdb mice | Homozygous Mutation ( leptin receptor gene) | 10 | 12 | 6 | 6 | Resistant starch type 2 (Hi-Maze 260) [Y, N] | Resistant Starch | 12.5 | Nothing | HPLC/ MS |
| Biruet e et al. 2023 (A) | USA | Cy/+ rat Model | Progressive kidney disease by cyst formation | 10 | 12 | 11 | 22 | Inulin [Y, N] | Fructan | 10 | Cellulose [N, N] | LC-MS |
| Biruet e et al. 2023 (B) | USA | Cy/+ rat Model | Progressive kidney disease by cyst formation | 12 | 12 | 12 | 22 | Inulin [Y, N] | Fructan | 10 | Cellulose [N, N] | LC-MS |
| Hu et al. 2023 (A) | China | Sprague Dawley Rats | 5/6x nephrectomy-induced | 4 | 10 | 5 | 10 | Psyllium seed husk (PSH) [N,Y] | Hemicellulose | 2.5 | Nothing | Elisa kit |
| Hu et al. 2023 (B) | China | Sprague Dawley Rats | 5/6x nephrectomy-induced | 4 | 10 | 5 | 10 | Psyllium seed husk (PSH) [N, Y] | Hemicellulose | 5 | Nothing | Elisa kit |
| Lakshamanan et al. 2021 | Oman | Sprague Dawley Rats | adenine-induced (0.75%) | 4 | 6 | 6 | 10 | Gum acacia [Y, Y] | Gum | 15 | Nothing | LC-MS |
| Melekoglu et al. 2021 | Turkey | Sprague Dawley Rats | adenine-induced (0.2%) | 4 | 8 | 8 | Not reported | Oligofructose-enriched inulin [Y, N] | Oligosacch aride | 5 | Nothing | HPLC |
| Al-Asmakh et al. 2020 | Oman | Wistar Rats | adenine-induced (0.75%) | 4 | 6 | 6 | 10.5 | Gum acacia [Y, Y] | Gum | 15 | Nothing | LC-MS |
| Tani et al. 2020 | Japan | Sprague-Dawley Rats | 5/6x nephrectomy-induced | 5 | 8 | 7 | 12 | Partially hydrolyzed guar gum [Y, N] | Gum | 3 | Nothing | Q-TOF |
| Yang et al. 2018 (A) | USA | C57BL/6J Mice | adenine-induced (0.2%) | 3 | 8 | 4 | 10 | XOS [Y, N] | Oligosacch aride | 2 | Nothing | HPLC & GCMS |
| Yang et al. 2018 (B) | USA | C57BL/6J Mice | adenine-induced (0.2%) | 3 | 8 | 4 | 10 | XOS [Y, N] | Oligosacch aride | 7 | Nothing | HPLC & GCMS |
| Hung and Suzuki et al. 2018 (A) | Japan | Institution of Cancer Research mice | adenine induced (0.25%) | 3 | 4 | 4 | 10 | Unmodified guar gum (GG) [Y, Y] | Gum | 10 | Nothing | LC-MS |
| Hung and Suzuki et al. 2018 (B) | Japan | Institution of Cancer Research mice | adenine induced (0.25%) | 3 | 5 | 4 | 10 | Partially hydrolyzed GG (PHGG) [Y, N] | Gum | 10 | Nothing | LC-MS |
| Al Za`abi et al. 2016 (A) | Oman | Wistar Rats | adenine-induced (0.25%) | 5 | 6 | 6 | 10.5 | Gum acacia [Y, Y] | Gum | 15 | Nothing | HPLC |
| Al Za`abi et al. 2016 (B) | Oman | Wistar Rats | adenine-induced (0.25%) + Streptozotocin-induced (diabetic) | 5 | 6 | 6 | 10.5 | Gum acacia [Y, Y] | Gum | 15 | Nothing | HPLC |
| Al Suleimani et al.<br>2015 | Oman | Wistar Rats | adenine-induced<br>(0.25%) | 5 | 8 | 8 | Not reported | Gum acacia<br>[Y, Y] | Gum | 15 | Nothing | LC-MS |
| Ali et al.<br>2014 | Oman | Wistar Rats | adenine-induced<br>(0.75%) | 4 | 6 | 6 | 10.5 | Gum acacia<br>[Y, Y] | Gum | 15 | Nothing | HPLC |
| Ali et al.<br>2014 | Oman | Wistar Rats | adenine-induced<br>(0.75%) | 4 | 6 | 6 | 10.5 | Gum acacia<br>[Y, Y] | Gum | 15 | Nothing | HPLC |
| Furuse et al.<br>2014 | Japan | Sprague Dawley<br>Rats | 5/6x nephrectomy-induced | 2 | 6 | 7 | 10 | Galacto-Oligosaccharide<br>[Y, N] | Oligosacch<br>aride | 5 | Nothing | HPLC |

### Risk of bias assessment

Risk of bias was assessed separately for human and animal studies. Overall, the majority of human trials were judged to be at low risk of bias based on Cochrane risk-of-bias tool for randomized trials (RoB 2) (27), with the exception of five studies(42–44,48,53), which were rated as having some concerns or a high risk of bias (**Figures 2a-b**). For animal studies, the risk of bias assessment using, risk of bias tool SYRCLE (23) indicated that most trials demonstrated a some concerns or high risk of bias across evaluated domains, reflecting generally low methodological quality based on the assessment tool (**Figures 3a-b**). Most animal studies provided limited methodological detail, resulting in unclear or high risk of bias across multiple domains. Specifically, randomization procedures and blinding of caregivers or outcome assessors were rarely described, and baseline characteristics were generally not reported or accounted for in the analyses. Allocation concealment was also not documented in nearly all studies. Overall, the reporting quality of animal studies was insufficient to fully assess potential sources of bias.

**Figure 2.**
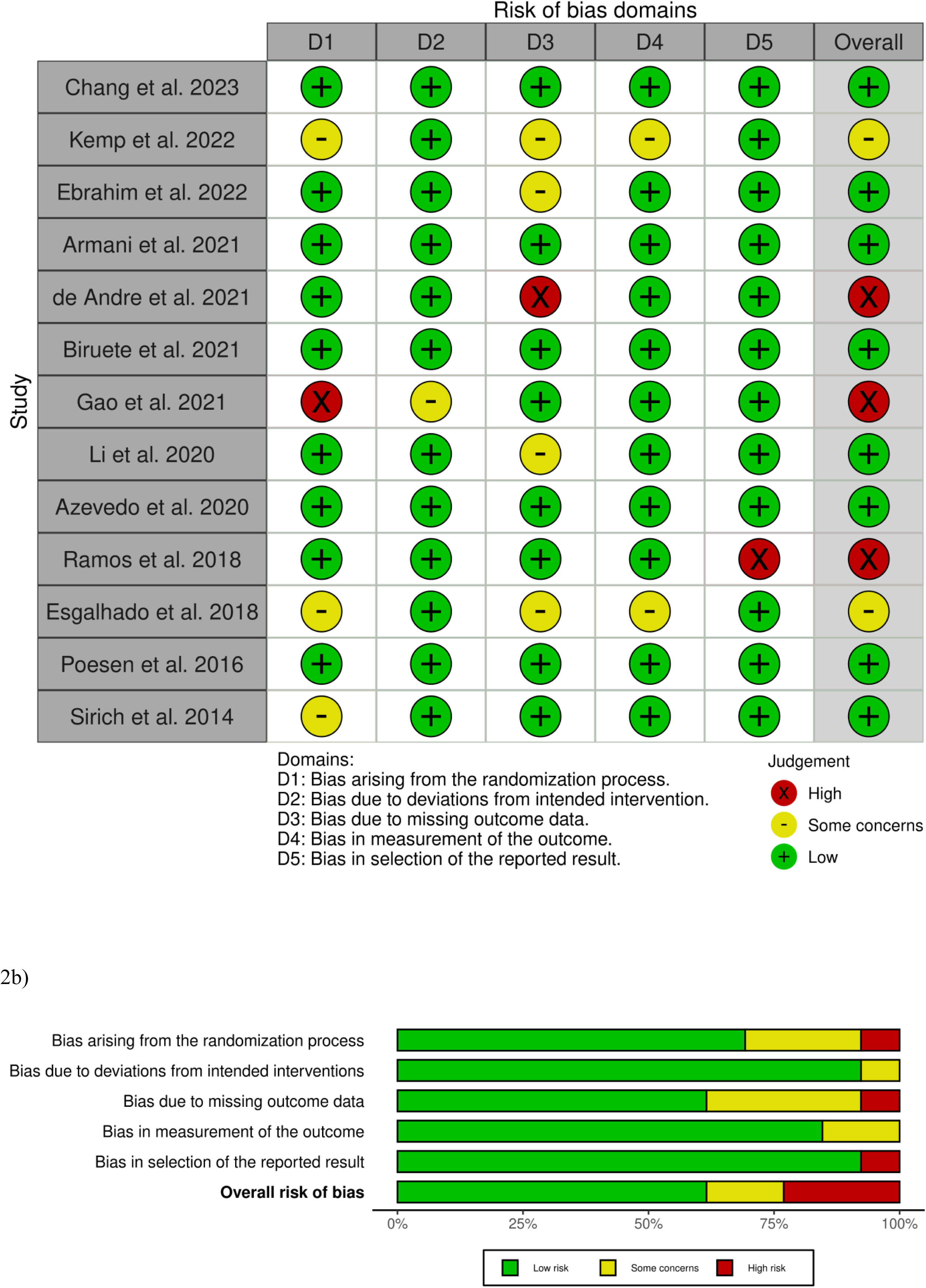
Quality assessment of human studies according to Cochrane risk-of-bias 2 (RoB-2) a) Traffic light plot, b) Summary plot

**Figure 3.**
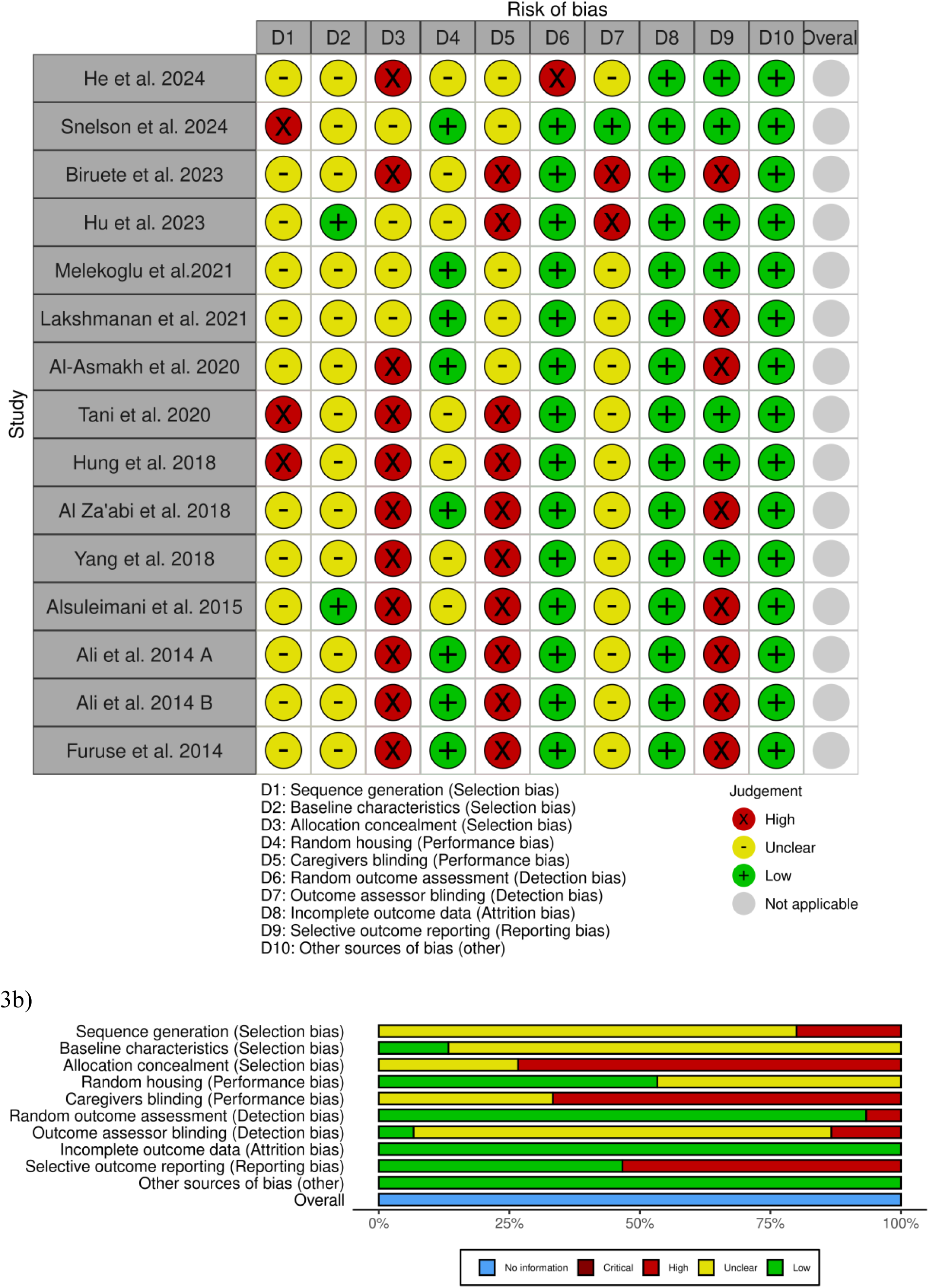
Quality assessment of human studies according to Cochrane risk-of-bias 2 (RoB-2) a) Traffic light plot, b) Summary plot

## Meta-analysis of RCTs

### Effects of dietary fiber supplementation on total serum/plasma IAA

A meta-analysis of four RCTs (43,47,51,53) found that supplementation with dietary fiber; resistant starch type 2, FOS and β-glucan did not result in a significant change in total serum/plasma IAA concentrations. The pooled WMD was 0.003 mg/dL (95% CI: −0.02 to 0.03; p = 0.86). Moderate heterogeneity was observed across studies (I² = 49.6%, p = 0.11). No subgroup analysis was conducted due limited number of studies contributing data to this outcome for each subgroup (**Figure 4a**).

**Figure 4.**
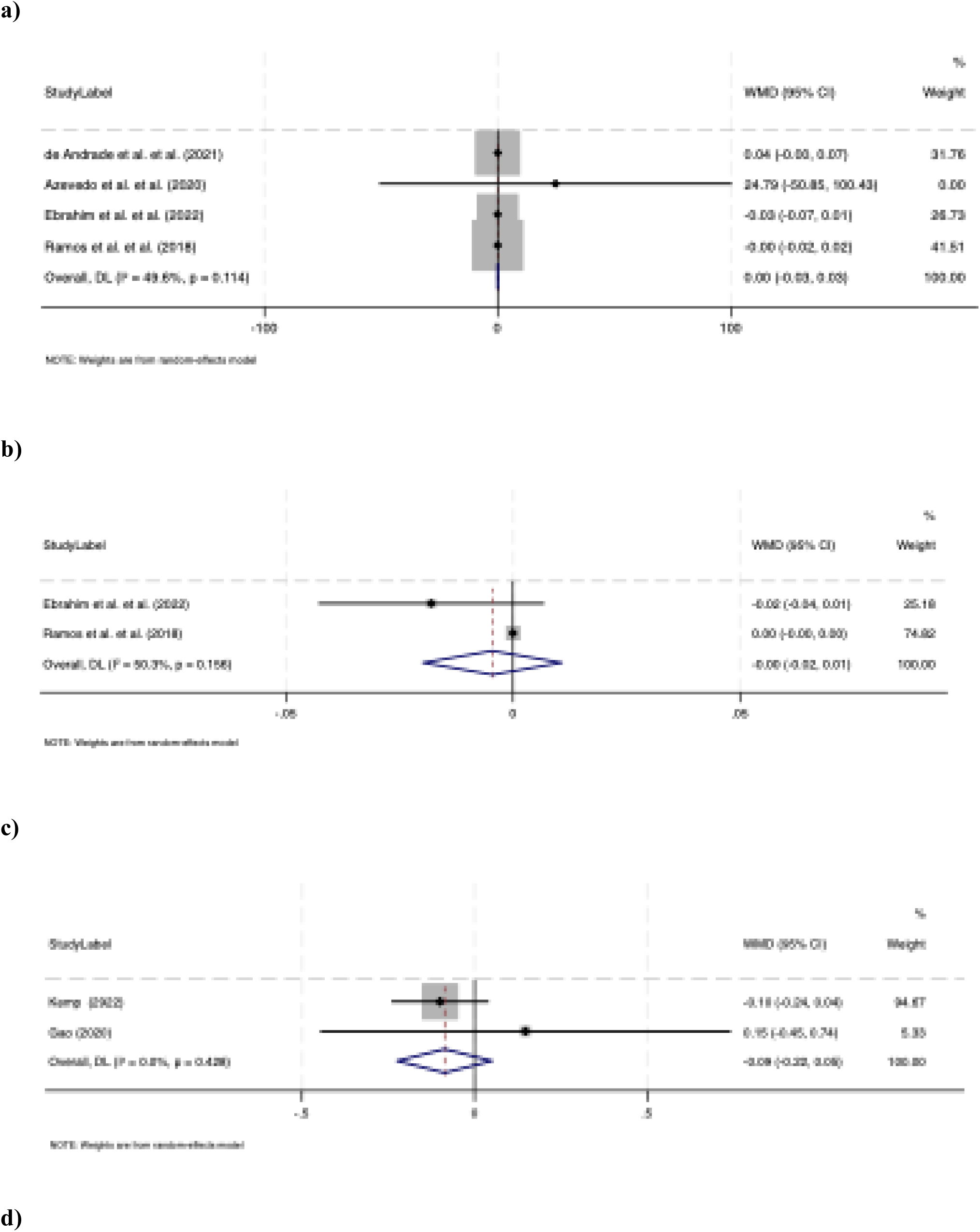
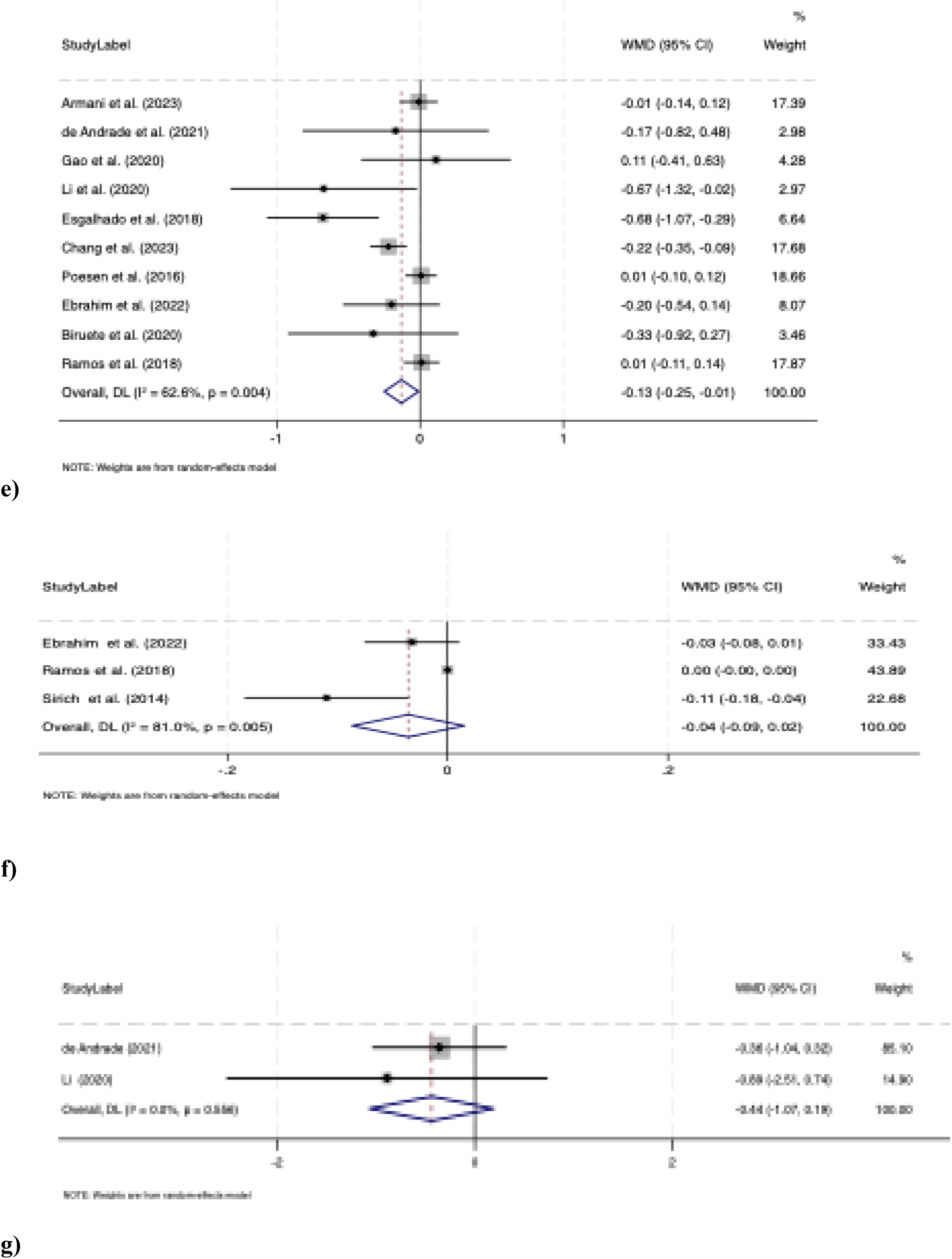
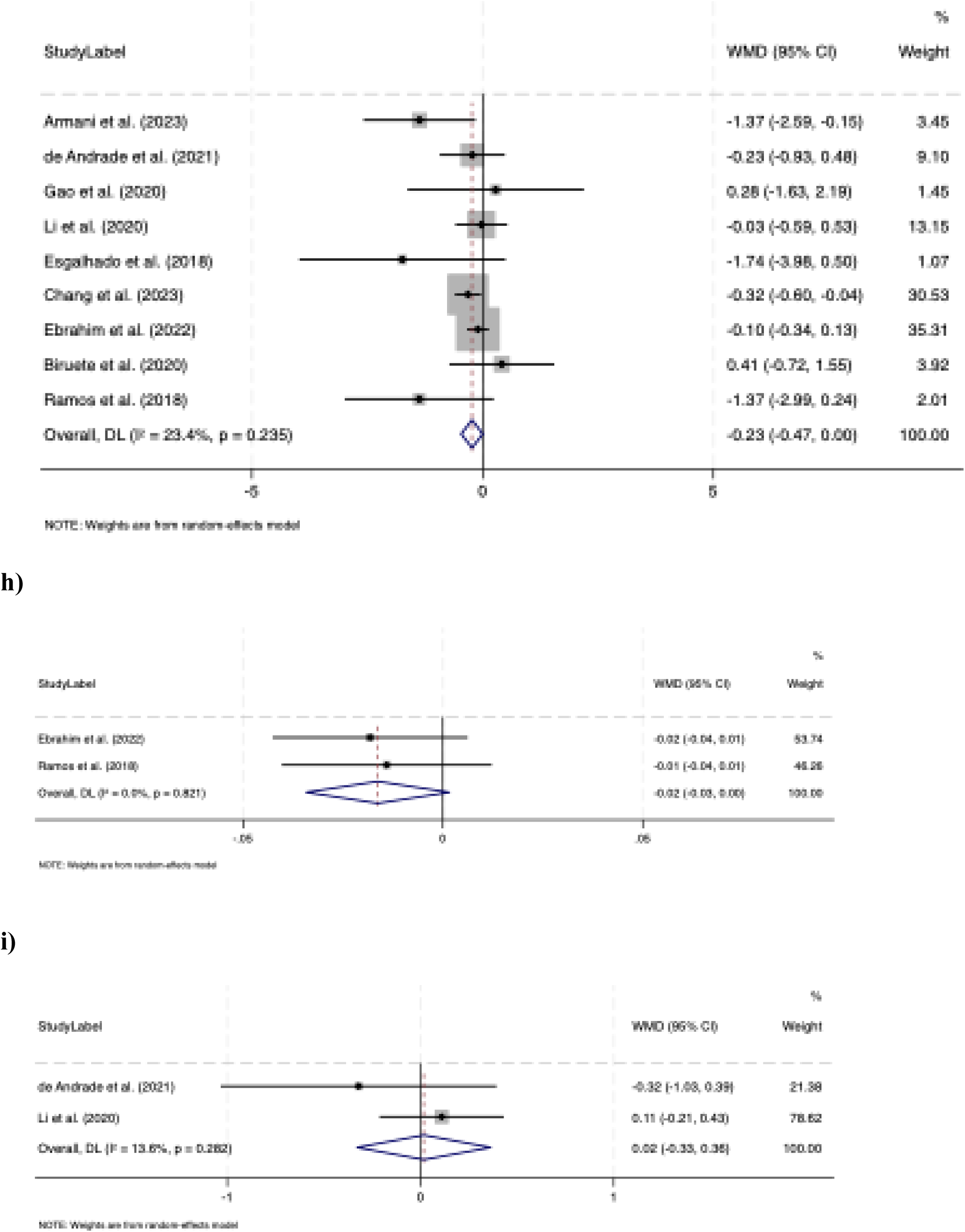
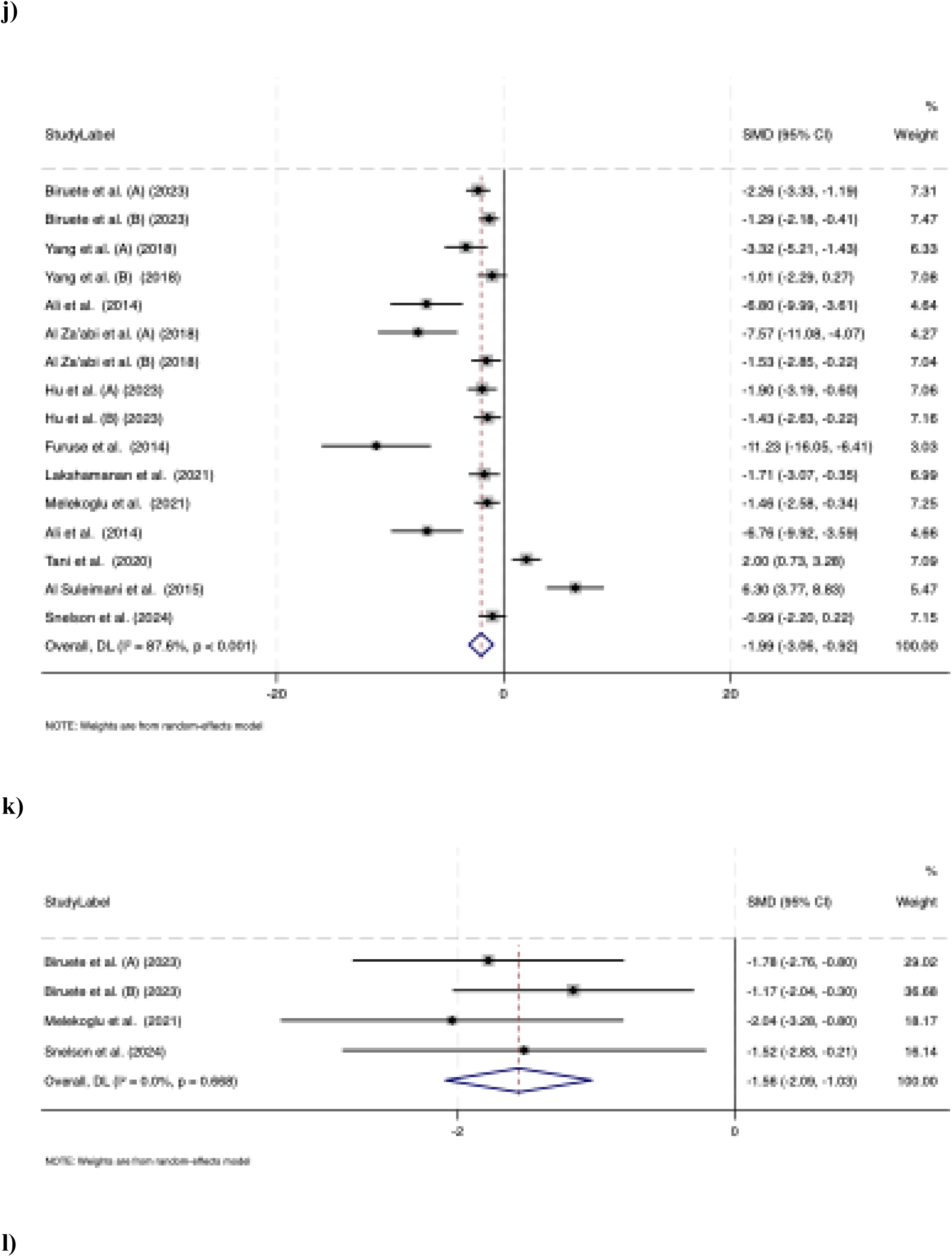
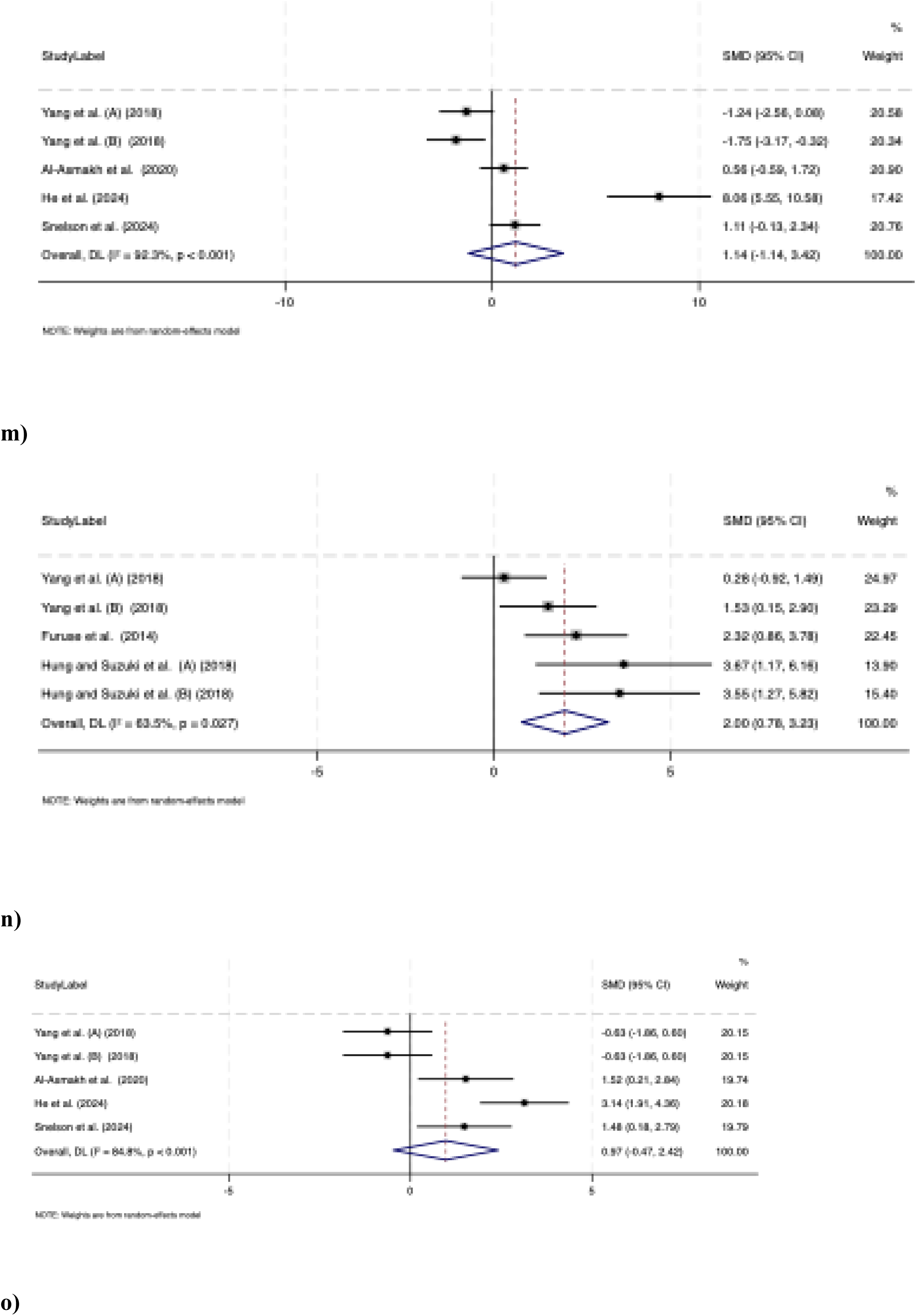
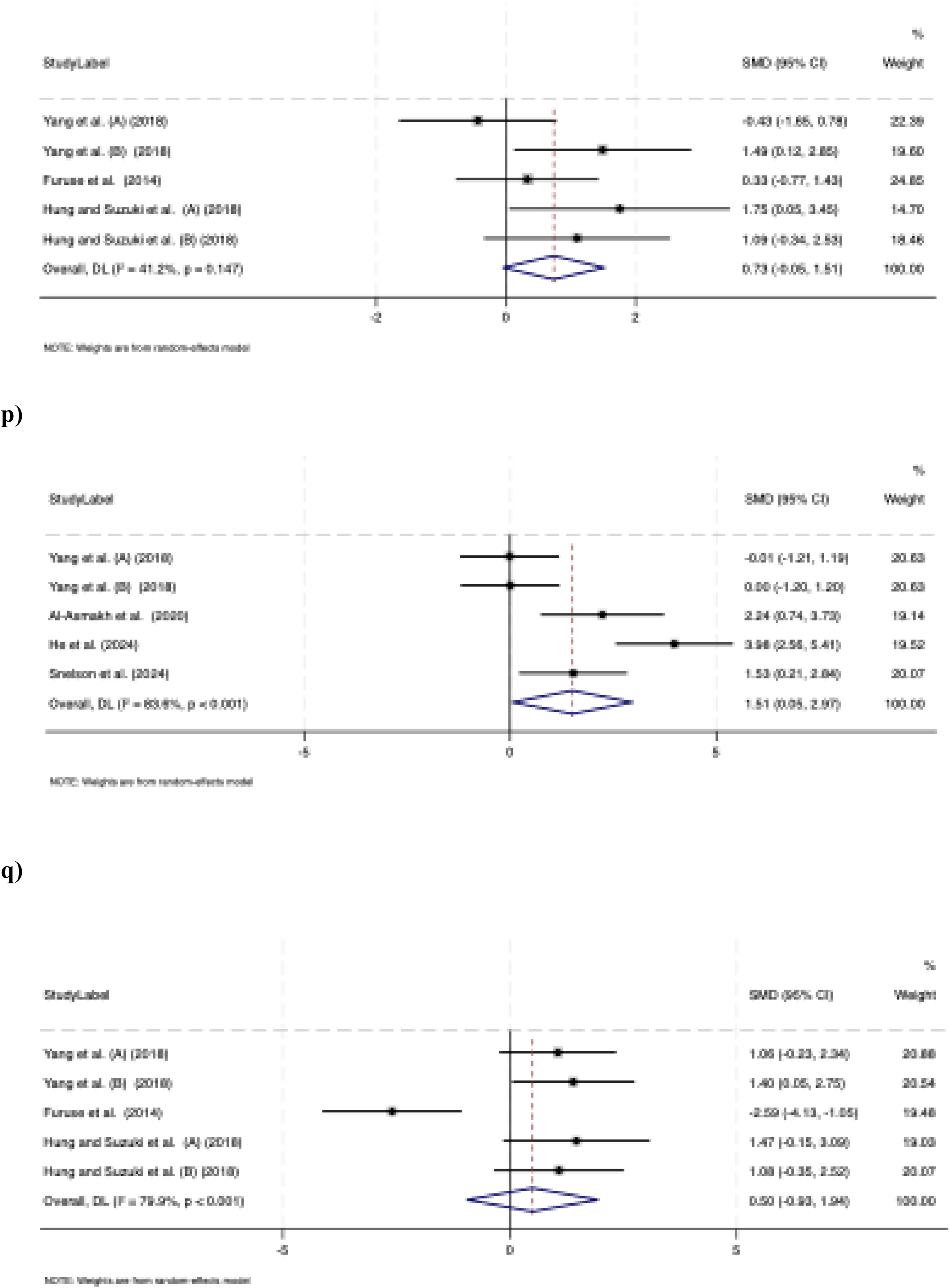
Forest plot detailing the random-effects model of weighted mean difference (WMD) with 95% confidence intervals (CIs) for the effect of dietary fiber supplementation on gut microbiota metabolites (mg/dL) in adults with Chronic Kidney Disease (CKD) **(a-i),** including: (**a**) Total plasma/serum Indole-3-acetic acid (IAA), (**b**) Free plasma/serum Indole-3-acetic acid (IAA), (**c**) Plasma/serum trimethylamine-N-oxide (TMAO), (**d**) Total serum/plasma indoxyl sulfate (IS), (**e**) Free serum/plasma indoxyl sulphate (IS), (**f**) Urinary indoxyl sulfate (IS), (**g**) Total serum/plasma p-cresyl sulfate (pCS), (**h**) Free serum/plasma p-cresyl sulfate (pCS), (**i**) Urinary serum/plasma p-cresyl sulfate (pCS). Forest plot detailing the random-effects model of standard mean difference (SMD) with 95% confidence intervals (CIs) for the effect of dietary fiber supplementation on gut microbiota metabolites in rodents of Chronic Kidney Disease (CKD) model (j-q) including: (**j**) Total serum/plasma indoxyl sulfate (IS), (**k**) serum/plasma p-cresyl sulfate (pCS), (**l**) serum/plasma acetate, (**m**) cecal acetate, (**n**) serum/plasma butyrate, (**o**) cecal butyrate, (**p**) serum/plasma propionate, **(q**) cecal propionate.

### Effects of dietary fiber supplementation on free serum/plasma IAA

A meta-analysis of two RCTs (51,53) found that supplementation with dietary fiber FOS and β-glucan did not result in a significant change in free serum/plasma IAA concentrations. The pooled WMD was - 0.005 mg/dL (95% CI: −0.02 to 0.01; p = 0.56). Moderate heterogeneity was observed across studies (I² = 50.3%, p = 0.15). Subgroup analyses were not performed for this outcome because too few studies contributed data to allow meaningful stratification (**Figure 4b**).

### Effects of dietary fiber supplementation on total serum/plasma TMAO

A meta-analysis including two RCTs (42,44) showed that supplementation with dietary fiber, resistant starch type 2 and oligofructose-enriched inulin, did not significantly affect circulating TMAO concentrations. The pooled WMD was −0.08 mg/dL (95% CI: −0.22 to 0.05; p = 0.215). No meaningful between-study heterogeneity was detected (I² = 0.0%, p = 0.428) (**Figure 4c**). Subgroup analyses were not performed for this outcome because too few studies contributed data to allow meaningful stratification.

### Effects of dietary fiber supplementation on total serum/plasma IS

A meta-analysis of nine RCTs (41,43,44,46,48–53) showed that dietary fiber supplementation, including FOS, resistant starch type 2, inulin-type fructans, and β-glucan significantly reduced total serum or plasma IS concentrations. The pooled WMD was −0.13 mg/dL (95% CI: −0.25 to -0.01; p = 0.03). Substantial between-study heterogeneity was observed (I² = 62.6%, p = 0.004) (**Figure 4d**).

### Subgroup Analysis of effects of dietary fiber supplementation on total serum/plasma IS

Results of subgroup analyses conducted for total serum/plasma IS are summarized in **Table 4a** by dietary supplementation type, intervention dose, duration and dialysis status. Subgroup analyses were not feasible by dietary fiber fermentability and viscosity due to limited number of studies per subgroup. In subgroup analyses by fiber type, resistant starch type 2 significantly reduced total serum/plasma IS levels in patients with CKD (WMD: −0.49 mg/dL; 95% CI: −0.97 to −0.01; p = 0.04) across two studies (43,48). I n contrast, fructan and beta-glucan supplementation did not result in statistically significant reductions in IS levels (p = 0.20 and p = 0.24, respectively). Subgroup analysis based on trial duration and dose did not show significant reductions (p=0.62 and 0.07, respectively). Subgroup analysis by dialysis status showed that fiber supplementation resulted in a significant reduction in IS among patients receiving dialysis (WMD: −0.36 mg/dL, 95% CI: −0.68, −0.04; p = 0.02), whereas no significant effect was observed in non-dialysis participants (p = 0.221) (**Figures S1a-d**).

**Table 4a.**
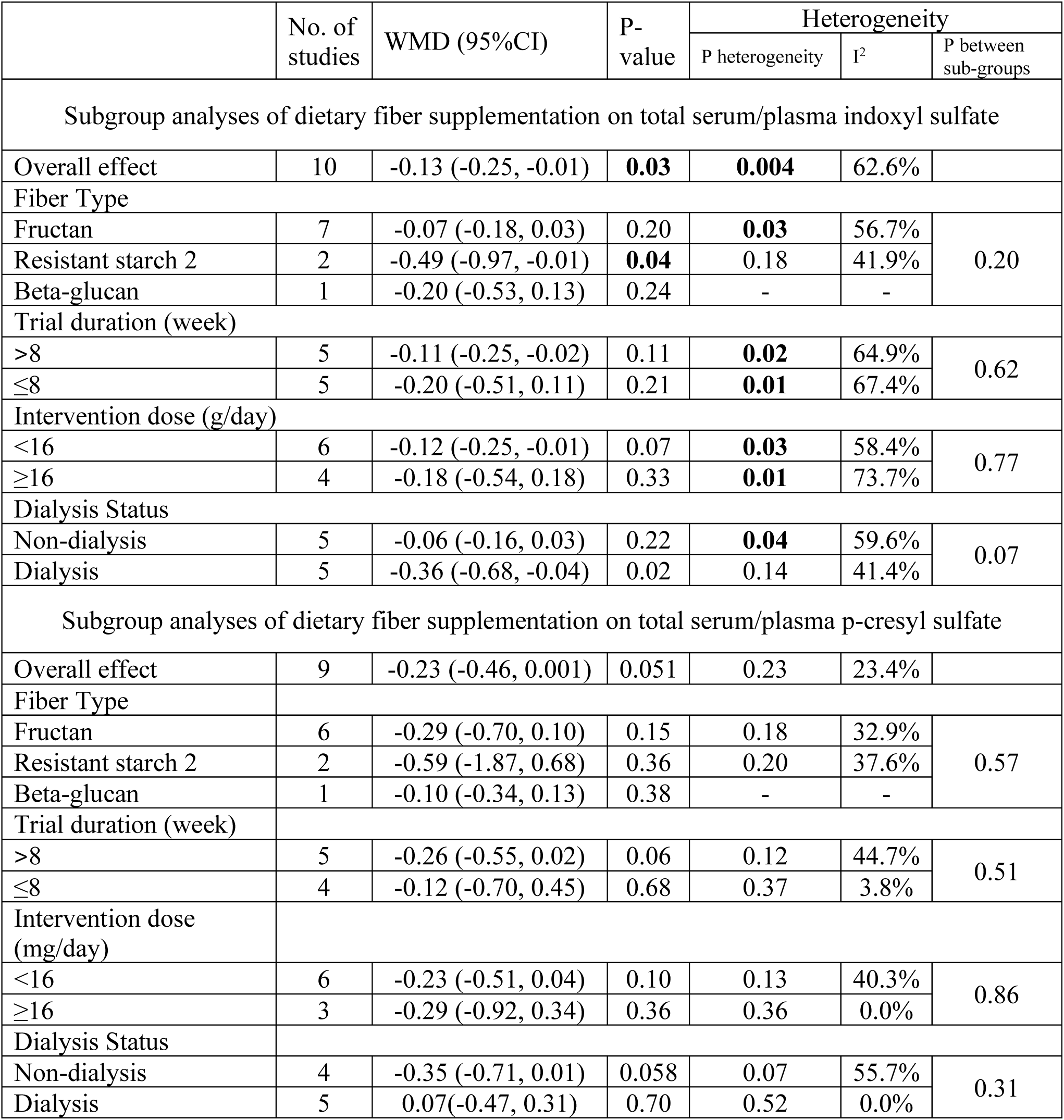
Subgroup analyses of dietary fiber supplementation on gut micribiota metabolites in adults with CKD. Abbreviations: CI, confidence interval; WMD, weighted mean differences.

**Table 4b.**
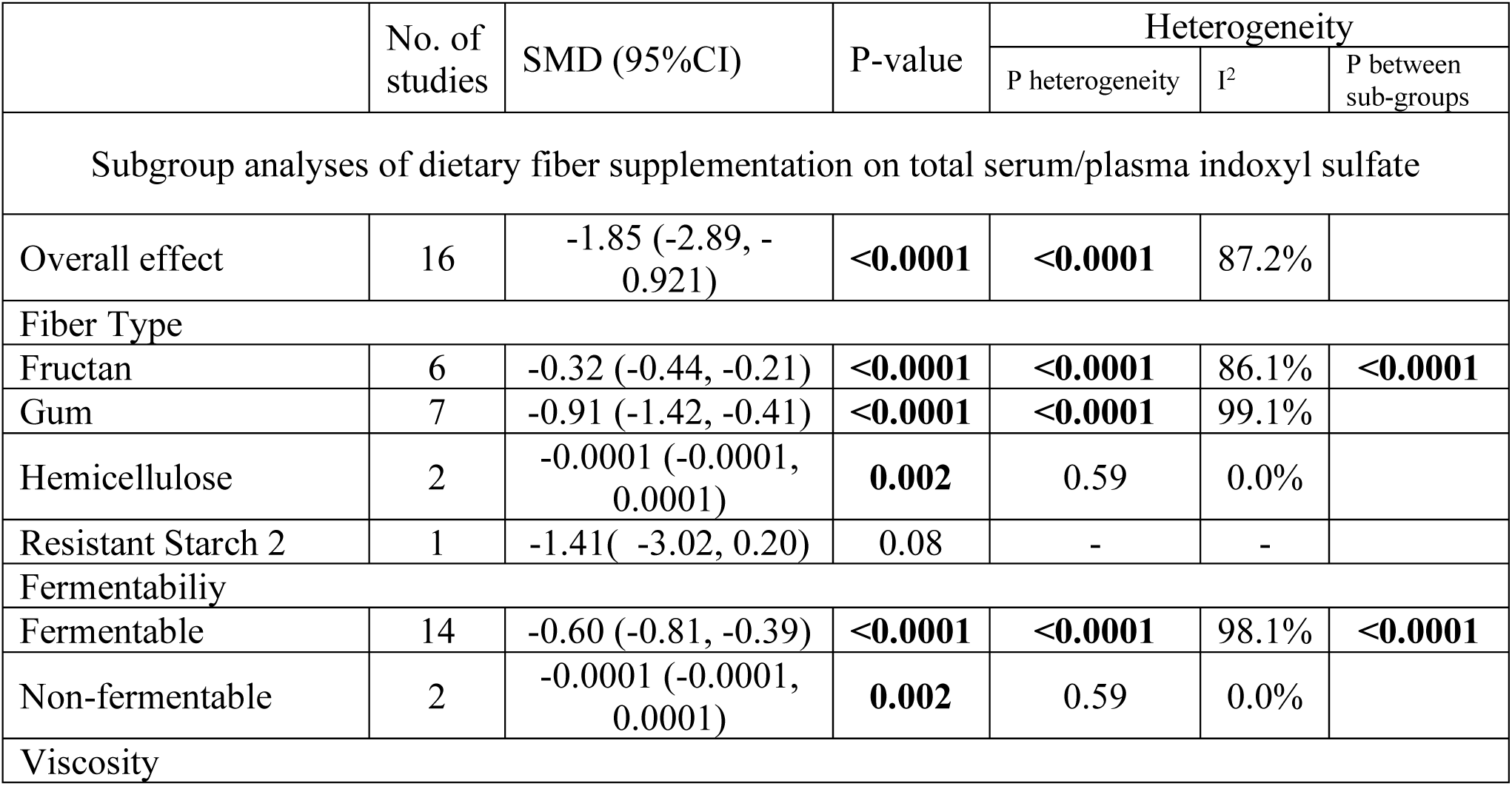
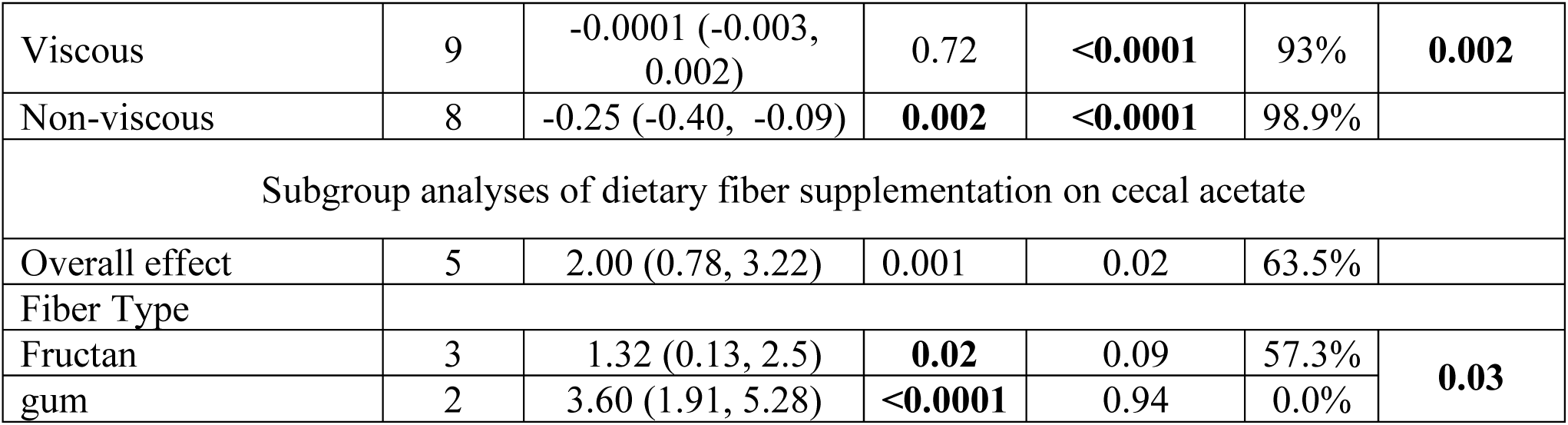
Subgroup analyses of dietary fiber supplementation on gut micribiota metabolites in animal models of CKD. Abbreviations: CI, confidence interval; SMD, standardized mean differences

### Effects of dietary fiber supplementation on free serum/plasma IS

A meta-analysis of three RCTs (45,51,53) indicated that supplementation with dietary fiber, did not change free serum/plasma IS concentrations. The pooled WMD was −0.03 mg/dL (95% CI: −0.08 to 0.01; p = 0.17). Substantial between-study heterogeneity was observed (I² = 81.0%, p = 0.005) (**Figure 4e**). Due to few numbers of studies, subgroup analysis was not performed.

### Effects of dietary fiber supplementation on urinary IS

A meta-analysis of two RCTs (43,46) found that supplementation with dietary fiber, including resistant starch type 2 and inulin-type fructans, did not significantly affect urinary IS concentrations. The pooled WMD was −0.43 mg/dL (95% CI: −1.06 to 0.18; p = 0.17), with no evidence of between-study heterogeneity (I² = 0.0%, p = 0.55) (**Figure 4f**). Due to limited numbers of studies, subgroup analysis was not conducted.

### Effects of dietary fiber supplementation on total serum/plasma pCS

A meta-analysis of nine RCTs (41,43,44,46,48,49,51–53) showed that dietary fiber supplementation, including FOS, resistant starch type 2, inulin-type fructans, and β-glucan reduced total serum or plasma pCS concentrations which did not reach statistical significance. The pooled WMD was −0.23 mg/dL (95% CI: −0.46 to 0.001; p = 0.051). Low between-study heterogeneity was observed (I² = 23.4%; p = 0.23) (**Figure 4g**).

### Subgroup Analysis of effects of dietary fiber supplementation on total serum/plasma pCS

Results of the subgroup analyses for total serum/plasma pCS are summarized in **Table 4a** according to dietary supplementation type, intervention dose, duration, and dialysis status. Subgroup analyses based on fiber fermentability and viscosity were not conducted because too few studies contributed data to these categories. Similarly, subgroup analysis showed that dietary fiber supplementation did not significantly reduce total serum/plasma pCS concentrations across supplementation type, dose, duration, or dialysis status (**Figures S1e-h**). Post hoc subgroup analysis was done after sensitivity analysis showed the impact of studies by Biruete et al. (52) and Armani et al. (41) which resulted in statistically meaningful reduction in pCS after supplementation by fructans (WMD: -0.27, CI95%: -0.52, -0.03; p = 0.02) unlike resistant starch and beta-glucan (p = 0.36 and p = 0.38, respectively).

### Effects of dietary fiber supplementation on free serum/plasma pCS

A meta-analysis of two RCTs (51,53) indicated that supplementation with dietary fiber, did not change free serum/plasma pCS concentrations. The pooled WMD was −0.02 mg/dL (95% CI: −0.03 to 0.002; p = 0.07). No between-study heterogeneity was observed (I² = 0.0%, p = 0.82) (**Figure 4h**). Due to few numbers of studies, subgroup analysis was not performed.

### Effects of dietary fiber supplementation on urinary pCS

A meta-analysis of two RCTs (43,46) indicated that supplementation with dietary fiber, did not change urinary pCS concentrations. The pooled WMD was −0.01 mg/dL (95% CI: −0.32 to 0.36; p = 0.92). Low between-study heterogeneity was observed (I² = 13.6%, p = 0.28) (**Figure 4i**). Subgroup analyses were not performed for this outcome because too few studies contributed data to allow meaningful stratification.

### Sensitivity analysis

Bias and sensitivity analysis results are summarized in **Table 5a and figures S2a-i.** No significant publication bias was detected for total and free serum or plasma IAA, total serum/plasma TMAO, urinary and free serum/plasma IS, and pCS. Sensitivity analyses indicated that the pooled estimate for total serum/plasma IS was influenced by individual studies, including Li et al. (46) (WMD: -0.11, CI95%: -0.22, 0.006), Esgalhado et al. (48) (WMD: -0.07, CI95%: -0.17, 0.01), Ebrahim et al. (51) (WMD: -0.12, CI95%: -0.25, 0.002) and Chang et al. (49) (WMD: -0.10, CI95%: -0.32, 0.02) suggesting potential instability of the overall effect. For total serum/plasma pCS, sensitivity was observed in the study by Biruete et al. (52) (WMD: -0.25, CI95%: -0.49, -0.02) and Armani et al. (41) (WMD: -0.18, CI95%: -0.34, -0.02) indicating a potential influence on total serum/plasma pCS outcomes (Table 5a). Exclusion of these studies shifted the pooled estimate and resulted in a statistically significant reduction in pCS (WMD: -0.19, CI95%: -0.36, -0.32).

**Table 5a.**
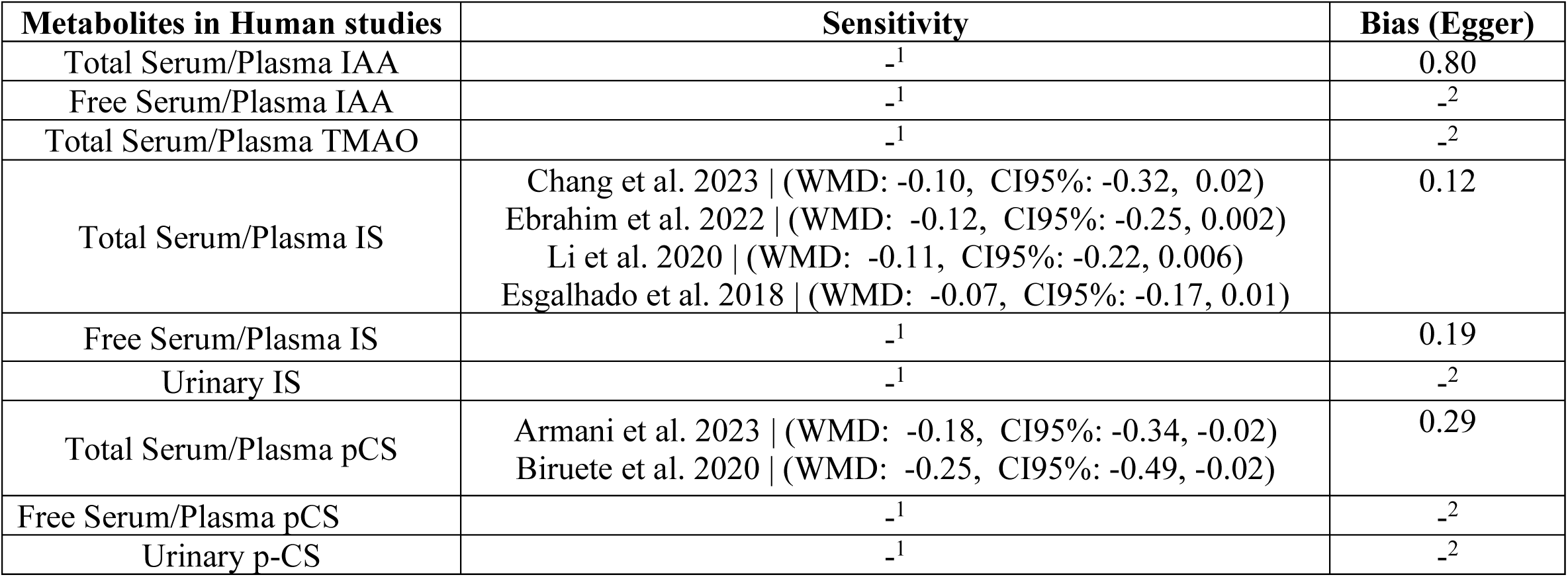
Bias sensitivity analysis and publication bias in RCTs 1. Not detected 2. Not enough number of studies to conduct sensitivity analysis or Egger’s test

### Publication bias

No evidence of publication bias was observed for IAA (p = 0.90), total serum or plasma IS (p = 0.12), or total serum or plasma pCS (p = 0.29) **(Table 5a)**. Assessment of publication bias was not feasible for TMAO, urinary IS, or free serum or plasma IS and pCS due to the limited number of studies. Visual inspection of funnel plots suggested relative symmetry across outcomes, indicating a low likelihood of small-study effects or selective reporting. Collectively, these findings support the robustness of the pooled estimates (**Fig 5a-i**).

**Figure 5a-q.**
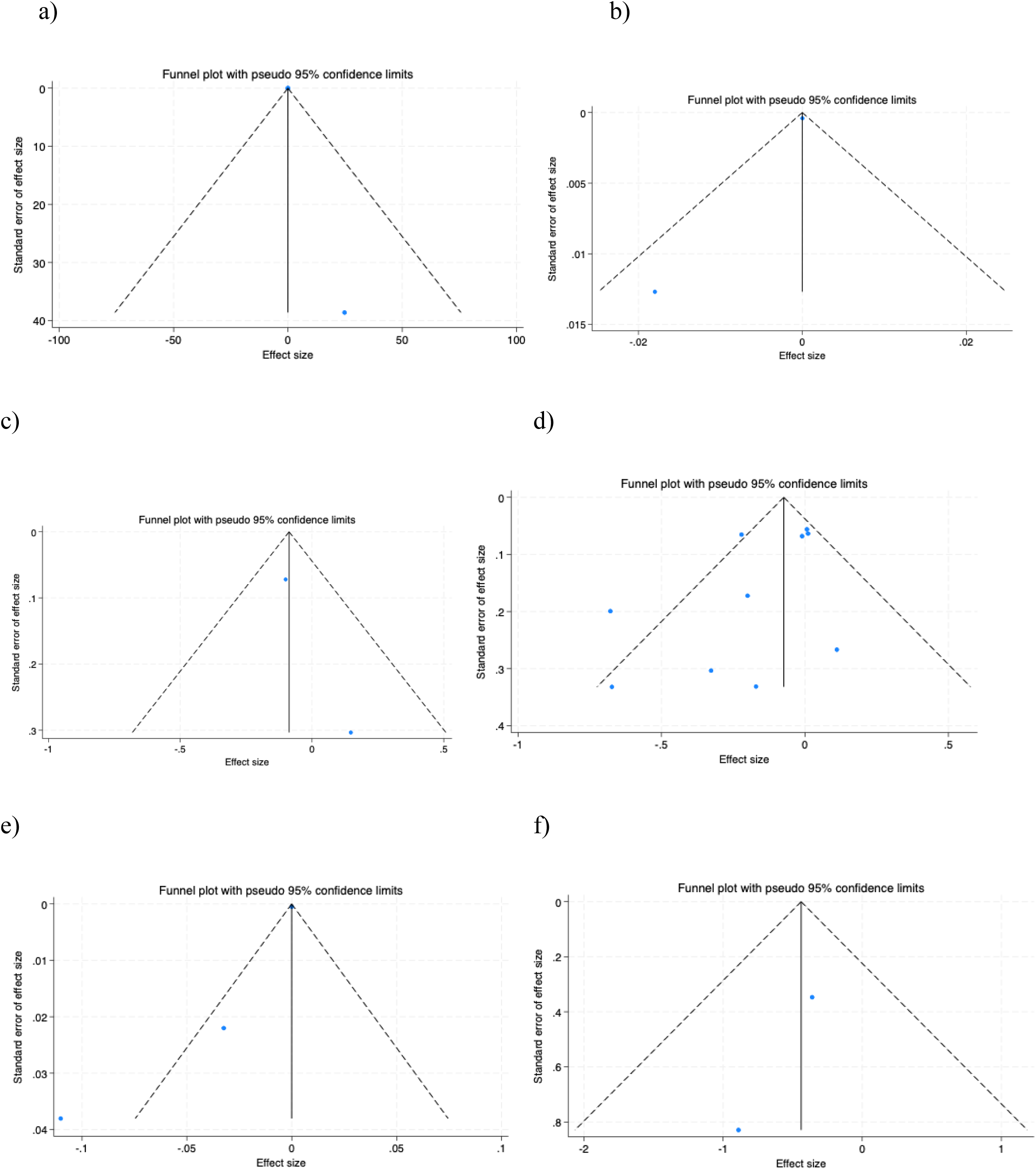
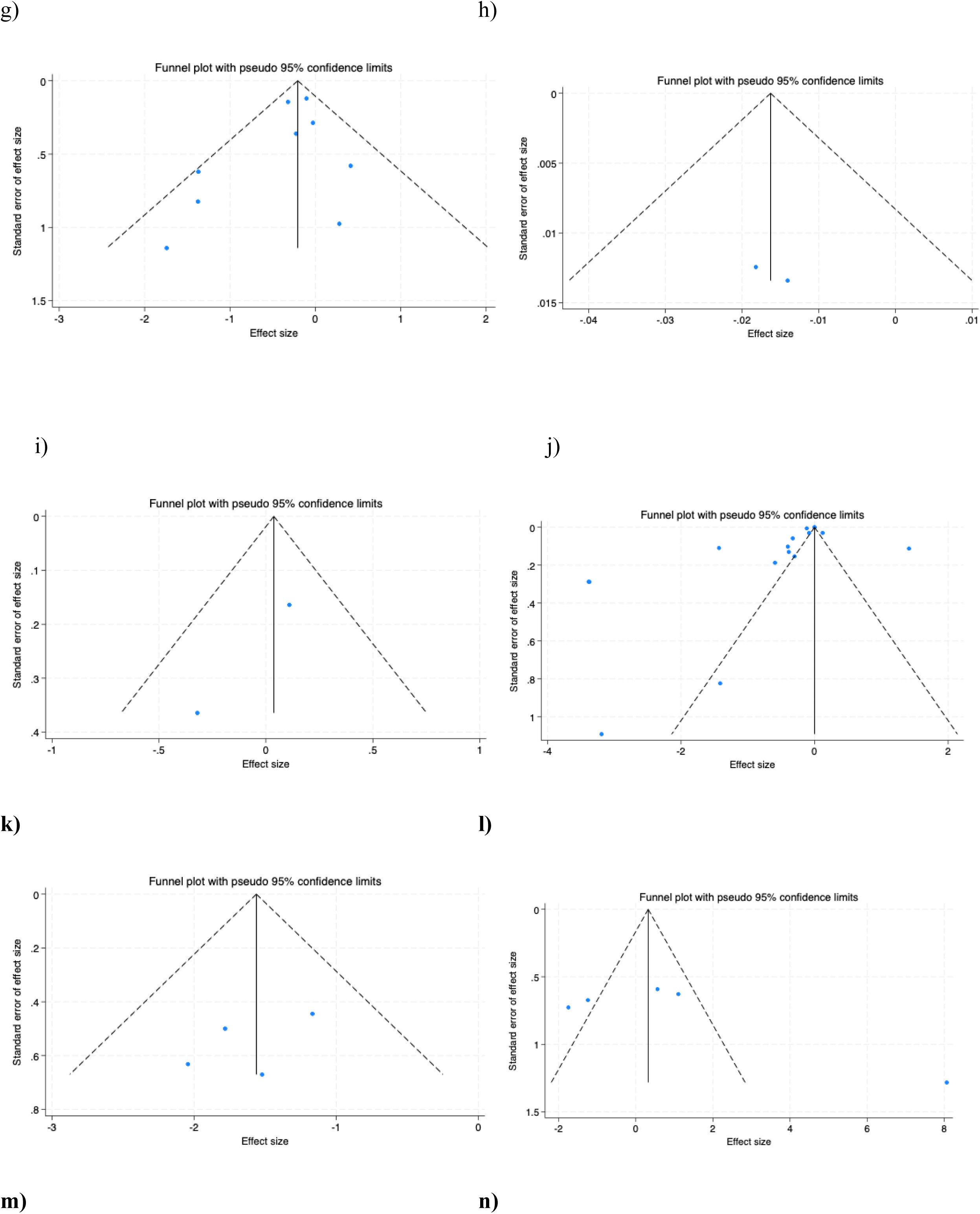
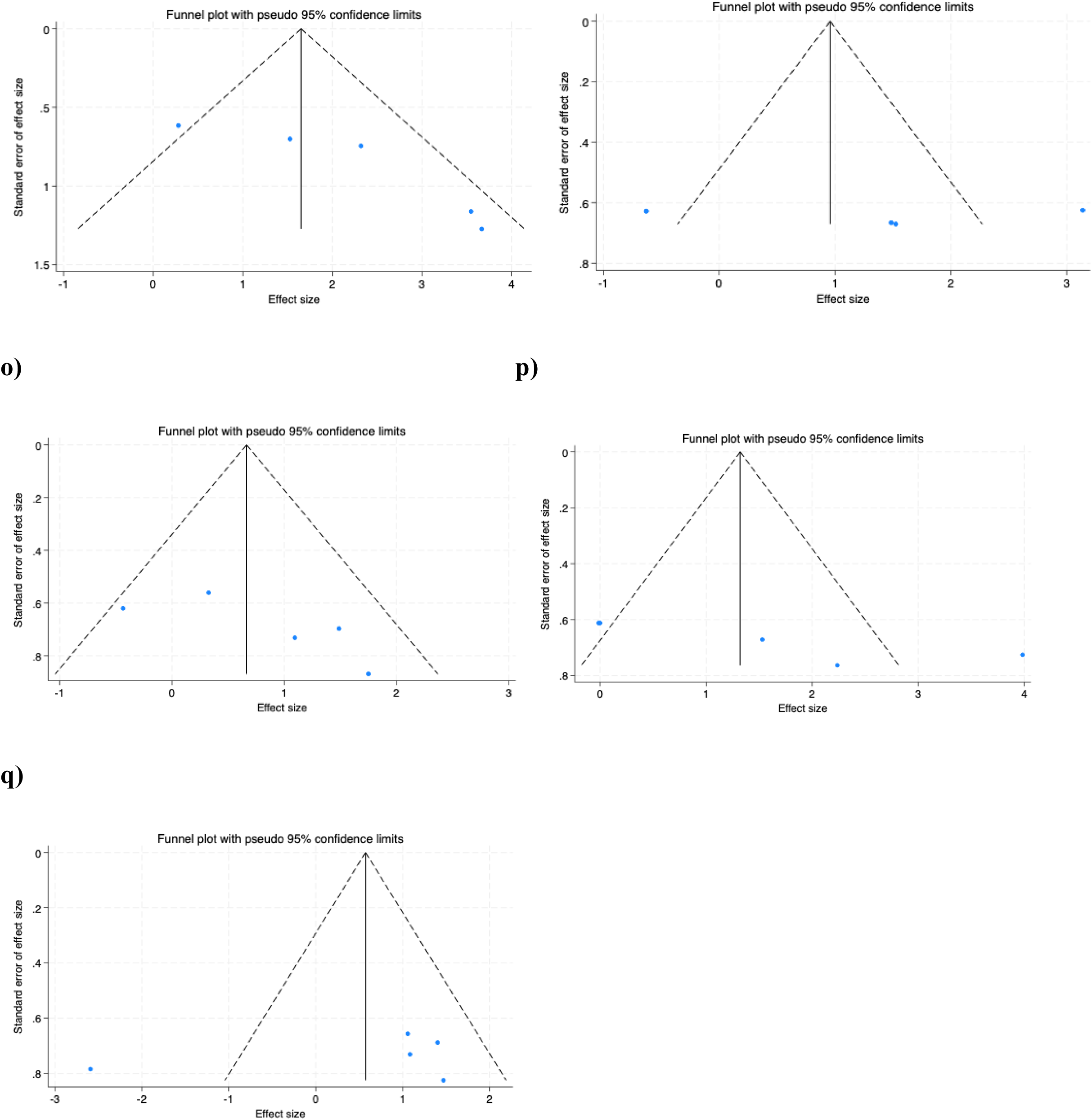
Funnel plots of publication bias for the effect of dietary fiber supplementation on on gut microbiota metabolites in adults with Chronic Kidney Disease (CKD) including: (**a**) Total serum/plasma Indole-3-acetic acid (IAA), (**b**) Free plasma/serum Indole-3-acetic acid (IAA), (**c**) Plasma/serum trimethylamine-N-oxide (TMAO, (**d**) Total serum/plasma indoxyl sulfate (IS), (**e**) Free serum/plasma indoxyl sulfate (IS), (**f**) Urinary indoxyl sulfate (IS), (**g**) Total serum/plasma p-cresyl sulfate (p-CS), (**h**) Free serum/plasma p-cresyl sulfate (p-CS), (**i**) Urinary p-cresyl sulfate (p-CS). Funnel plots of publication bias for the effect of dietary fiber supplementation on on gut microbiota metabolites in animal models with Chronic Kidney Disease (CKD) including: (**j**) Total serum/plasma indoxyl sulfate (IS), (**k**) serum/plasma p-cresyl sulfate (pCS), (**l**)serum/plasma acetate, (**m**) cecal acetate, (**n**) serum/plasma butyrate, (**o**) cecal butyrate, (**p**) serum/plasma propionate, **(q**) cecal propionate.

**Table 5b.**
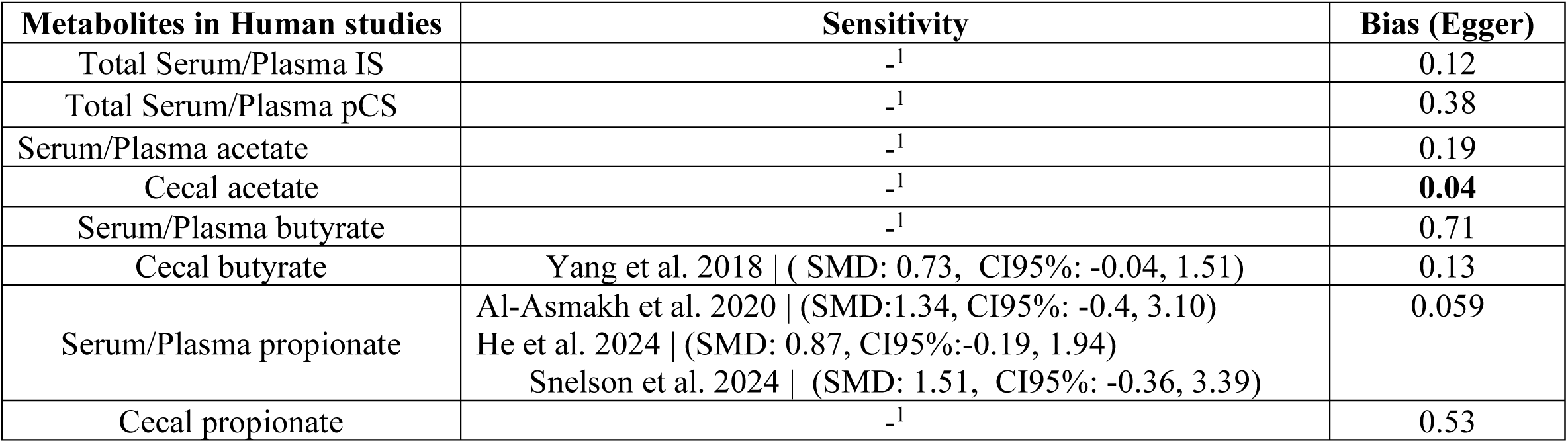
Bias sensitivity analysis and publication bias in animal models 1. Not detected 2. Not enough number of studies to conduct sensitivity analysis or Egger’s test

| Metabolites in Human studies | Sensitivity | Bias (Egger) |
| --- | --- | --- |
| Total Serum/Plasma IS | <sup>-1</sup> | 0.12 |
| Total Serum/Plasma pCS | <sup>-1</sup> | 0.38 |
| Serum/Plasma acetate | <sup>-1</sup> | 0.19 |
| Cecal acetate | <sup>-1</sup> | **0.04** |
| Serum/Plasma butyrate | <sup>-1</sup> | 0.71 |
| Cecal butyrate | Yang et al. 2018 \| ( SMD: 0.73, CI95%: -0.04, 1.51) | 0.13 |
| Serum/Plasma propionate | Al-Asmakh et al. 2020 \| (SMD:1.34, CI95%: -0.4, 3.10)<br>He et al. 2024 \| (SMD: 0.87, CI95%:-0.19, 1.94)<br>Snelson et al. 2024 \| (SMD: 1.51, CI95%: -0.36, 3.39) | 0.059 |
| Cecal propionate | <sup>-1</sup> | 0.53 |

**Figure 6.**
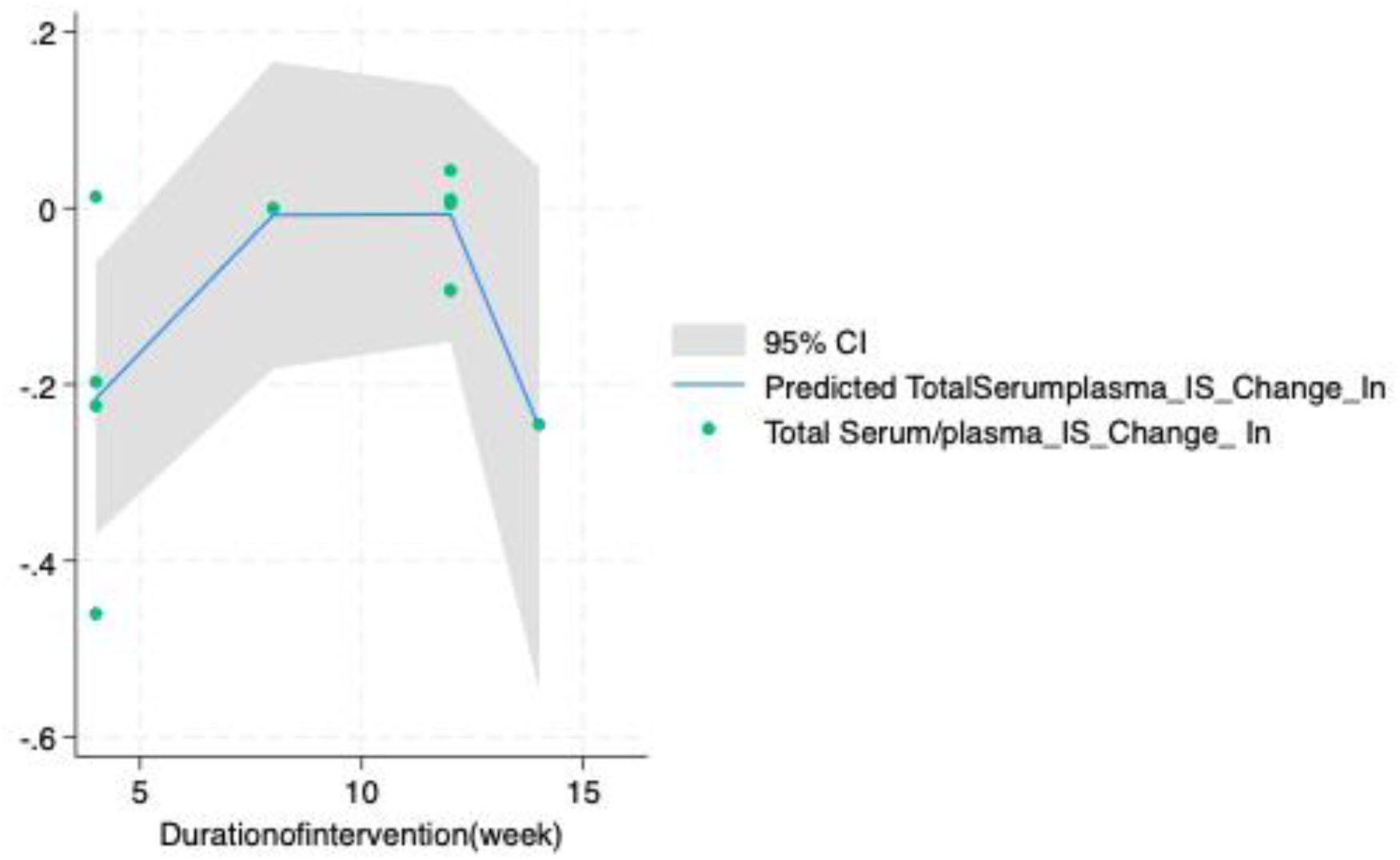
Dose-response analysis of the effect of fiber supplementation based on the duration on levels of pCS.

### Meta-regression and dose-response analysis

After conducting meta-regression and dose-response analysis of the effect of dietary fiber on total serum/plasma IS and p-CS, no significant results were shown suggesting that there is not an optimal dose to observe the best result. **(Table 6a and Figures S3a-d)**. Unlike the dietary fiber dose, duration of the intervention showed a nonlinear dose response association where the optimal reduction in total serum/plasma IS was 4 weeks **(Table 6a and Figure S4a)**. However, there was no non-linear relation between the duration of dietary fiber on total serum/plasma pCS.

**Table 6a.** Meta regression and dose-response analysis in RCTs

|  | Regression |  |  |  | Dose-response |  |  |  |
| --- | --- | --- | --- | --- | --- | --- | --- | --- |
|  | Dose |  | Duration |  | Dose |  | Duration |  |
|  | Coef | p | Coef | p | Coef | p | Coef | p |
| Total Serum/plasma IS | -0.95 | 0.34 | 0.08 | 0.87 | 17.42 | 0.21 | 0.64 | 0.44 |

**Table 6b.** Meta regression and dose-response analysis in animal models

|  | Regression |  |  |  | Dose-response |  |  |  |
| --- | --- | --- | --- | --- | --- | --- | --- | --- |
|  | Dose |  | Duration |  | Dose |  | Duration |  |
|  | Coef | p | Coef | p | Coef | p | Coef | p |
| Total Serum/plasma IS | 3.43 | 0.61 | 1.74 | 0.75 | 0.23 | 0.36 | 0.34 | **0.03** |
| Total Serum/Plasma pCS | -0.76 | 0.76 | -0.40 | 0.84 | -0.52 | 0.32 | 1.08 | 0.56 |

### GRADE assessment

The GRADE assessment indicated that the overall quality of evidence for the effect of dietary supplementation on gut microbiota metabolites in adults with CKD ranged from very low to moderate. The quality of evidence for total serum/plasma and free IAA and IS were rated as low, primarily due to serious inconsistency (moderate heterogeneity, I² >40%) and serious imprecision (insignificant results) similar to the quality of evidence for total serum/plasma TMAO primarily due to serious imprecision and low number of studies (n=2) and insignificant results. The quality of evidence for total plasma/serum and urinary IS and pCS and free serum/plasma pCS were rated as moderate, primarily due to serious inconsistency (moderate heterogeneity, I² >40%) (see **Table 7a).**

**Table 7a.**
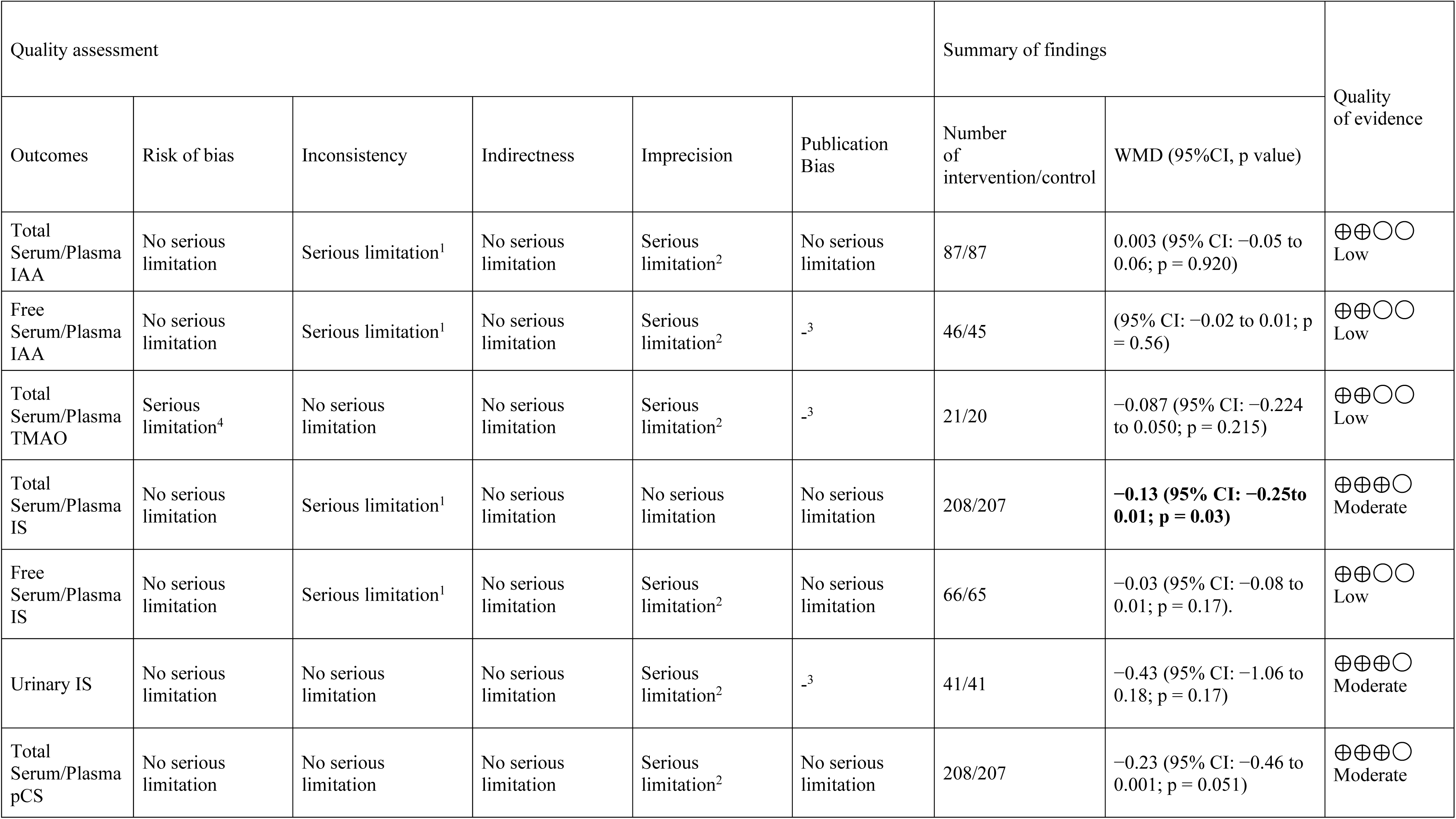
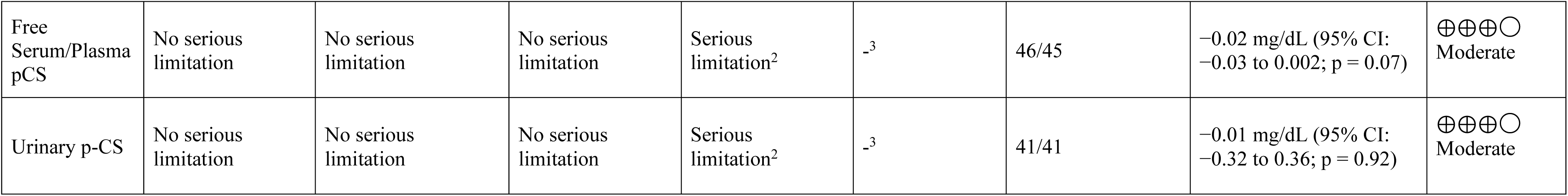
GRADE profile for the effect of dietary fiber supplementation on gut microbiota metabolites in adults with chronic kidney disease (CKD) Abbreviations: (IAA:) 1. There is moderate heterogeneity (I^2^ >40%) 2. There is no evidence of significant effects of dietary fiber supplementation on IAA, TMAO and free serum/plasma and urinary IS, total serum/plasma pCS levels 3. Not enough number of studies to conduct Egger’s test 4. Using the Cochrane Risk of Bias (RoB 2) assessment, one study (42) was classified as having some concerns, whereas the other (44) was assessed as high risk of bias

**Table 7b.** GRADE profile for the effect of dietary fiber supplementation on gut microbiota metabolites in animal models with chronic kidney disease (CKD) Abbreviations: (IAA:) 1. Studies with high risk of bias 2. There is moderate heterogeneity (I^2^ >40%) 3. There is no evidence of significant effects of dietary fiber supplementation on IAA level

| Quality assessment |  |  |  |  |  | Summary of findings |  | Quality of evidence |
| --- | --- | --- | --- | --- | --- | --- | --- | --- |
| Outcomes | Risk of bias | Inconsistency | Indirectness | Imprecision | Publication Bias | Number of intervention/control | SMD (95%CI, p value) |  |
| Total Serum/Plasma IS | Serious limitation<sup>1</sup> | Serious limitation<sup>2</sup> | No serious limitation | No serious limitation | Serious limitation | 126/107 | **−1.99 (95% CI: −3.06 to −0.92; p< 0.0001).** | ⊕○○○<br>Very Low |
| Total Serum/Plasma pCS | Serious limitation<sup>1</sup> | No serious limitation | No serious limitation | No serious limitation | No serious limitation | 38/37 | **−1.56 (95% CI: −2.08 to −1.03; p< 0.0001)** | ⊕⊕⊕○<br>Moderate |
| Serum/Plasma acetate | Serious limitation<sup>1</sup> | Serious limitation<sup>2</sup> | No serious limitation | Serious limitation<sup>2</sup> | No serious limitation | 40/36 | 1.14 (95% CI: −1.13 to 3.42; p = 0.32) | ⊕○○○<br>Very Low |
| Cecal acetate | Serious limitation<sup>1</sup> | Serious limitation<sup>2</sup> | No serious limitation | No serious limitation | Serious limitation | 31/23 | **2.00 (95% CI: 0.78 to 3.22; p = 0.001)** | ⊕○○○<br>Very Low |
| Serum/Plasma butyrate | Serious limitation<sup>1</sup> | Serious limitation<sup>2</sup> | No serious limitation | Serious limitation<sup>2</sup> | No serious limitation | 40/36 | 0.97 (95% CI: −0.47 to 2.41; p=0.18) | ⊕○○○<br>Very Low |
| Cecal butyrate | Serious limitation<sup>1</sup> | Serious limitation<sup>2</sup> | No serious limitation | Serious limitation<sup>2</sup> | No serious limitation | 31/23 | 0.73 (95% CI: -0.04 to 1.51; p = 0.06) | ⊕○○○<br>Very Low |
| Serum/Plasma propionate | Serious limitation<sup>1</sup> | Serious limitation<sup>2</sup> | No serious limitation | No serious limitation | No serious limitation | 40/36 | **1.51 (95% CI: 0.054 to 2.96; p=0.04)** | ⊕⊕⊕○<br>Moderate |
| Cecal propionate | Serious limitation<sup>1</sup> | Serious limitation<sup>2</sup> | No serious limitation | Serious limitation<sup>2</sup> | No serious limitation | 31/23 | 0.50 (95% CI: -0.93 to 1.93; p = 0.49) | ⊕○○○<br>Very Low |

## Meta-analysis of Animal Studies

### Effects of dietary fiber supplementation on total serum/plasma IS

A meta-analysis of 12 animal studies (54–62,65–67) four of which included two distinct intervention arms, yielding 16 separate comparisons, showed that dietary fiber supplementation, including xylo-oligosaccharides (XOS), resistant starch type 2, oligofructose-enriched inulin, inulin, gum acacia, psyllium seed husk (PSH), galacto-oligosaccharides, partially hydrolyzed guar gum, and β-glucan significantly reduced total serum or plasma IS concentrations. The pooled SMD was −1.99 (95% CI: −3.06 to −0.92; p< 0.0001). Substantial between-study heterogeneity was observed (I² = 87.6%, p< 0.0001) (**Figure 4j**).

### Subgroup Analysis of effects of dietary fiber supplementation on total serum/plasma IS

Results of subgroup analyses conducted for total serum/plasma IS are summarized in **Table 4b** by dietary supplementation type, fermentability, viscosity, and intervention dose. Subgroup analysis revealed that supplementation with dietary fiber significantly reduced total serum/plasma IS levels in animal models of CKD based on fiber type, fructan (SMD: −0.34, 95% CI: −0.52, −0.15; p< 0.0001) in seven studies (54,55,59,61), gum (SMD: −0.91], 95% CI: −1.42, −0.15; p< 0.0001) in seven studies (56,57,60,62,65,66), and hemicellulose (SMD: −0.001, 95% CI: −0.001, −0.001; p=0.002) in two arms of a study (58) compared to resistant starch type 2, which was not statistically significant (p = 0.08). Subgroup analysis based on fermentability showed that fermentable fiber reduced the serum/plasma IS significantly (SMD: −0.60, 95% CI: −0.81, −0.39; p< 0.0001) in 14 studies (54–57,59,60,62,65–67) similar to non-fermentable fibers (SMD: −0.0001, 95% CI: −0.0001, −0.0001; p=0.002) in 2 arms of a study (58). Subgroup analysis based on viscosity revealed that non-viscous fiber reduced the serum/plasma IS significantly (SMD: −0.25, 95% CI: −0.40, −0.09; p=0.002) in 8 studies (54,55,59,61,65,67) unlike viscous fibers (p=0.72) in 9 other studies (56–58,60,62,66). Subgroup analyses by intervention dose and duration did not materially alter the pooled estimates and yielded consistent results. (**Figures S1i-m**).

### Effects of dietary fiber supplementation on total serum/plasma pCS

A meta-analysis of 3 animal studies (54,61,67) one of which included two distinct intervention arms, yielding four separate comparisons, showed that dietary fiber supplementation, including resistant starch type 2, oligofructose-enriched inulin, and inulin reduced total serum/plasma pCS concentrations significantly. The pooled SMD was −1.56 (95% CI: −2.08 to −1.03; p< 0.0001). Low between-study heterogeneity was observed (I² = 40.0%, p=0.66) (**Figure 4k**). Due to limited numbers of studies, subgroup analysis was not conducted.

### Short chain fatty acids (SCFAs)

#### Effects of dietary fiber supplementation on serum/plasma acetate

A meta-analysis of four studies (55,64,67,68) one of which included two distinct intervention arms, yielding five separate comparisons, found that supplementation with dietary fiber, including XOS, gum acacia, resistant starch type 2 and inulin did not significantly affect serum/plasma acetate concentrations. The pooled SMD was 1.14 (95% CI: −1.13 to 3.42; p = 0.32), with high evidence of between-study heterogeneity (I² = 97.3%, p< 0.0001) (**Figure 4l**). Due to limited numbers of studies, subgroup analysis was not conducted (**Table 4b**).

### Effects of dietary fiber supplementation on cecal acetate

A meta-analysis of three studies (55,59,63) two of which included two distinct intervention arms, yielding five separate comparisons, found that supplementation with dietary fiber, including XOS, galacto-oligosaccharide, unmodified guar gum and partially hydrolyzed guar gum increased cecal acetate concentrations significantly. The pooled SMD was 2.00 (95% CI: 0.78 to 3.22; p = 0.001), with high evidence of between-study heterogeneity (I² = 63.5%, p=0.02) (**Figure 4m**). Due to limited numbers of studies, subgroup analysis was just conducted on fiber type which does not show any stratification between fiber types.

### Effects of dietary fiber supplementation on serum/plasma butyrate

A meta-analysis of four animal studies (55,64,67,68) one of which included two distinct intervention arms, yielding five separate comparisons, including XOS, gum acacia, resistant starch type 2 and inulin did not significantly affect serum/plasma butyrate concentrations. The pooled SMD was 0.97 (95% CI: −0.47 to 2,41; p=0.18). High between-study heterogeneity was observed (I² = 84.8%, p< 0.0001) (**Figure 4n**). Due to limited numbers of studies, subgroup analysis was not conducted.

### Effects of dietary fiber supplementation on cecal butyrate

A meta-analysis of three studies (55,59,63) two of which included two distinct intervention arms, yielding five separate comparisons, found that supplementation with dietary fiber, including XOS, galacto-oligosaccharides, unmodified guar gum and partially hydrolyzed guar gum did not change cecal butyrate concentrations significantly. The pooled SMD was 0.73 (95% CI: -0.04 to 1.51; p = 0.06), with low evidence of between-study heterogeneity (I² = 41.2%, p=0.14) (**Figure 4o**). Due to limited numbers of studies, subgroup analysis was not conducted.

### Effects of dietary fiber supplementation on serum/plasma propionate

A meta-analysis of four animal studies (55,64,67,68) one of which included two distinct intervention arms, yielding five separate comparisons, including XOS, gum acacia, resistant starch type 2 and inulin increased serum/plasma propionate concentrations significantly. The pooled SMD was 1.51 mg/dL (95% CI: 0.054 to 2,96; p=0.04). High between-study heterogeneity was observed (I² = 83.6%, p< 0.0001) (**Figure 4p**). Due to limited numbers of studies, subgroup analysis was not conducted.

### Effects of dietary fiber supplementation on cecal propionate

A meta-analysis of three studies (55,59,63) two of which included two distinct intervention arms, yielding five separate comparisons, found that supplementation with dietary fiber, including XOS, galacto-oligosaccharides, unmodified guar gum and partially hydrolyzed guar gum did not change cecal propionate concentrations significantly. The pooled SMD was 0.50 mg/dL (95% CI: -0.93 to 1.93; p = 0.49), with substantial evidence of between-study heterogeneity (I² = 79.9%, p=0.001) (**Figure 4q**). Due to limited numbers of studies, subgroup analysis was not conducted.

### Sensitivity analysis

Bias and sensitivity analysis results are summarized in **Table 5b and figures S2j-q.** Sensitivity analyses indicated that the pooled estimate for cecal butyrate was influenced by individual studies, including Yang et al. (55) (SMD: 1.00, CI95%: 0.32, 1,67) suggesting potential instability of the overall effect. Sensitivity analysis for serum/plasma propionate indicated that the studies by Al-Asmakh et al (64) (SMD: 0.73, CI95%: -0.4, 3.10), He et al. (68) (SMD: 0.87, CI95%: -0.19, 1.94), Snelson et al. (67) (SMD: 1.51, CI95%: -0.36, 3.39) impacted the overall results significantly indicating a potential influence on serum/plasma pCS outcomes (Table 5b).

### Publication bias

No evidence of publication bias was observed for total serum/plasma IS, pCS, acetate, butyrate, propionate and cecal butyrate **(Table 5b)**. Publication bias was detected for cecal acetate (p=0.04). Visual inspection of funnel plots suggested relative symmetry across all outcomes, indicating a low likelihood of small-study effects or selective reporting except for total serum/plasma IS, cecal acetate and propionate. Collectively, these findings support the robustness of the pooled estimates (**Fig 5j-q**).

### Meta-regression and dose-response analysis

After conducting meta-regression and dose-response analysis of the effect of dietary fiber on total serum/plasma IS in animal models of CKD, no significant results were shown suggesting that there is not an optimum dose to observe the best result in current literareture. **(Table 6b and Figures S3e-h)**.

### GRADE assessment

The GRADE assessment indicated that the overall quality of evidence for the effect of dietary supplementation on gut microbiota metabolites in animal models of CKD ranged from very low to moderate. The quality of evidence for total serum/plasma IS, acetate, butyrate, cecal butyrate and propionate were rated as very low, primarily due to serious inconsistency (moderate heterogeneity, I² >40%) and serious imprecision due low number of studies or insignificant results. The quality of evidence for plasma/serum propionate and total serum/plasma pCS was rated as moderate, primarily due to serious inconsistency (moderate heterogeneity, I² >40%) (see **Table 7b**).

## Discussion

### Aim and Main Findings

This systematic review and meta-analysis evaluated the effects of isolated dietary fibers based on viscosity and fermentability on gut microbiota metabolites in human and animal models with CKD. Across 13 RCTs and 15 animal studies, dietary fiber reduced cirulating IS and pCS in both human and animal models of CKD. In animal studies, dietary fiber increased cecal acetate and circulating propionate. There were no significant effects on ILA, IPA, IAA, TMAO, or butyrate, which may reflect the low number of studies. The certainty of evidence ranged from very low to moderate because of small sample sizes, risk of bias and study variability. These findings are clinically relevant because IS and pCS are protein-bound, gut-derived uremic toxins that accumulate as kidney function declines and have been implicated in CKD progression and end-organ damage, including inflammation, oxidative stress, endothelial dysfunction, cardiovascular injury, and renal fibrosis (74,75). Therefore, reductions in these metabolites may reflect not only a microbiome-related response to dietary fiber, but also a potentially meaningful strategy to reduce the uremic toxin burden that contributes to systemic complications of CKD.

Subgroup analyses showed that resistant starch type 2 reduced circulating IS in individuals with CKD, and fructans and gums reduced circulating IS and increased cecal acetate in animal models of CKD. Fermentable and non-viscous fiber reduced circulating IS in animal models. The fiber types evaluated across human and animal studies differ in their expected fermentation kinetics. Fructans, such as inulin and oligofructose, are generally considered highly fermentable and relatively rapidly fermented, whereas resistant starches, including resistant starch type 2, are fermented more gradually and may support fermentation in more distal colonic regions. Gums, pectin, and partially hydrolyzed guar gum are fermentable fibers, but their viscosity, solubility, and chemical structure can influence both the rate and site of fermentation. Psyllium, in contrast, is viscous but minimally fermented, and cellulose is largely non-fermentable. These differences are important because *in vitro* fermentation studies suggest that fibers can produce distinct SCFA profiles and fermentation rates over time, meaning that the metabolic effects of fiber in CKD may depend not only on whether a fiber is fermentable, but also on how quickly and where fermentation occurs in the gut (52). The subgroup analysis of participants on dialysis showed significant reductions in IS. There were also trends toward lower circulating IS in human studies with intervention duration greater than 8 weeks, and dose below 10 g per day. However, the apparent dose pattern should be interpreted with caution because supplemental doses are strongly influenced by fiber type and tolerability. For example, rapidly fermentable fructans are often administered at lower doses due to gastrointestinal tolerance, whereas resistant starches are commonly provided at higher doses because they are more slowly fermented and less osmotically active in the proximal colon. Therefore, a lower effective dose may reflect the physicochemical and fermentative properties of the fiber rather than a true advantage of lower-dose interventions (70–72). Subgroup analyses were not feasible for several outcomes because of the limited number of studies. Dose-response analysis showed a significant nonlinear association between intervention duration and changes in circulating IS in human studies, with no significant linear or non-linear associations for dose or duration for other metabolites in human or animal studies. A summary of all the results is included in **Table 8**.

**Table 8.**
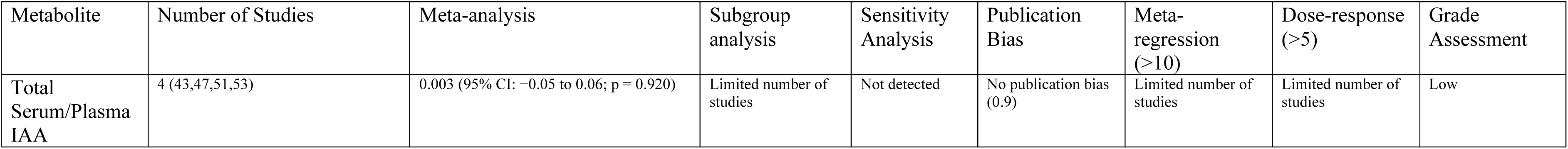
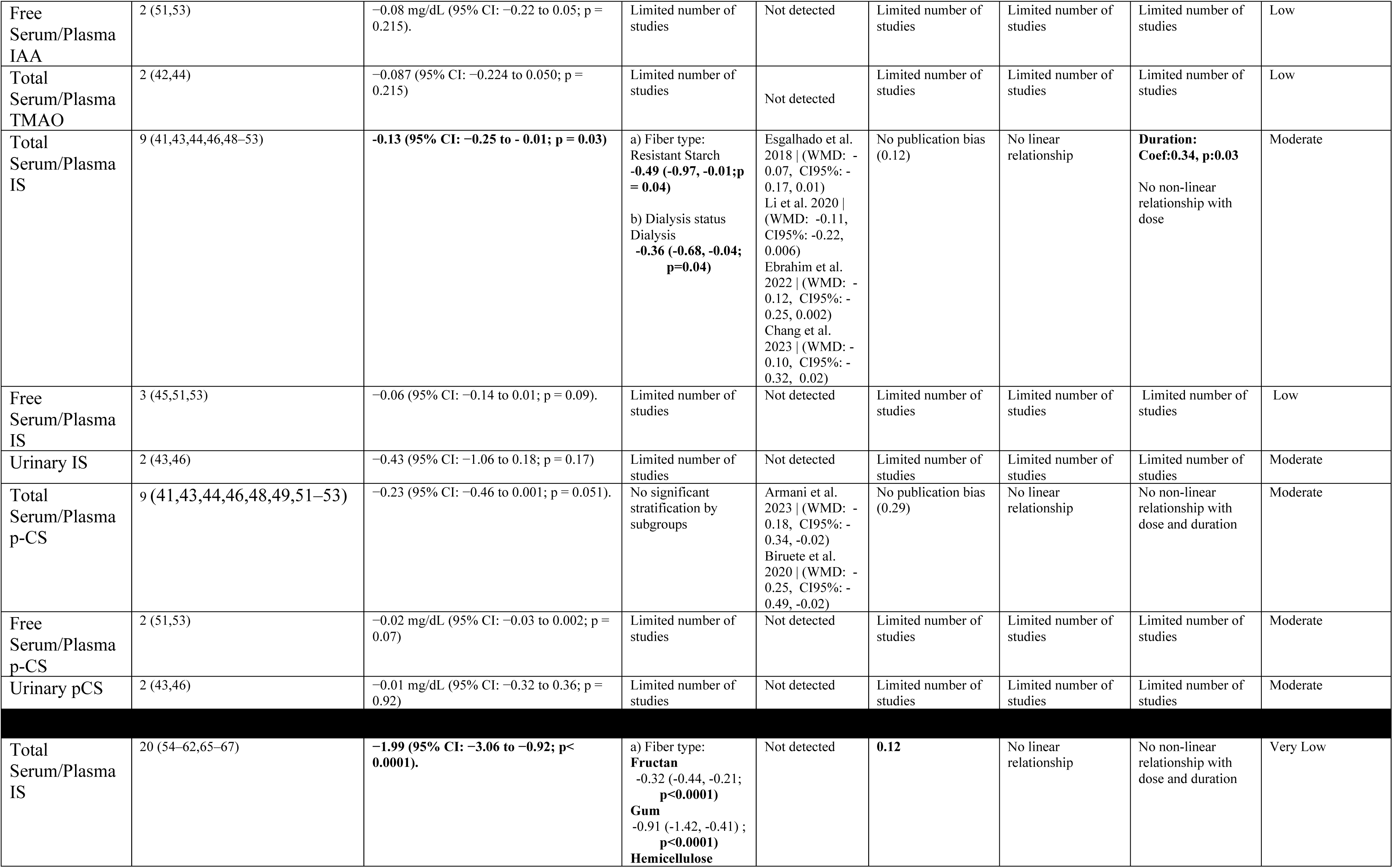
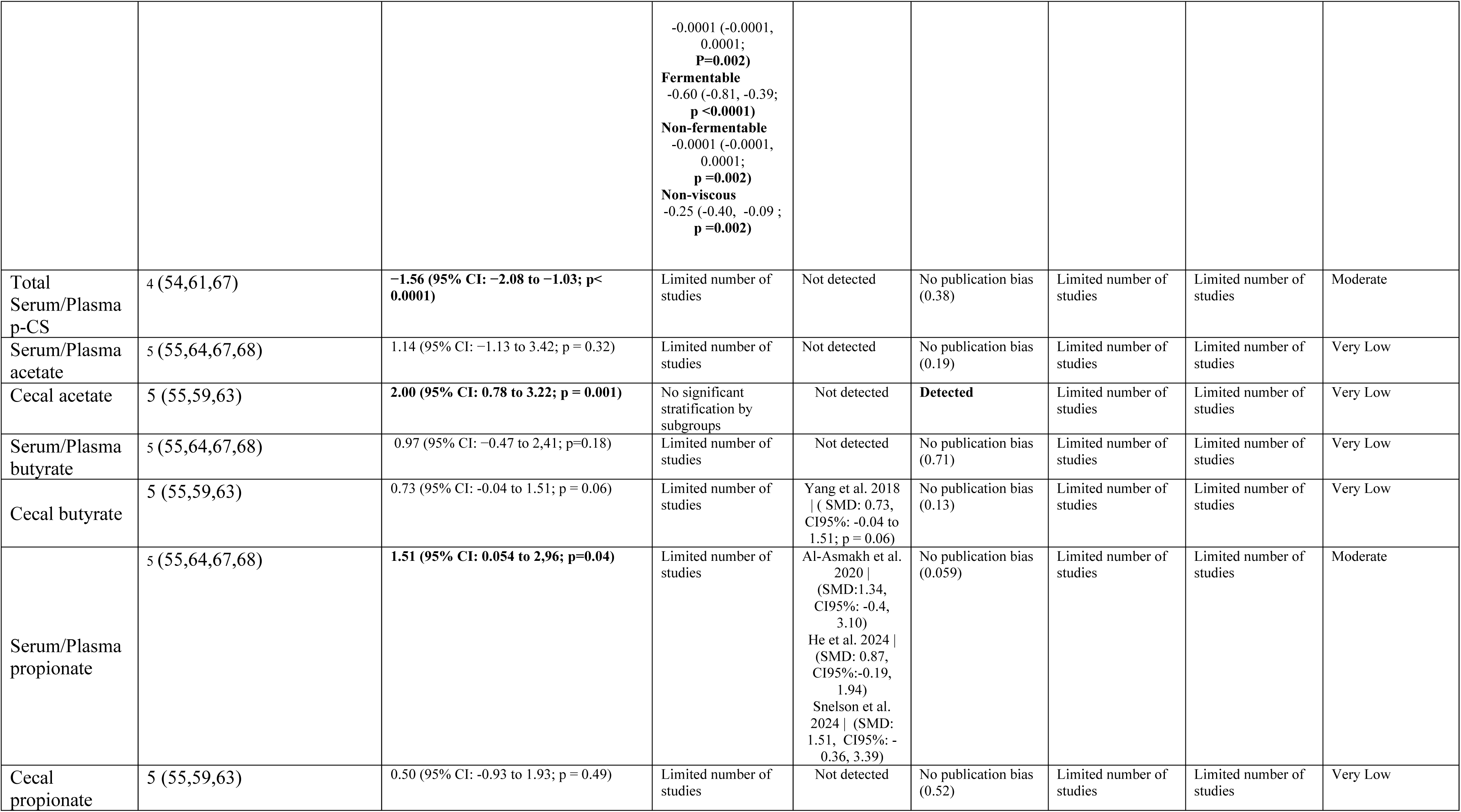
Summary table of all measurement and outcomes for RCTs and animal studies. The significant results are in bold.

Our results are broadly aligned with the existing body of evidence demonstrating that isolated dietary fiber can modulate gut-derived uremic toxins in CKD. A comprehensive systematic review and meta-analysis of 21 RCTs reported that fiber supplementation significantly lowered circulating IS and pCS, although no effect was observed for TMAO (73). Consistent with this, a network meta-analysis evaluating prebiotic, probiotic, and synbiotic interventions found significant reductions in IS and pCS, likely mediated through alterations in gut microbial composition (74). Evidence specific to resistant starch type 2 further supports these findings: a meta-analysis of 10 RCTs identified a significant decrease in IS but not pCS, whereas another review incorporating 5 RCTs for IS and 6 RCTs for pCS reported significant reductions in both metabolites following fiber interventions (75). Additionally, observational and experimental data reinforce the relevance of microbially derived metabolites in CKD pathophysiology; Wang et al. (76) demonstrated inverse associations between SCFAs and CKD, consistent with findings from Zhong et al. (77). Together, these studies corroborate our results and highlight the mechanistic importance of fiber-microbiota interactions in mitigating uremic toxin burden in CKD. Importantly, most human evidence has focused on fermentable fibers, particularly resistant starch 2 and fructans, whereas less is known about poorly fermentable or non-fermentable fibers in patients with CKD. This limits the ability to distinguish whether reductions in uremic toxins are driven primarily by fermentation-dependent mechanisms, such as increased SCFA production and reduced proteolytic fermentation, or by fermentation-independent mechanisms, such as improved bowel regularity, faster colonic transit, and reduced contact time between luminal protein substrates and proteolytic bacteria.

### Possible Underlying Mechanisms

Findings from this systematic review and meta-analysis demonstrate that dietary fiber supplementation consistently reduces circulating concentrations of the gut-derived uremic solutes IS and pCS in patients and animal models with CKD, with more variable effects observed for TMAO and other microbial metabolites. These results are biologically plausible within the framework of the gut-kidney axis. CKD is characterized by increased proteolytic fermentation, impaired intestinal barrier integrity, and reduced renal clearance of protein-bound solutes, all of which contribute to toxin accumulation (78). The observed reductions in IS and pCS likely reflect a shift from proteolytic to saccharolytic metabolism when fermentable fiber is available, limiting formation of indole and p-cresol precursors and decreasing downstream sulfated toxin production. This shift may be especially relevant for rapidly or highly fermentable fibers, such as fructans and some gums, which can provide readily available carbohydrate substrates for saccharolytic bacteria. More slowly fermented fibers, such as resistant starch type 2, may have additional relevance because they can escape fermentation in the proximal colon and provide substrate more distally, where proteolytic fermentation is more likely to occur when carbohydrate availability is limited. In this way, slow fermentation may help suppress protein fermentation across a broader region of the colon (70). Improved epithelial barrier function through SCFA-mediated tightening of junctional complexes may further restrict translocation of microbial metabolites into circulation (22). For fibers that are poorly fermented, such as psyllium, or largely non-fermentable, such as cellulose, potential benefits may occur through a different pathway. By increasing stool bulk, water-holding capacity, and intestinal transit, these fibers may reduce the time available for microbial proteolysis and decrease contact between amino acid substrates and bacteria capable of producing indole and p-cresol. This mechanism may be particularly important in CKD, where constipation and prolonged colonic transit are common and may contribute to greater generation and absorption of gut-derived uremic toxins. We observed a significant increase in cecal acetate and serum/plasma propionate, and a trend towards increased cecal butyrate in animal studies, providing mechanistic support for these effects. Acetate, propionate, and butyrate generated through microbial fermentation of fiber, act not only as substrates for colonocyte energy metabolism but also as signaling molecules activating G protein-coupled receptors (GPR41, GPR43, GPR109A, OLFR78, OLFR558) expressed across immune, vascular, and renal tissues (79). Activation of these receptors modulates inflammatory pathways, oxidative stress, and vascular tone, and has been linked to improved blood pressure regulation and renal homeostasis (80). Thus, the reduction in uremic solutes seen across studies may be mediated both by decreased precursor availability and by SCFA-driven improvements in intestinal integrity and systemic inflammatory status (6). Collectively, these mechanisms suggest that dietary fiber may lower gut-derived uremic toxins through both fermentation-dependent and fermentation-independent pathways. Fermentable fibers may reduce toxin generation by increasing saccharolytic fermentation and SCFA production, whereas viscous or poorly fermentable fibers may reduce toxin generation by accelerating transit and limiting the duration of protein fermentation. However, because human studies have included relatively few non-fermentable or poorly fermentable fiber interventions, future trials are needed to determine whether these transit-related mechanisms independently reduce IS and pCS in CKD.

### Implications for Clinical Practice

This review indicates that increasing fermentable dietary fiber may be a practical adjunct for reducing gut-derived uremic solutes in CKD. However, current clinical guidelines offer limited direction regarding optimal fiber intake for this clinical population. In practice, dietary counseling for CKD often prioritizes restriction of phosphorus, potassium, sodium and modulation of protein depending on the CKD stage (81). This approach can inadvertently reduce fiber intake because many high-fiber plant foods are presumed to increase mineral-related risks (82). Importantly, phosphorus in vegetables, legumes, and whole grains is largely phytate-bound and substantially less absorbable than phosphorus from animal sources or additives (83). Consequently, patients may be limiting foods that could beneficially influence the gut-kidney axis. While the nutrition guidance has evolved in the last decade, studies consistenty report patients with CKD having a low fiber intake (84,85). Our findings highlight the clinical need to communicate that, when appropriately selected and individualized selected dietary fiber supplements can be safely integrated into CKD diets and may contribute to lowering lower uremic toxin burden. The subgroup findings also highlight important considerations for translating evidence into practice. Differences in fiber supplement dose (<16 vs ≥16 g/day) did not yield statistically distinct effects, likely because supplemental fiber does not reflect total fiber intake, which varied substantially across studies and was not consistently reported. In addition, dose ranges differed by fiber type, with isolated fermentable fibers such as inulin-type fructans typically administered at lower doses than less fermentable fibers such as resistant starch, limiting direct dose-based comparisons across interventions. This suggests that clinical recommendations should account for habitual intake rather than focusing solely on the added dose, as well as the fiber type. Intervention duration showed a non-significant but consistent trend toward greater reductions in IS with longer supplementation, supporting the concept that meaningful modulation of gut microbial metabolism requires sustained exposure. Larger reductions among dialysis patients may reflect their higher baseline toxin concentrations and the limited dialyzability of protein-bound solutes such as IS and PCS, making them more responsive to interventions that reduce precursor generation in the colon.

These findings collectively point to a gap between current dietary guidance and emerging evidence on gut microbiota-derived metabolites. Integrating fiber type and quantity into routine CKD nutrition counseling, as well as clarifying the differential bioavailability of plant versus additive-derived phosphorus may improve patient understanding and reduce unnecessary avoidance of beneficial plant-based foods. Future clinical trials should report both baseline and achieved total fiber intake to enable more precise dose-response analyses and inform evidence-based targets that can be incorporated into evolving CKD nutrition guidelines.

### Study strengths and Limitations

This systematic review and meta-analysis have several notable strengths. To our knowledge, this is the first synthesis to jointly evaluate both human and animal evidence on isolated dietary fiber interventions and gut-derived metabolites in CKD, enabling a more comprehensive assessment of mechanistic plausibility and translational relevance. We also focused specifically on isolated dietary fibers, reducing confounding from whole-diet interventions. Another strength is the inclusion of an expanded metabolite profile IS, PCS, TMAO, multiple tryptophan metabolites as well as SCFAs such as acetate, propionate and butyrate which provides a more complete characterization of the gut-kidney axis. Additionally, this study is among the first to incorporate fiber-type subgroup analyses based on fermentability and viscosity, allowing examination of physiologically meaningful differences across fiber classes. The separate evaluation of dosing and intervention duration further strengthens the interpretability of the findings.

However, several limitations should be acknowledged. For several metabolites, particularly IPA, IAA, ILA, and TMAO, the number of available studies was small, limiting statistical power and precluding subgroups that would have been clinically informative. The overall certainty of evidence ranged from very-low to moderate, reflecting methodological limitations in primary studies, including incomplete reporting of baseline fiber intake, heterogeneous intervention durations, and variable analytical techniques for metabolite quantification. Evidence of between-study heterogeneity was observed for some outcomes, and small-study effects could not be excluded, suggesting possible publication bias. Finally, most interventions were short-term, and the long-term sustainability, tolerability, and real-world feasibility of increasing fiber intake in CKD remain insufficiently studied. However, while it is clear that most of the studies included fermentable fibers, there is an area of opportunity for exploring viscous fibers.

Overall, while the evidence suggests biologically meaningful reductions in gut-derived toxins with fermentable fiber, larger and longer-duration trials with standardized reporting of total fiber intake, kidney function, and clinical endpoints are needed to support integration of fiber-specific recommendations into CKD dietary guidelines.

### Conclusion

In summary, this systematic review and meta-analysis demonstrates that isolated dietary fiber supplementation reduces key gut-derived uremic solutes, particularly IS and pCS, in both human and animal models of CKD, with limited evidence for increased SCFA production as a potential mechanistic pathway in animal studies. These findings reinforce the central role of the gut-kidney axis in CKD and suggest that fermentable and non-viscous fibers may modulate microbial metabolism and attenuate protein-bound toxins. Although subgroup analyses indicated that fiber characteristics, total intake, and intervention duration may influence the magnitude of response, the available evidence remains limited for several metabolites and constrained by heterogeneity in study design and quality. Taken together, the results point to dietary fiber as a promising, low-cost, and biologically plausible adjunctive strategy for reducing gut-derived toxin burden in CKD, but the current evidence base is insufficient to guide specific clinical recommendations. Well-designed, longer-term trials with standardized reporting of total fiber intake, fiber physicochemical properties, and clinically relevant outcomes are needed to clarify dose–response relationships and determine whether improvements in microbial metabolites translate into measurable benefits in kidney function, cardiovascular risk, and patient-centered outcomes.

## Abbreviations

CKD: chronic kidney disease
CKD-MBD: chronic kidney disease–mineral and bone disorder
FOS: fructo-oligosaccharide
FMO3: flavin-containing monooxygenase 3
GFR: glomerular filtration rate
IAA: indole-3-acetic acid
IAld: indole-3-aldehyde
ILA: indole-3-lactic acid
IPA: indole-3-propionic acid
IS: indoxyl sulfate
pCG: *p*-cresyl glucuronide
pCS: *p*-cresyl sulfate
RCT: randomized clinical trial
SCFAs: short-chain fatty acids
SMD: standardized mean difference
SYRCLE: systematic review centre for laboratory animal experimentation
TMA: trimethylamine
TMAO: trimethylamine *N*-oxide
WMD: weighted mean difference

## Supplementary Materials

The following supporting information can be downloaded at (). Search strategy in all databases, all codes in STATA SE., Figure S1: Forest plots of subgroup analysis for both RCTs and animal models; Figure S2: Forest plots of sensitivity analysis both RCTs and animal models. Figure S3: Meta-regression graph of linear association between dietary fiber and IS based on dose and duration of intervention.

## Author Contributions

Conceptualization: SNM, AB; Methodology: SMN, AB, and JR; Formal Analysis: SNM; Investigation: SNM, CC, HEW, AB and BH.; Writing—Original Draft Preparation: SNM.; Writing—Review and editing: SNM, AB, SM, BK, CC, JR; Project Administration: SNM and AB. All authors have read and agreed to the published version of the manuscript.

## Data availability

All data used in this meta-analysis were extracted from published studies. The datasets supporting the findings of this study are available from the original sources cited in the manuscript. Additional information can be provided by the corresponding author upon reasonable request.

