## Supplemental files for "“The effect of dietary fiber based on fermentability and viscosity on the gut microbial metabolites in chronic kidney disease: a systematic review and meta-analysis of experimental and clinical trials.”"

**Supplementary Materials**

Systematic review search terms in all databases for both RCTs and animal studies.

**PubMed**

1. "Dietary Fiber"[Mesh] OR "Cellulose"[Mesh] OR "Polysaccharides"[Mesh] OR "Resistant Starch"[Mesh] OR "Chicory"[Mesh] OR "Dextrins"[Mesh] OR "Pectins"[Mesh] OR "Psyllium"[Mesh] OR "locust bean gum"[Supplementary Concept] OR "Alginates"[Mesh] OR "high-amylose maize type 2 resistant starch, maize"[Supplementary Concept] OR "polydextrose"[Supplementary Concept] OR "4'-galactooligosaccharide"[Supplementary Concept] OR "1-6-alpha-glucomannan"[Supplementary Concept] OR "Gum Arabic"[Mesh]
2. "Dietary fiber"[Title/Abstract] OR "fiber*"[Title/Abstract] OR "dietary fibre"[Title/Abstract] OR "cellulose"[Title/Abstract] OR "hemicellulose*"[Title/Abstract] OR "Resistant maltodextrin"[Title/Abstract] OR "resistant starch"[Title/Abstract] OR "galactooligosaccharides"[Title/Abstract] OR "dextrin"[Title/Abstract] OR "Pectin"[Title/Abstract] OR "beta-glucan"[Title/Abstract] OR "partially-hydrolyzed guar gum"[Title/Abstract] OR "PHGG"[Title/Abstract] OR "methylcellulose"[Title/Abstract] OR "fruit"[Title/Abstract] OR "vegetable"[Title/Abstract] OR "polysaccharide*"[Title/Abstract] OR "psyllium*"[Title/Abstract] OR "ispagula*"[Title/Abstract] OR "metamucil*"[Title/Abstract] OR "polymer*"[Title/Abstract] OR "carbohydrate*"[Title/Abstract] OR "dietary carbohydrate*"[Title/Abstract] OR "fructan*"[Title/Abstract] OR "chicory"[Title/Abstract] OR "fructooligosaccharides"[Title/Abstract] OR "arabinoxylan*"[Title/Abstract] OR "asteraceae"[Title/Abstract] OR "fructooligosaccharide*"[Title/Abstract] OR "oligofructose*"[Title/Abstract] OR "inulin"[Title/Abstract] OR "whole grain*"[Title/Abstract] OR "wholegrain"[Title/Abstract] OR "whole meal"[Title/Abstract] OR "whole wheat"[Title/Abstract] OR "edible grain"[Title/Abstract] OR "wheat"[Title/Abstract] OR "rice"[Title/Abstract] OR "brown rice"[Title/Abstract] OR "maize"[Title/Abstract] OR "oat"[Title/Abstract] OR "barley"[Title/Abstract] OR "corn"[Title/Abstract] OR "rye"[Title/Abstract] OR "millet"[Title/Abstract] OR "sorghum"[Title/Abstract] OR "Beta-glucan soluble fiber"[Title/Abstract] OR "Psyllium husk"[Title/Abstract] OR "locust bean gum"[Title/Abstract] OR "Hydroxypropylmethylcellulose"[Title/Abstract] OR "Alginate"[Title/Abstract] OR "High amylose starch"[Title/Abstract] OR "Resistant maltodextrin"[Title/Abstract] OR "Resistant dextrin"[Title/Abstract] OR "resistant starch 2"[Title/Abstract] OR "Polydextrose"[Title/Abstract] OR "Galactooligosaccharide"[Title/Abstract] OR "Glucomannan"[Title/Abstract] OR "Acacia"[Title/Abstract] OR "gum arabic"[Title/Abstract] OR "inulin-type fructans"[Title/Abstract]
3. #1 OR #2

4. "Kidney Diseases"[Mesh] OR "Kidney Failure, Chronic"[Mesh] OR "Renal Insufficiency"[Mesh] OR "Acute Kidney Injury"[Mesh]
5. "Kidney"[Title/Abstract] OR "renal"[Title/Abstract]
6. "Disease"[Title/Abstract] OR "failure"[Title/Abstract] OR "function"[Title/Abstract] OR "insufficiency"[Title/Abstract] OR "dysfunction"[Title/Abstract]
7. #5 AND #6
8. #4 OR #7
9. "Gastrointestinal Microbiome"[Mesh] OR "Lactobacillus"[Mesh] OR "Bifidobacterium"[Mesh] OR "Bacteroides"[Mesh] OR "Akkermansia"[Mesh] OR "Faecalibacterium"[Mesh] OR "Prevotella"[Mesh] OR "Escherichia coli"[Mesh] OR "Enterococcus"[Mesh] OR "Fecal Microbiota Transplantation"[Mesh] OR "gut flora"[Title/Abstract] OR "Sequence Analysis"[Mesh] OR "RNA, Ribosomal, 16S"[Mesh] OR "RNA-Seq"[Mesh] OR "Denaturing Gradient Gel Electrophoresis"[Mesh]
10. "gut microbiota "[Title/Abstract] OR "gut microbiota composition"[Title/Abstract] OR "gut microbiome"[Title/Abstract] OR "gut bacteria"[Title/Abstract] OR Lactobacillus[Title/Abstract] OR Bifidobacterium[Title/Abstract] OR Bacteroides[Title/Abstract] OR Akkermansia[Title/Abstract] OR Ruminococcaceae[Title/Abstract] OR Prevotella[Title/Abstract] OR "Escherichia coli"[Title/Abstract] OR E. coli[Title/Abstract] OR Enterococcus[Title/Abstract] OR "gut bacteria"[Title/Abstract] OR "fecal microbiota"[Title/Abstract] OR "bacteriome"[Title/Abstract] OR "amplicon sequencing"[Title/Abstract] OR "16S rRNA"[Title/Abstract] OR "16S rRNA sequencing"[Title/Abstract] OR "shotgun metagenomics"[Title/Abstract] OR "shallow metagenomics"[Title/Abstract] OR "454 pyrosequencing"[Title/Abstract] OR "dgge "[Title/Abstract] OR "denaturating gradient gel electrophoresis"[Title/Abstract] OR "alpha-diversity"[Title/Abstract] OR "beta-diversity"[Title/Abstract]
11. "trimethyloxamine" [Supplementary Concept] OR "Fatty Acids, Volatile"[Mesh] OR "Cholic Acid"[Mesh] OR "Propionates"[Mesh] OR "Uremic Toxins"[Mesh]
12. "bile acids"[Title/Abstract] OR "short-chain fatty acids"[Title/Abstract] OR SCFA[Title/Abstract] OR "branched-chain amino acids"[Title/Abstract] OR "BCCA"[Title/Abstract] OR "trimethylamine n-oxide"[Title/Abstract] OR TMAO[Title/Abstract] OR "cholic acid"[Title/Abstract] OR cholate[Title/Abstract] OR "chenodeoxycholic acid"[Title/Abstract] OR chenodeoxycholate[Title/Abstract] OR "taurocholic acid"[Title/Abstract] OR taurocholate[Title/Abstract] OR "glycocholic acid"[Title/Abstract] OR glycocholate[Title/Abstract] OR "taurochenodeoxycholic acid"[Title/Abstract] OR taurochenodeoxycholate[Title/Abstract] OR "glycochenodeoxycholic acid"[Title/Abstract] OR glycochenodeoxycholate[Title/Abstract] OR "glycocholenate sulfate"[Title/Abstract] OR "glycohyocholate"[Title/Abstract] OR "glycolithocholate sulfate"[Title/Abstract] OR glycoursodeoxycholate[Title/Abstract] OR taurocholenate_sulfate[Title/Abstract] OR taurodeoxycholate[Title/Abstract] OR "taurolithocholate_3_sulfate"[Title/Abstract] OR tauroursodeoxycholate[Title/Abstract] OR "deoxycholic Acid"[Title/Abstract] OR "wheat

bran"[Title/Abstract] OR deoxycholate[Title/Abstract] OR "lithocholic acid"[Title/Abstract] OR lithocholate[Title/Abstract] OR "acetic acid"[Title/Abstract] OR acetate[Title/Abstract] OR "propionic acid"[Title/Abstract] OR propionate[Title/Abstract] OR "butyric Acid"[Title/Abstract] OR butyrate[Title/Abstract] OR leucine[Title/Abstract] OR isoleucine[Title/Abstract] OR valine[Title/Abstract] OR indole[Title/Abstract] OR tryptophan[Title/Abstract] OR indole-3-lactate[Title/Abstract] OR "indole-3-lactic acid"[Title/Abstract] OR indole-3-propionate[Title/Abstract] OR "indole-3-propionic acid"[Title/Abstract] OR indole-3-aldehyde[Title/Abstract] OR "indoxyl sulphate"[Title/Abstract] OR "IPA"[Title/Abstract] OR " indoxyl sulfate "[Title/Abstract] OR "p-cresyl sulphate"[Title/Abstract] OR "phenol sulfate"[Title/Abstract] OR "phenylacetylglutamine "[Title/Abstract] OR "indoxylsulfuric acid"[Title/Abstract] OR indole-3- acetate[Title/Abstract] OR "indole-3-acetic acid"[Title/Abstract] OR indole-3-carboxylate[Title/Abstract] OR "indole-3-carboxylic acid"[Title/Abstract] OR indole-acetaldehyde[Title/Abstract] OR "p-cresyl sulfate"[Title/Abstract] OR "gut microbiota metabolite*"[Title/Abstract] OR "gut metabolite*"[Title/Abstract] OR "gastrointestinal metabolite*"[Title/Abstract] OR "uremic toxin*"[Title/Abstract]

13. #9 OR #10 OR #11 OR #12
14. **14.** #3 AND #8 AND #13

**<u>Results: 3,139</u>**

**CINAHL**

1. (MH "Polysaccharides+") OR (MH "Dietary Fiber") OR (MH "Cellulose") OR (MH "Alginates") OR (MH "Psyllium") OR (MH "Resistant Starch") OR (MH "Fruit+") OR (MH "Vegetables+") OR (MH "Polymers+") OR (MH "Dietary Carbohydrates+") OR (MH "Inulin") OR (MH "Cereals+") OR (MH "Wheat") OR (MH "Rice") OR (MH "Corn") OR (MH "Oats") OR (MH "Barley")
2. TI ( "Dietary fiber" OR "fiber*" OR "dietary fibre" OR "cellulose" OR "hemicellulose*" OR "Resistant maltodextrin" OR "resistant starch" OR "galactooligosaccharides" OR "dextrin" OR "Pectin" OR "beta-glucan" OR "partially-hydrolyzed guar gum" OR "PHGG" OR "methylcellulose" OR "fruit" OR "vegetable" OR "polysaccharide*" OR "psyllium*" OR "ispagula*" OR "metamucil*" OR "polymer*" OR "carbohydrate*" OR "dietary carbohydrate*" OR "fructan*" OR "chicory" OR "fructooligosaccharides" OR "arabinoxylan*" OR "asteraceae" OR "fructooligosaccharide*" OR "oligofructose*" OR "inulin" OR "whole grain*" OR "wholegrain" OR "whole meal" OR "whole wheat" OR "edible grain" OR "wheat" OR "rice" OR "brown rice" OR "maize" OR "oat" OR "barley" OR "corn" OR "rye" OR "millet" OR "sorghum" OR "Beta-glucan soluble fiber" OR "Psyllium husk" OR "locust bean gum" OR "Hydroxypropylmethylcellulose" OR "Alginate" OR "High amylose starch" OR "Resistant maltodextrin" OR "Resistant dextrin" OR "resistant starch 2" OR "Polydextrose" OR "Galactooligosaccharide" OR "Glucomannan" OR "Acacia" OR "gum arabic" OR "inulin-type fructans" ) OR AB ( "Dietary fiber" OR "fiber*" OR "dietary fibre" OR "cellulose" OR "hemicellulose*" OR "Resistant maltodextrin" OR "resistant starch" OR "galactooligosaccharides" OR

"dextrin" OR "Pectin" OR "beta-glucan" OR "partially-hydrolyzed guar gum" OR "PHGG" OR "methylcellulose" OR "fruit" OR "vegetable" OR "polysaccharide*" OR "psyllium*" OR "ispagula*" OR "metamucil*" OR "polymer*" OR "carbohydrate*" OR "dietary carbohydrate*" OR "fructan*" OR "chicory" OR "fructooligosaccharides" OR "arabinoxylan*" OR "asteraceae" OR "fructooligosaccharide*" OR "oligofructose*" OR "inulin" OR "whole grain*" OR "wholegrain" OR "whole meal" OR "whole wheat" OR "edible grain" OR "wheat" OR "rice" OR "brown rice" OR "maize" OR "oat" OR "barley" OR "corn" OR "rye" OR "millet" OR "sorghum" OR "Beta-glucan soluble fiber" OR "Psyllium husk" OR "locust bean gum" OR "Hydroxypropylmethylcellulose" OR "Alginate" OR "High amylose starch" OR "Resistant maltodextrin" OR "Resistant dextrin" OR "resistant starch 2" OR "Polydextrose" OR "Galactooligosaccharide" OR "Glucomannan" OR "Acacia" OR "gum arabic" OR "inulin-type fructans" ) OR MW ( "Dietary fiber" OR "fiber*" OR "dietary fibre" OR "cellulose" OR "hemicellulose*" OR "Resistant maltodextrin" OR "resistant starch" OR "galactooligosaccharides" OR "dextrin" OR "Pectin" OR "beta-glucan" OR "partially-hydrolyzed guar gum" OR "PHGG" OR "methylcellulose" OR "fruit" OR "vegetable" OR "polysaccharide*" OR "psyllium*" OR "ispagula*" OR "metamucil*" OR "polymer*" OR "carbohydrate*" OR "dietary carbohydrate*" OR "fructan*" OR "chicory" OR "fructooligosaccharides" OR "arabinoxylan*" OR "asteraceae" OR "fructooligosaccharide*" OR "oligofructose*" OR "inulin" OR "whole grain*" OR "wholegrain" OR "whole meal" OR "whole wheat" OR "edible grain" OR "wheat" OR "rice" OR "brown rice" OR "maize" OR "oat" OR "barley" OR "corn" OR "rye" OR "millet" OR "sorghum" OR "Beta-glucan soluble fiber" OR "Psyllium husk" OR "locust bean gum" OR "Hydroxypropylmethylcellulose" OR "Alginate" OR "High amylose starch" OR "Resistant maltodextrin" OR "Resistant dextrin" OR "resistant starch 2" OR "Polydextrose" OR "Galactooligosaccharide" OR "Glucomannan" OR "Acacia" OR "gum arabic" OR "inulin-type fructans" )

3. #1 OR #2
4. (MH "Kidney Diseases+") OR (MH "Kidney Failure, Chronic+") OR (MH "Renal Insufficiency+") OR (MH "Renal Insufficiency, Chronic+") OR (MH "Kidney Failure, Acute+")
5. TI ( "Kidney" OR "renal" ) OR AB ( "Kidney" OR "renal" ) OR MW ( "Kidney" OR "renal" )
6. TI ( "Disease" OR "failure" OR "function" OR "insufficiency" OR "dysfunction" ) OR AB ( "Disease" OR "failure" OR "function" OR "insufficiency" OR "dysfunction" ) OR MW ( "Disease" OR "failure" OR "function" OR "insufficiency" OR "dysfunction" )
7. #5 AND #6
8. #4 OR #7
9. (MH "Gut Microbiota") OR (MH "Microbiota+") OR (MH "Bifidobacterium") OR (MH "Faecalibacterium") OR (MH "Escherichia Coli") OR (MH "Enterococcus+") OR (MH "Fecal Microbiota Transplantation") OR
   (MH "Sequence Analysis+") OR (MH "Fatty Acids+") OR (MH "Fatty Acids, Essential+") OR (MH "Propionates+") OR (MH "Uremic Toxins") OR MH "Uremic

Toxins") OR (MH "Bile Acids and Salts") OR (MH "Amino Acids, Branched-Chain+") OR (MH "Leucine")

10. TI ( "gut microbiota " OR "gut microbiota composition" OR "gut microbiome" OR "gut bacteria" OR Lactobacillus OR Bifidobacterium OR Bacteroides OR Akkermansia OR Ruminococcaceae OR Prevotella OR "Escherichia coli" OR E. coli OR Enterococcus  OR "gut bacteria" OR "fecal microbiota" OR "bacteriome" OR "amplicon sequencing" OR "16S rRNA" OR "16S rRNA sequencing" OR "shotgun metagenomics" OR "shallow metagenomics" OR "454 pyrosequencing" OR "dgge " OR "denaturating gradient gel electrophoresis" OR "alpha-diversity" OR "beta-diversity" ) OR AB ( "gut microbiota " OR "gut microbiota composition" OR "gut microbiome" OR "gut bacteria" OR Lactobacillus OR Bifidobacterium OR Bacteroides OR Akkermansia OR Ruminococcaceae OR Prevotella OR "Escherichia coli" OR E. coli OR Enterococcus  OR "gut bacteria" OR "fecal microbiota" OR "bacteriome" OR "amplicon sequencing" OR "16S rRNA" OR "16S rRNA sequencing" OR "shotgun metagenomics" OR "shallow metagenomics" OR "454 pyrosequencing" OR "dgge " OR "denaturating gradient gel electrophoresis" OR "alpha-diversity" OR "beta-diversity" ) OR MW ( "gut microbiota " OR "gut microbiota composition" OR "gut microbiome" OR "gut bacteria" OR Lactobacillus OR Bifidobacterium OR Bacteroides OR Akkermansia OR Ruminococcaceae OR Prevotella OR "Escherichia coli" OR E. coli OR Enterococcus  OR "gut bacteria" OR "fecal microbiota" OR "bacteriome" OR "amplicon sequencing" OR "16S rRNA" OR "16S rRNA sequencing" OR "shotgun metagenomics" OR "shallow metagenomics" OR "454 pyrosequencing" OR "dgge " OR "denaturating gradient gel electrophoresis" OR "alpha-diversity" OR "beta-diversity" )
11. TI ( "bile acids" OR "short-chain fatty acids" OR SCFA OR "branched-chain amino acids" OR "BCCA" OR "trimethylamine n-oxide" OR TMAO OR "cholic acid" OR cholate OR "chenodeoxycholic acid" OR chenodeoxycholate OR "taurocholic acid" OR taurocholate OR "glycocholic acid" OR glycocholate OR "taurochenodeoxycholic acid" OR taurochenodeoxycholate OR "glycochenodeoxycholic acid" OR glycochenodeoxycholate OR "glycocholenate sulfate" OR "glycohyocholate" OR "glycolithocholate sulfate" OR glycoursodeoxycholate OR taurocholenate_sulfate OR taurodeoxycholate OR "taurolithocholate_3_sulfate" OR tauroursodeoxycholate OR "deoxycholic Acid" OR "wheat bran" OR deoxycholate OR "lithocholic acid" OR lithocholate OR "acetic acid" OR acetate OR "propionic acid" OR propionate OR "butyric Acid" OR butyrate OR leucine OR isoleucine OR valine OR indole OR tryptophan OR indole-3-lactate OR "indole-3-lactic acid" OR indole-3-propionate OR "indole-3-propionic acid" OR indole-3-aldehyde OR "indoxyl sulphate" OR "IPA" OR " indoxyl sulfate " OR "p-cresyl sulphate" OR "phenol sulfate" OR "phenylacetylglutamine " OR "indoxylsulfuric acid" OR indole-3- acetate OR "indole-3-acetic acid" OR indole-3-carboxylate OR "indole-3-carboxylic acid" OR indole-acetaldehyde OR "p-cresyl sulfate" OR "gut microbiota metabolite*" OR "gut metabolite*" OR "gastrointestinal metabolite*" OR "uremic toxin*" ) OR AB ( "bile acids" OR "short-chain fatty acids" OR SCFA OR "branched-chain amino acids" OR "BCCA" OR "trimethylamine n-oxide" OR TMAO OR "cholic acid" OR cholate OR "chenodeoxycholic acid" OR

chenodeoxycholate OR "taurocholic acid" OR taurocholate OR "glycocholic acid" OR glycocholate OR "taurochenodeoxycholic acid" OR taurochenodeoxycholate OR "glycochenodeoxycholic acid" OR glycochenodeoxycholate OR "glycocholenate sulfate" OR "glycohyocholate" OR "glycolithocholate sulfate" OR glycoursodeoxycholate OR taurocholenate_sulfate OR taurodeoxycholate OR "taurolithocholate_3_sulfate" OR tauroursodeoxycholate OR "deoxycholic Acid" OR "wheat bran" OR deoxycholate OR "lithocholic acid" OR lithocholate OR "acetic acid" OR acetate OR "propionic acid" OR propionate OR "butyric Acid" OR butyrate OR leucine OR isoleucine OR valine OR indole OR tryptophan OR indole-3-lactate OR "indole-3-lactic acid" OR indole-3-propionate OR "indole-3-propionic acid" OR indole-3-aldehyde OR "indoxyl sulphate" OR "IPA" OR " indoxyl sulfate " OR "p-cresyl sulphate" OR "phenol sulfate" OR "phenylacetylglutamine " OR "indoxylsulfuric acid" OR indole-3- acetate OR "indole-3-acetic acid" OR indole-3-carboxylate OR "indole-3-carboxylic acid" OR indole-acetaldehyde OR "p-cresyl sulfate" OR "gut microbiota metabolite*" OR "gut metabolite*" OR "gastrointestinal metabolite*" OR "uremic toxin*" ) OR MW ( "bile acids" OR "short-chain fatty acids" OR SCFA OR "branched-chain amino acids" OR "BCCA" OR "trimethylamine n-oxide" OR TMAO OR "cholic acid" OR cholate OR "chenodeoxycholic acid" OR chenodeoxycholate OR "taurocholic acid" OR taurocholate OR "glycocholic acid" OR glycocholate OR "taurochenodeoxycholic acid" OR taurochenodeoxycholate OR "glycochenodeoxycholic acid" OR glycochenodeoxycholate OR "glycocholenate sulfate" OR "glycohyocholate" OR "glycolithocholate sulfate" OR glycoursodeoxycholate OR taurocholenate_sulfate OR taurodeoxycholate OR "taurolithocholate_3_sulfate" OR tauroursodeoxycholate OR "deoxycholic Acid" OR "wheat bran" OR deoxycholate OR "lithocholic acid" OR lithocholate OR "acetic acid" OR acetate OR "propionic acid" OR propionate OR "butyric Acid" OR butyrate OR leucine OR isoleucine OR valine OR indole OR tryptophan OR indole-3-lactate OR "indole-3-lactic acid" OR indole-3-propionate OR "indole-3-propionic acid" OR indole-3-aldehyde OR "indoxyl sulphate" OR "IPA" OR " indoxyl sulfate " OR "p-cresyl sulphate" OR "phenol sulfate" OR "phenylacetylglutamine " OR "indoxylsulfuric acid" OR indole-3- acetate OR "indole-3-acetic acid" OR indole-3-carboxylate OR "indole-3-carboxylic acid" OR indole-acetaldehyde OR "p-cresyl sulfate" OR "gut microbiota metabolite*" OR "gut metabolite*" OR "gastrointestinal metabolite*" OR "uremic toxin*" )

12. #9 OR #10 OR #11
13. #3 AND #8 AND #12

**Results: 1,205**

**Embase**

1. 'dietary fiber'/exp OR 'cellulose'/exp OR 'polysaccharides'/exp OR 'resistant starch'/exp OR 'chicory'/exp OR 'dextrins'/exp OR 'pectins'/exp OR 'psyllium'/exp OR 'alginic acid'/exp

2. 'dietary fiber':ti,ab,kw OR 'fiber*':ti,ab,kw OR 'dietary fibre':ti,ab,kw OR 'cellulose':ti,ab,kw OR 'hemicellulose*':ti,ab,kw OR 'resistant starch':ti,ab,kw OR 'galactooligosaccharides':ti,ab,kw OR 'dextrin':ti,ab,kw OR 'pectin':ti,ab,kw OR 'beta-glucan':ti,ab,kw OR 'partially-hydrolyzed guar gum':ti,ab,kw OR 'phgg':ti,ab,kw OR 'methylcellulose':ti,ab,kw OR 'fruit':ti,ab,kw OR 'vegetable':ti,ab,kw OR 'polysaccharide*':ti,ab,kw OR 'psyllium*':ti,ab,kw OR 'ispagula*':ti,ab,kw OR 'metamucil*':ti,ab,kw OR 'polymer*':ti,ab,kw OR 'carbohydrate*':ti,ab,kw OR 'dietary carbohydrate*':ti,ab,kw OR 'fructan*':ti,ab,kw OR 'chicory':ti,ab,kw OR 'fructooligosaccharides':ti,ab,kw OR 'arabinoxylan*':ti,ab,kw OR 'asteraceae':ti,ab,kw OR 'fructooligosaccharide*':ti,ab,kw OR 'oligofructose*':ti,ab,kw OR 'inulin':ti,ab,kw OR 'whole grain*':ti,ab,kw OR 'wholegrain':ti,ab,kw OR 'whole meal':ti,ab,kw OR 'whole wheat':ti,ab,kw OR 'edible grain':ti,ab,kw OR 'wheat':ti,ab,kw OR 'rice':ti,ab,kw OR 'brown rice':ti,ab,kw OR 'maize':ti,ab,kw OR 'oat':ti,ab,kw OR 'barley':ti,ab,kw OR 'corn':ti,ab,kw OR 'rye':ti,ab,kw OR 'millet':ti,ab,kw OR 'sorghum':ti,ab,kw OR 'beta-glucan soluble fiber':ti,ab,kw OR 'psyllium husk':ti,ab,kw OR 'locust bean gum':ti,ab,kw OR 'hydroxypropylmethylcellulose':ti,ab,kw OR 'alginate':ti,ab,kw OR 'high amylose starch':ti,ab,kw OR 'resistant maltodextrin':ti,ab,kw OR 'resistant dextrin':ti,ab,kw OR 'resistant starch 2':ti,ab,kw OR 'polydextrose':ti,ab,kw OR 'galactooligosaccharide':ti,ab,kw OR 'glucomannan':ti,ab,kw OR 'acacia':ti,ab,kw OR 'gum arabic':ti,ab,kw OR 'inulin-type fructans':ti,ab,kw
3. #1 OR #2
4. 'kidney disease'/exp OR 'kidney failure, chronic'/exp OR 'kidney failure'/exp OR 'acute kidney failure'/exp
5. 'kidney':ti,ab,kw OR 'renal':ti,ab,kw
6. 'disease':ti,ab,kw OR 'failure':ti,ab,kw OR 'function':ti,ab,kw OR 'insufficiency':ti,ab,kw OR 'dysfunction':ti,ab,kw
7. #5 AND #6
8. #4 OR #7
9. 'gastrointestinal microbiome'/exp OR 'lactobacillus'/exp OR 'bifidobacterium'/exp OR 'bacteroides'/exp OR 'akkermansia'/exp OR 'faecalibacterium'/exp OR 'escherichia coli'/exp OR 'enterococcus'/exp OR 'fecal microbiota transplantation'/exp OR 'sequence analysis'/exp OR 'rna 16s'/exp OR 'rna sequencing'/exp OR 'denaturing gradient gel electrophoresis'/exp OR 'volatile fatty acid'/exp OR 'cholic acid'/exp OR 'propionic acid derivative'/exp OR 'uremic toxins'/exp OR 'trimethylamine oxide'/exp
10. 'gut microbiota':ti,ab,kw OR 'gut microbiota composition':ti,ab,kw OR 'gut microbiome':ti,ab,kw OR lactobacillus:ti,ab,kw OR bifidobacterium:ti,ab,kw OR bacteroides:ti,ab,kw OR akkermansia:ti,ab,kw OR ruminococcaceae:ti,ab,kw OR prevotella:ti,ab,kw OR 'escherichia coli':ti,ab,kw OR 'e. coli':ti,ab,kw OR enterococcus:ti,ab,kw OR 'gut bacteria':ti,ab,kw OR 'fecal microbiota':ti,ab,kw OR 'bacteriome':ti,ab,kw OR 'amplicon sequencing':ti,ab,kw OR '16s rrna':ti,ab,kw OR '16s rrna sequencing':ti,ab,kw OR 'shotgun metagenomics':ti,ab,kw OR 'shallow metagenomics':ti,ab,kw OR '454 pyrosequencing':ti,ab,kw OR 'dgge':ti,ab,kw OR

'denaturating gradient gel electrophoresis':ti,ab,kw OR 'alpha-diversity':ti,ab,kw OR 'beta-diversity':ti,ab,kw

11. 'bile acids':ti,ab,kw OR 'short-chain fatty acids':ti,ab,kw OR scfa:ti,ab,kw OR 'branched-chain amino acids':ti,ab,kw OR 'bcca':ti,ab,kw OR 'trimethylamine n-oxide':ti,ab,kw OR tmao:ti,ab,kw OR 'cholic acid':ti,ab,kw OR cholate:ti,ab,kw OR 'chenodeoxycholic acid':ti,ab,kw OR chenodeoxycholate:ti,ab,kw OR 'taurocholic acid':ti,ab,kw OR taurocholate:ti,ab,kw OR 'glycocholic acid':ti,ab,kw OR glycocholate:ti,ab,kw OR 'taurochenodeoxycholic acid':ti,ab,kw OR taurochenodeoxycholate:ti,ab,kw OR 'glycochenodeoxycholic acid':ti,ab,kw OR glycochenodeoxycholate:ti,ab,kw OR 'glycocholenate sulfate':ti,ab,kw OR 'glycohyocholate':ti,ab,kw OR 'glycolithocholate sulfate':ti,ab,kw OR glycoursodeoxycholate:ti,ab,kw OR taurocholenate_sulfate:ti,ab,kw OR taurodeoxycholate:ti,ab,kw OR 'taurolithocholate_3_sulfate':ti,ab,kw OR tauroursodeoxycholate:ti,ab,kw OR 'deoxycholic acid':ti,ab,kw OR 'wheat bran':ti,ab,kw OR deoxycholate:ti,ab,kw OR 'lithocholic acid':ti,ab,kw OR lithocholate:ti,ab,kw OR 'acetic acid':ti,ab,kw OR acetate:ti,ab,kw OR 'propionic acid':ti,ab,kw OR propionate:ti,ab,kw OR 'butyric acid':ti,ab,kw OR butyrate:ti,ab,kw OR leucine:ti,ab,kw OR isoleucine:ti,ab,kw OR valine:ti,ab,kw OR indole:ti,ab,kw OR tryptophan:ti,ab,kw OR 'indole 3 lactate':ti,ab,kw OR 'indole-3-lactic acid':ti,ab,kw OR 'indole 3 propionate':ti,ab,kw OR 'indole-3-propionic acid':ti,ab,kw OR 'indole 3 aldehyde':ti,ab,kw OR 'indoxyl sulphate':ti,ab,kw OR 'ipa':ti,ab,kw OR 'indoxyl sulfate':ti,ab,kw OR 'p-cresyl sulphate':ti,ab,kw OR 'phenol sulfate':ti,ab,kw OR 'phenylacetylglutamine':ti,ab,kw OR 'indoxylsulfuric acid':ti,ab,kw OR 'indole-3- acetate':ti,ab,kw OR 'indole-3-acetic acid':ti,ab,kw OR 'indole 3 carboxylate':ti,ab,kw OR 'indole-3-carboxylic acid':ti,ab,kw OR 'indole acetaldehyde':ti,ab,kw OR 'p-cresyl sulfate':ti,ab,kw OR 'gut microbiota metabolite*':ti,ab,kw OR 'gut metabolite*':ti,ab,kw OR 'gastrointestinal metabolite*':ti,ab,kw OR 'uremic toxin*':ti,ab,kw
12. #9 OR #10 OR #11
13. #3 AND #8 AND #12

**Results: 3,896**

**Cochrane**

ID Search

#1 MeSH descriptor: [Dietary Fiber] explode all trees

#2 MeSH descriptor: [Cellulose] explode all trees

#3 MeSH descriptor: [Polysaccharides] explode all trees

#4 MeSH descriptor: [Resistant Starch] explode all trees

#5 MeSH descriptor: [Chicory] explode all trees

#6 MeSH descriptor: [Dextrins] explode all trees

#7 MeSH descriptor: [Pectins] explode all trees

#8 MeSH descriptor: [Psyllium] explode all trees

#9 MeSH descriptor: [Alginates] explode all trees

#10 MeSH descriptor: [Zea mays] explode all trees

#11MeSH descriptor: [Gum Arabic] explode all trees

#12 #1 OR #2 OR #3 OR #4 OR #5 OR #6 OR #7 OR #8 OR #9 OR #10 OR #11

#13 ("Dietary fiber" OR fiber OR "dietary fibre" OR "cellulose" OR "hemicellulose" OR "Resistant maltodextrin" OR "resistant starch" OR "galactooligosaccharides" OR "dextrin" OR "Pectin" OR "beta-glucan" OR "partially-hydrolyzed guar gum" OR "PHGG" OR "methylcellulose" OR "fruit" OR "vegetable" OR "polysaccharide" OR "psyllium" OR "ispagula" OR "metamucil" OR polymer OR carbohydrate OR (dietary NEXT carbohydrate) OR fructan OR "chicory" OR "fructooligosaccharides" OR arabinoxylan OR "asteraceae" OR "fructooligosaccharide" OR "oligofructose" OR "inulin" OR (whole NEXT grain) OR "wholegrain" OR "whole meal" OR "whole wheat" OR "edible grain" OR "wheat" OR "rice" OR "brown rice" OR "maize" OR "oat" OR "barley" OR "corn" OR "rye" OR "millet" OR "sorghum" OR "Beta-glucan soluble fiber" OR "Psyllium husk" OR "locust bean gum" OR "Hydroxypropylmethylcellulose" OR "Alginate" OR "High amylose starch" OR "Resistant maltodextrin" OR "Resistant dextrin" OR "resistant starch 2" OR "Polydextrose" OR "Galactooligosaccharide" OR "Glucomannan" OR "Acacia" OR "gum arabic" OR "inulin-type fructans"):ti,ab,kw

#14 #12 OR #13

#15 MeSH descriptor: [Kidney Diseases] explode all trees

#16 MeSH descriptor: [Kidney Failure, Chronic] explode all trees

#17 MeSH descriptor: [Renal Insufficiency] explode all trees

#18 MeSH descriptor: [Acute Kidney Injury] explode all trees

#19 #15 OR #16 OR #17 OR #18

#20 ("Kidney" OR "renal"):ti,ab,kw

#21 ("Disease" OR "failure" OR "function" OR "insufficiency" OR "dysfunction"):ti,ab,kw

#22 #20 AND #21

#23 #19 OR #22

#24 MeSH descriptor: [Gastrointestinal Microbiome] explode all trees

#25 MeSH descriptor: [Lactobacillus] explode all trees

#26 MeSH descriptor: [Bifidobacterium] explode all trees

#27 MeSH descriptor: [Bacteroides] explode all trees

#28 MeSH descriptor: [Akkermansia] explode all trees

#29 MeSH descriptor: [Faecalibacterium prausnitzii] explode all trees

#30 MeSH descriptor: [Prevotella] explode all trees

#31 MeSH descriptor: [Escherichia coli] explode all trees

#32 MeSH descriptor: [Enterococcus] explode all trees

#33 MeSH descriptor: [Fecal Microbiota Transplantation] explode all trees

#34 MeSH descriptor: [Gastrointestinal Microbiome] explode all trees

#35 MeSH descriptor: [Sequence Analysis] explode all trees

#36 MeSH descriptor: [RNA, Ribosomal, 16S] explode all trees

#37 MeSH descriptor: [Fatty Acids, Volatile] explode all trees

#38 MeSH descriptor: [Cholic Acid] explode all trees

#39 MeSH descriptor: [Cholic Acid] explode all trees

#40 MeSH descriptor: [Propionates] explode all trees

#41 MeSH descriptor: [Uremic Toxins] explode all trees

#42 #24 OR #25 OR #26 OR #27 OR #28 OR #29 OR #30 OR #31 OR #32 OR #33 OR #34 OR #35 OR #36 OR #37 OR #38 OR #39 OR #40 OR #41

#43 ("gut microbiota " OR "gut microbiota composition" OR "gut microbiome" OR "gut bacteria" OR Lactobacillus OR Bifidobacterium OR Bacteroides OR Akkermansia OR Ruminococcaceae OR Prevotella OR "Escherichia coli" OR E. coli OR Enterococcus OR "gut bacteria" OR "fecal microbiota" OR "bacteriome" OR "amplicon sequencing" OR "16S rRNA" OR "16S rRNA sequencing" OR "shotgun metagenomics" OR "shallow metagenomics" OR "454 pyrosequencing" OR "dgge " OR "denaturating gradient gel electrophoresis" OR "alpha-diversity" OR "beta-diversity"):ti,ab,kw

#44 ("bile acids" OR "short-chain fatty acids" OR SCFA OR "branched-chain amino acids" OR "BCCA" OR "trimethylamine n-oxide" OR TMAO OR "cholic acid" OR cholate OR "chenodeoxycholic acid" OR chenodeoxycholate OR "taurocholic acid" OR taurocholate OR "glycocholic acid" OR glycocholate OR "taurochenodeoxycholic acid" OR taurochenodeoxycholate OR "glycochenodeoxycholic acid" OR glycochenodeoxycholate OR "glycocholenate sulfate" OR "glycohyocholate" OR "glycolithocholate sulfate" OR glycoursodeoxycholate OR taurocholenate_sulfate OR taurodeoxycholate OR "taurolithocholate_3_sulfate"):ti,ab,kw

#45 (tauroursodeoxycholate OR "deoxycholic Acid" OR "wheat bran" OR deoxycholate OR "lithocholic acid" OR lithocholate OR "acetic acid" OR acetate OR "propionic acid" OR propionate OR "butyric Acid" OR butyrate OR leucine OR isoleucine OR valine OR indole OR tryptophan):ti,ab,kw

#46 ("indole 3 lactate" OR "indole 3 lactic acid" OR indole 3 propionate OR "indole 3 propionic acid" OR indole 3 aldehyde OR "indoxyl sulphate" OR "IPA" OR " indoxyl sulfate " OR "p cresyl sulphate" OR "phenol sulfate" OR "phenylacetylglutamine " OR "indoxylsulfuric acid" OR indole 3 acetate OR "indole 3 acetic acid" OR indole 3 carboxylate OR "indole 3 carboxylic acid" OR indole acetaldehyde OR "p cresyl sulfate" OR "gut microbiota metabolite" OR "gut metabolite" OR "gastrointestinal metabolite" OR "uremic toxin"):ti,ab,kw

#47 #42 OR #43 OR #44 OR #45 OR #46

#48 #14 AND #23 AND #47

**Results: 276**

**All results= 8517**

**Codes in Stata SE version 19.0**

The bolded elements denote the sections of code that require modification according to the study variables. A structured color scheme was applied to support clarity and reproducibility, with annotations displayed in green, variable-dependent components in blue, and the fixed code in black.

**General Code guide**

gen **Label_Name** = **Variable_of_ Interest** + " (" + string(**Year**) + ")" # Generate a study label to be used in forest plot

**metan Intervention_Sample_Size Mean_of_Interevnetion_Variable SD_of_Intervention Control_Sample_Size Mean_of_Control_Variable SD_of_Control**, random md label (namevar= **Label_Name** ) # For variables reported in the same units, the analysis used the weighted mean difference (WMD) based on study-level means and standard deviations. For variables reported in different units, or when substantial heterogeneity was expected due to differences in animal strains, no WMD specification was required; *Stata* automatically calculated the standardized mean difference (SMD).

**metan Intervention_Sample_Size Mean_of_Interevnetion_Variable SD_of_Intervention Control_Sample_Size Mean_of_Control_Variable SD_of_Control**, random label (namevar= **Label_Name** ) # For variables reported in the different units or species of animals

**metan Intervention_Sample_Size Mean_of_Interevnetion_Variable SD_of_Intervention Control_Sample_Size Mean_of_Control_Variable SD_of_Control**, random md label (namevar= **Label_Name** ) by (**Subroup_of_Interest**) # For subgroup analysis of interest, best robist with having at least two studies in each subgroup

metabias _ES _seES # For publication bias evaluation (Egg and Begger`s Test)

metafunnel _ES _seES # For funnel plot

destring **Variable_of_Interest**, replace # change string variable to numeric

metareg **Variable_of_Interest** _ES , wsse ( _seES ) graph # Meta-regression based on the variable of interest

**Statistical Codes for RCTs in Stata SE**

gen StudyLabel = Study + " (" + string(Year) + ")"

destring Dosegday, replace # change string variable to numeric

**Total serum/plasma IAA**

metan INsam TotalSerum_IAA_Change_In BN Csam TotalSerum_IAA_Change_Pl BP, random md label (namevar= StudyLabel ) # Meta-analysis

metan INsam TotalSerum_IAA_Change_In BN Csam TotalSerum_IAA_Change_Pl BP, random md label (namevar= StudyLabel ) influence # Sensitivity Analysis

metabias _ES _seES # For publication bias evaluation (Egg and Begger`s Test)

metafunnel _ES _seES # For funnel plot

**Free serum/plasma IAA**

metan INsam FreeSerum_IAA_Change_In BJ Csam FreeSerum_IAA_Change_Pl BL, random md label (namevar= StudyLabel ) # Meta-analysis

metan INsam FreeSerum_IAA_Change_In BJ Csam FreeSerum_IAA_Change_Pl BL, random md label (namevar= StudyLabel )  influence # Sensitivity Analysis

metabias _ES _seES # For publication bias evaluation (Egg and Begger`s Test)

metafunnel _ES _seES # For funnel plot

**Total serum/plasma TMAO**

metan INsam SerumPlasma_TMAO_Change_In AP Csam SerumPlasma_TMAO_Change_Pl AR, random md label (namevar= StudyLabel ) # Meta-analysis

metan INsam SerumPlasma_TMAO_Change_In AP Csam SerumPlasma_TMAO_Change_Pl AR, random md label (namevar= StudyLabel )  influence # Sensitivity Analysis

metabias _ES _seES # For publication bias evaluation (Egg and Begger`s Test)

metafunnel _ES _seES # For funnel plot

**Total serum/plasma IS**

metan INsam TotalSerumplasma_IS_Change_In BB Csam TotalSerumplasma_IS_Change_Pl BD, random md label (namevar= StudyLabel ) # Meta-analysis

metan INsam TotalSerumplasma_IS_Change_In BB Csam TotalSerumplasma_IS_Change_Pl BD, random md label (namevar= StudyLabel )  influence # Sensitivity Analysis

metabias _ES _seES # For publication bias evaluation (Egg and Begger`s Test)

metafunnel _ES _seES # For funnel plot

**Subgroup Analysis of total serum/plasma IS**

metan INsam TotalSerumplasma_IS_Change_In BB Csam TotalSerumplasma_IS_Change_Pl BD, random md label (namevar= StudyLabel ) by (Fiber_Code) # subgroup analysis by fiber type

metan INsam TotalSerumplasma_IS_Change_In BB Csam TotalSerumplasma_IS_Change_Pl BD, random md label (namevar= StudyLabel ) by (dur_Cod) ) # subgroup analysis by duration metan INsam TotalSerumplasma_IS_Change_In BB Csam TotalSerumplasma_IS_Change_Pl BD, random md label (namevar= StudyLabel ) by (dose_cod) ) # subgroup analysis by fiber intervention dose

metan INsam TotalSerumplasma_IS_Change_In BB Csam TotalSerumplasma_IS_Change_Pl BD, random md label (namevar= StudyLabel ) by (participant_Dialysis_COD) ) # subgroup analysis by dialysis status

**Meta regression of total serum/plasma IS**

metareg Dosegday _ES , wsse ( _seES ) graph # Meta-regression based on fiber dose

metareg Durationofinterventionweek _ES , wsse ( _seES ) graph # Meta-regression based on dutaion of intervention

**Dose response analysis of total serum/plasma IS**

fracpoly regress TotalSerumplasma_IS_Change_In Dosegday # Dose response analysis based on fiber dose

twoway fpfitci TotalSerumplasma_IS_Change_In Dosegday ||scatter TotalSerumplasma_IS_Change_In Dosegday # Scatterplot of dose reponse analysis

fracpoly regress TotalSerumplasma_IS_Change_In Durationofinterventionweek # Dose response analysis based on duration of intervention

twoway TotalSerumplasma_IS_Change_In Durationofinterventionweek ||scatter TotalSerumplasma_IS_Change_In Durationofinterventionweek # Scatterplot of dose response analysis

**Free serum/plasma IS**

metan INsam FreeSerumplasma_IS_Change_In AX Csam FreeSerumplasma_IS_Change_Pl AZ, random md label (namevar= StudyLabel ) # Meta-analysis

metan INsam FreeSerumplasma_IS_Change_In AX Csam FreeSerumplasma_IS_Change_Pl AZ, random md label (namevar= StudyLabel ) influence # Sensitivity Analysis

metabias _ES _seES # For publication bias evaluation (Egg and Begger`s Test)

metafunnel _ES _seES # For funnel plot

**Urinary IS**

metan INsam Urine_IS_Change_In BF Csam Urine_IS_Change_Pl BH, random md label (namevar= StudyLabel ) # Meta-analysis metan INsam Urine_IS_Change_In BF Csam Urine_IS_Change_Pl BH, random md label (namevar= StudyLabel ) influence # Sensitivity Analysis

metabias _ES _seES # For publication bias evaluation (Egg and Begger`s Test)

metafunnel _ES _seES # For funnel plot

**Total serum/plasma pCS**

metan INsam TotalSerumplasma_pCS_Change_I BV Csam TotalSerumplasma_pCS_Change_P BX, random md label (namevar= StudyLabel )# Meta-analysis

metan INsam TotalSerumplasma_pCS_Change_I BV Csam TotalSerumplasma_pCS_Change_P BX, random md label (namevar= StudyLabel ) influence # Sensitivity Analysis

metabias _ES _seES # For publication bias evaluation (Egg and Begger`s Test)

metafunnel _ES _seES # For funnel plot

**Subgroup Analysis of total serum/plasma pCS**

metan INsam TotalSerumplasma_pCS_Change_I BV Csam TotalSerumplasma_pCS_Change_P BX, random md label (namevar= StudyLabel )by (Fiber_Code) # subgroup analysis by fiber type

metan INsam TotalSerumplasma_pCS_Change_I BV Csam TotalSerumplasma_pCS_Change_P BX, random md label (namevar= StudyLabel ) by (dur_Cod) # subgroup analysis by duration

metan INsam TotalSerumplasma_pCS_Change_I BV Csam TotalSerumplasma_pCS_Change_P BX, random md label (namevar= StudyLabel ) by (dose_cod) # subgroup analysis by fiber intervention dose

metan INsam TotalSerumplasma_pCS_Change_I BV Csam TotalSerumplasma_pCS_Change_P BX, random md label (namevar= StudyLabel ) by (participant_Dialysis_COD) # subgroup analysis by dialysis status

**Meta regression of total serum/plasma pCS**

metareg Dosegday _ES , wsse ( _seES ) graph # Meta-regression based on fiber dose

metareg Durationofinterventionweek _ES , wsse ( _seES ) graph # Meta-regression based on dutaion of intervention

**Dose response analysis of total serum/plasma pCS**

fracpoly regress TotalSerumplasma_pCS_Change_I Dosegday # Dose response analysis based on fiber dose twoway fpfitci TotalSerumplasma_pCS_Change_I Dosegday ||scatter TotalSerumplasma_pCS_Change_I Dosegday # Scatterplot of dose response analysis

fracpoly regress TotalSerumplasma_pCS_Change_I Durationofinterventionweek # Dose response analysis based on duration of intervention

twoway fpfitci TotalSerumplasma_pCS_Change_I Durationofinterventionweek ||scatter TotalSerumplasma_pCS_Change_I Durationofinterventionweek # Scatterplot of dose response analysis

**Free serum/plasma pCS**

metan INsam FreeSerumplasma_pCS_Change_In BR Csam FreeSerumplasma_pCS_Change_Pl BT, random md label (namevar= StudyLabel ) # Meta-analysis

metan INsam FreeSerumplasma_pCS_Change_In BR Csam FreeSerumplasma_pCS_Change_Pl BT, random md label (namevar= StudyLabel ) influence # Sensitivity Analysis

metabias _ES _seES # For publication bias evaluation (Egg and Begger`s Test)

metafunnel _ES _seES # For funnel plot

**Urinary pCS**

metan INsam Urine_pCS_Change_In BZ Csam Urine_pCS_Change_Pl CB, random md label (namevar= StudyLabel ) # Meta-analysis

metan INsam Urine_pCS_Change_In BZ Csam Urine_pCS_Change_Pl CB, random md label (namevar= StudyLabel ) influence # Sensitivity Analysis

metabias _ES _seES # For publication bias evaluation (Egg and Begger`s Test)

metafunnel _ES _seES # For funnel plot

**Statistical Codes for Animal Models in Stata SE**

**Total serum/plasma IS**

metan INsam SerumPlasma_IS_In CB Csam SerumPlasma_IS_Pla BZ, random label (namevar= StudyLabel ) # Meta-analysis

metan INsam SerumPlasma_IS_In CB Csam SerumPlasma_IS_Pla BZ, random label (namevar= StudyLabel ) influence # Sensitivity Analysis

metabias _ES _seES # For publication bias evaluation (Egg and Begger`s Test)

metafunnel _ES _seES # For funnel plot

**Subgroup Analysis of total serum/plasma IS**

metan INsam SerumPlasma_IS_In CB Csam SerumPlasma_IS_Pla BZ, random md label (namevar= StudyLabel ) by (Fiber_Code) # subgroup analysis by fiber type

metan INsam SerumPlasma_IS_In CB Csam SerumPlasma_IS_Pla BZ, random md label (namevar= StudyLabel ) by (Fermentability) # subgroup analysis by fermentability

metan INsam SerumPlasma_IS_In CB Csam SerumPlasma_IS_Pla BZ, random md label (namevar= StudyLabel ) by (Viscosity) # subgroup analysis by viscosity

metan INsam SerumPlasma_IS_In CB Csam SerumPlasma_IS_Pla BZ, random md label (namevar= StudyLabel )by (dur_Cod) # subgroup analysis by duration

metan INsam SerumPlasma_IS_In CB Csam SerumPlasma_IS_Pla BZ, random md label (namevar= StudyLabel )by (dose_cod) # subgroup analysis by fiber intervention dose

**Meta regression of total serum/plasma IS**

metareg Doseww_ES , wsse ( _seES ) graph # Meta-regression based on fiber dose

metareg Durationofinterventionweek _ES , wsse ( _seES ) graph # Meta-regression based on dutaion of intervention

**Dose response analysis of total serum/plasma IS**

fracpoly regress SerumPlasma_IS_In Doseww # Dose response analysis based on fiber dose

twoway fpfitci SerumPlasma_IS_In Doseww ||scatter SerumPlasma_IS_In Doseww # Scatterplot of dose reponse analysis

fracpoly regress SerumPlasma_IS_In Durationofinterventionweek # Dose response analysis based on duration of intervention

twoway SerumPlasma_IS_In Durationofinterventionweek ||scatter SerumPlasma_IS_In Durationofinterventionweek # Scatterplot of dose response analysis

**Total serum/plasma pCS**

metan INsam SerumPlasma_pCS_In CI Csam SerumPlasma_pCS_Pla CG, random label (namevar= StudyLabel ) # Meta-analysis metan INsam SerumPlasma_pCS_In CI Csam SerumPlasma_pCS_Pla CG, random label (namevar= StudyLabel ) influence # Sensitivity Analysis

metabias _ES _seES # For publication bias evaluation (Egg and Begger`s Test)

metafunnel _ES _seES # For funnel plot

**Serum/plasma acetate**

metan INsam SerumPlasma_Acetate_In BG Csam SerumPlasma_Acetate_Pla BE, random label (namevar= StudyLabel ) # Meta-analysis

metan INsam SerumPlasma_Acetate_In BG Csam SerumPlasma_Acetate_Pla BE, random label (namevar= StudyLabel ) influence # Sensitivity Analysis

metabias _ES _seES # For publication bias evaluation (Egg and Begger`s Test)

metafunnel _ES _seES # For funnel plot

**Cecal acetate**

metan INsam Cecal_Acetate_In BN Csam Cecal_Acetate_Pla BL, random label (namevar= StudyLabel ) # Meta-analysis

etan INsam Cecal_Acetate_In BN Csam Cecal_Acetate_Pla BL, random label (namevar= StudyLabel ) influence # Sensitivity Analysis

etan INsam Cecal_Acetate_In BN Csam Cecal_Acetate_Pla BL, random label (namevar= StudyLabel ) by (Fiber_Code) # subgroup analysis by fiber type

metabias _ES _seES # For publication bias evaluation (Egg and Begger`s Test)

metafunnel _ES _seES # For funnel plot

**Serum/plasma butyrate**

metan INsam SerumPlasma_Butyrate_In AS Csam SerumPlasma_Butyrate_Pla AQ, random label (namevar= StudyLabel ) # Meta-analysis

metan INsam SerumPlasma_Butyrate_In AS Csam SerumPlasma_Butyrate_Pla AQ, random label (namevar= StudyLabel ) influence # Sensitivity Analysis

metabias _ES _seES # For publication bias evaluation (Egg and Begger`s Test)

metafunnel _ES _seES # For funnel plot

**Cecal butyrate**

metan INsam Cecal_Butyrate_In AZ Csam Cecal_Butyrate_Pla AX, random label (namevar= StudyLabel ) # Meta-analysis

metan INsam Cecal_Butyrate_In AZ Csam Cecal_Butyrate_Pla AX, random label (namevar= StudyLabel ) influence # Sensitivity Analysis

metabias _ES _seES # For publication bias evaluation (Egg and Begger`s Test)

metafunnel _ES _seES # For funnel plot

**Serum/plasma propionate**

metan INsam SerumPlasma_Propionate_In AE Csam SerumPlasma_Propionate_Pla SD, random label (namevar= StudyLabel ) # Meta-analysis

metan INsam SerumPlasma_Propionate_In AE Csam SerumPlasma_Propionate_Pla SD, random label (namevar= StudyLabel ) influence # Sensitivity Analysis

metabias _ES _seES # For publication bias evaluation (Egg and Begger`s Test)

metafunnel _ES _seES # For funnel plot

**Cecal propionate**

metan INsam Cecal_Propionate_In AL Csam Cecal_Propionate_Pla AJ, random label (namevar= StudyLabel ) # Meta-analysis

metan INsam Cecal_Propionate_In AL Csam Cecal_Propionate_Pla AJ, random label (namevar= StudyLabel ) influence # Sensitivity Analysis

metabias _ES _seES # For publication bias evaluation (Egg and Begger`s Test)

metafunnel _ES _seES # For funnel plot

a)

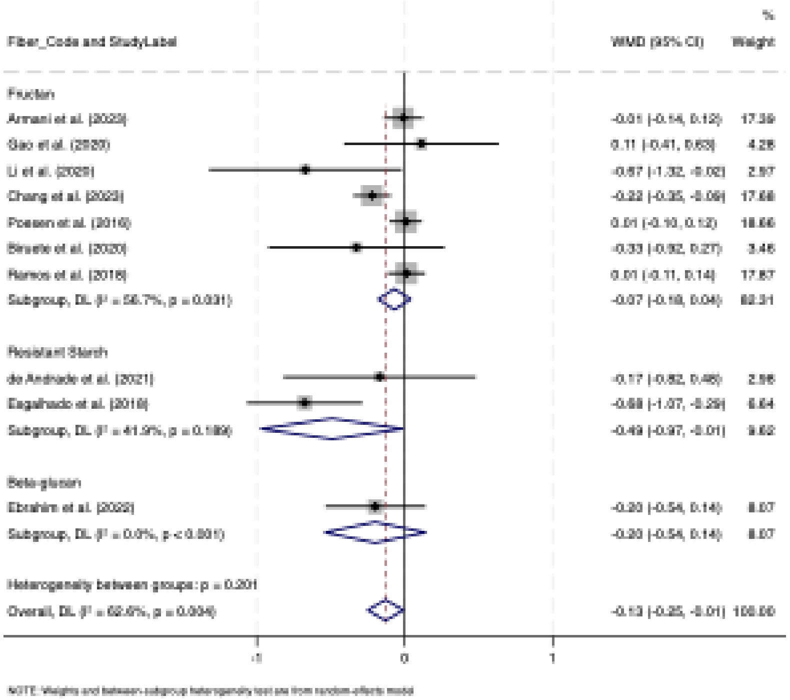

Fiber_Code and StudyLabel
WMD (95% CI)
%
Weight
Fructan
Armani et al. (2023) -0.01 (-0.14, 0.12) 17.39
Gao et al. (2020) 0.11 (-0.41, 0.63) 4.28
Li et al. (2020) -0.67 (-1.32, -0.02) 2.97
Chang et al. (2023) -0.22 (-0.35, -0.09) 17.68
Poesen et al. (2016) 0.01 (-0.10, 0.12) 18.66
Biruete et al. (2020) -0.33 (-0.92, 0.27) 3.46
Ramos et al. (2018) 0.01 (-0.11, 0.14) 17.87
Subgroup, DL (I² = 56.7%, p = 0.031) -0.07 (-0.18, 0.04) 82.31
Resistant Starch
de Andrade et al. (2021) -0.17 (-0.82, 0.48) 2.98
Esgalhado et al. (2018) -0.68 (-1.07, -0.29) 6.64
Subgroup, DL (I² = 41.9%, p = 0.189) -0.49 (-0.97, -0.01) 9.62
Beta-glucan
Ebrahim et al. (2022) -0.20 (-0.54, 0.14) 8.07
Subgroup, DL (I² = 0.0%, p < 0.001) -0.20 (-0.54, 0.14) 8.07
Heterogeneity between groups: p = 0.201
Overall, DL (I² = 62.6%, p = 0.004) -0.13 (-0.25, -0.01) 100.00
-1
0
1
NOTE: Weights and between-subgroup heterogeneity test are from random-effects model


b)

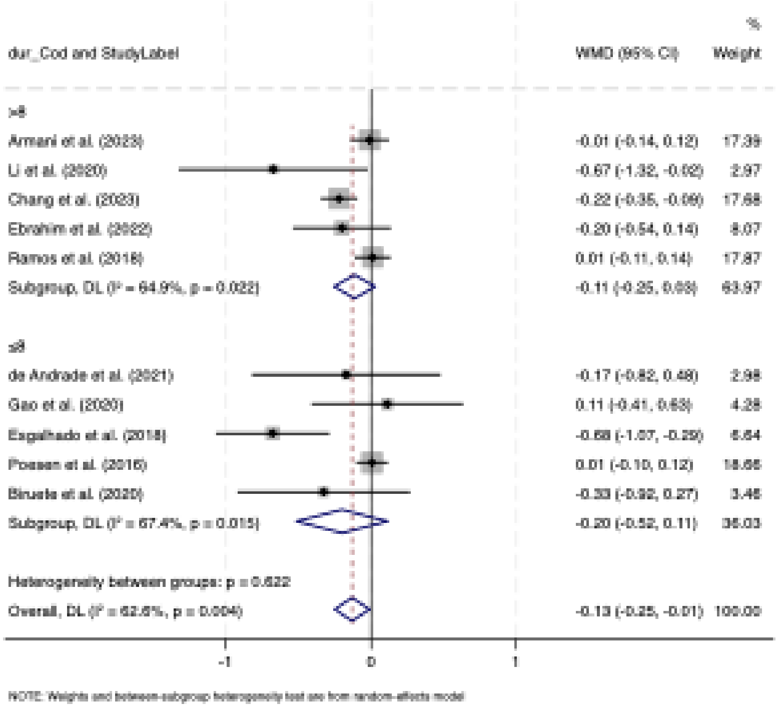

dur_Cod and StudyLabel
WMD (95% CI)
%
Weight
>8
Armani et al. (2023) -0.01 (-0.14, 0.12) 17.39
Li et al. (2020) -0.67 (-1.32, -0.02) 2.97
Chang et al. (2023) -0.22 (-0.35, -0.09) 17.68
Ebrahim et al. (2022) -0.20 (-0.54, 0.14) 8.07
Ramos et al. (2018) 0.01 (-0.11, 0.14) 17.87
Subgroup, DL (I² = 64.9%, p = 0.022) -0.11 (-0.25, 0.03) 63.97
≤8
de Andrade et al. (2021) -0.17 (-0.82, 0.48) 2.98
Gao et al. (2020) 0.11 (-0.41, 0.63) 4.28
Esgalhado et al. (2018) -0.68 (-1.07, -0.29) 6.64
Poesen et al. (2016) 0.01 (-0.10, 0.12) 18.66
Biruete et al. (2020) -0.33 (-0.92, 0.27) 3.46
Subgroup, DL (I² = 67.4%, p = 0.015) -0.20 (-0.52, 0.11) 36.03
Heterogeneity between groups: p = 0.622
Overall, DL (I² = 62.6%, p = 0.004) -0.13 (-0.25, -0.01) 100.00
-1
0
1
NOTE: Weights and between-subgroup heterogeneity test are from random-effects model


c)

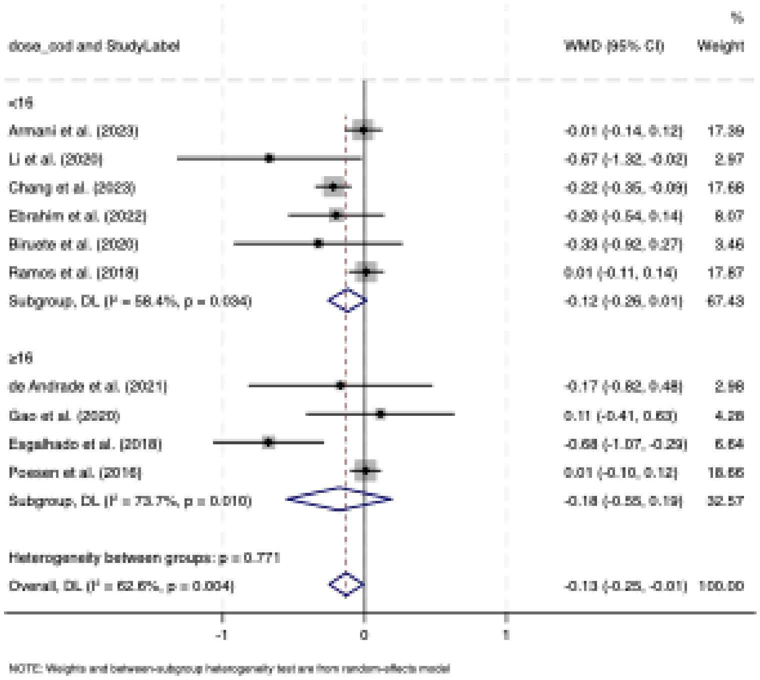

%
dose_cod and StudyLabel
WMD (95% CI)
Weight
<16
Armani et al. (2023)
-0.01 (-0.14, 0.12)
17.39
Li et al. (2020)
-0.67 (-1.32, -0.02)
2.97
Chang et al. (2023)
-0.22 (-0.35, -0.09)
17.68
Ebrahim et al. (2022)
-0.20 (-0.54, 0.14)
8.07
Biruete et al. (2020)
-0.33 (-0.92, 0.27)
3.46
Ramos et al. (2018)
0.01 (-0.11, 0.14)
17.87
Subgroup, DL (I² = 58.4%, p = 0.034)
-0.12 (-0.26, 0.01)
67.43
≥16
de Andrade et al. (2021)
-0.17 (-0.82, 0.48)
2.98
Gao et al. (2020)
0.11 (-0.41, 0.63)
4.28
Esgalhado et al. (2018)
-0.68 (-1.07, -0.29)
6.64
Poesen et al. (2016)
0.01 (-0.10, 0.12)
18.66
Subgroup, DL (I² = 73.7%, p = 0.010)
-0.18 (-0.55, 0.19)
32.57
Heterogeneity between groups: p = 0.771
Overall, DL (I² = 62.6%, p = 0.004)
-0.13 (-0.25, -0.01)
100.00
-1
0
1
NOTE: Weights and between-subgroup heterogeneity test are from random-effects model


d)

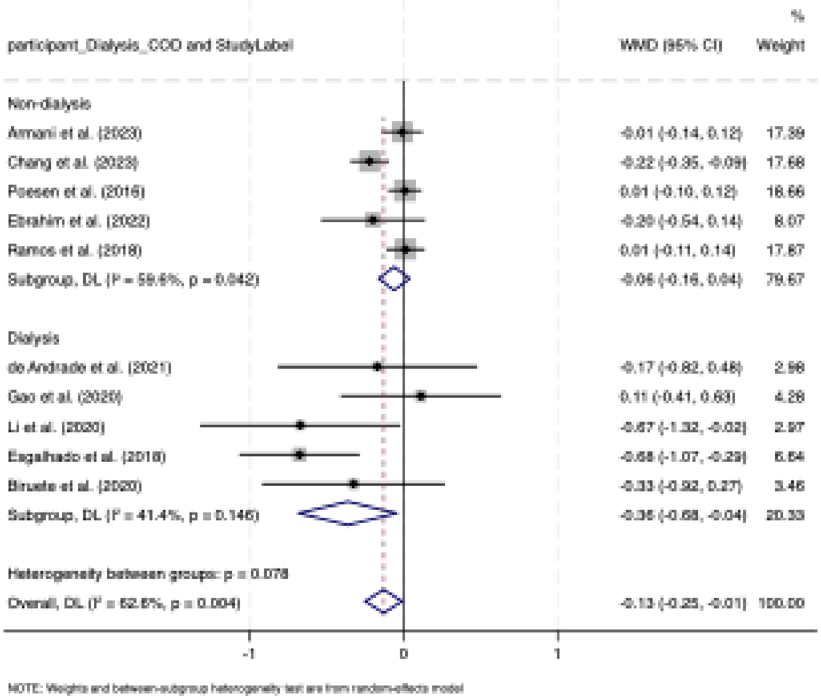

%
participant_Dialysis_COD and StudyLabel
WMD (95% CI)
Weight
Non-dialysis
Armani et al. (2023)
-0.01 (-0.14, 0.12)
17.39
Chang et al. (2023)
-0.22 (-0.35, -0.09)
17.68
Poesen et al. (2016)
0.01 (-0.10, 0.12)
18.66
Ebrahim et al. (2022)
-0.20 (-0.54, 0.14)
8.07
Ramos et al. (2018)
0.01 (-0.11, 0.14)
17.87
Subgroup, DL (I² = 59.6%, p = 0.042)
-0.06 (-0.16, 0.04)
79.67
Dialysis
de Andrade et al. (2021)
-0.17 (-0.82, 0.48)
2.98
Gao et al. (2020)
0.11 (-0.41, 0.63)
4.28
Li et al. (2020)
-0.67 (-1.32, -0.02)
2.97
Esgalhado et al. (2018)
-0.68 (-1.07, -0.29)
6.64
Biruete et al. (2020)
-0.33 (-0.92, 0.27)
3.46
Subgroup, DL (I² = 41.4%, p = 0.146)
-0.36 (-0.68, -0.04)
20.33
Heterogeneity between groups: p = 0.078
Overall, DL (I² = 62.6%, p = 0.004)
-0.13 (-0.25, -0.01)
100.00
-1
0
1
NOTE: Weights and between-subgroup heterogeneity test are from random-effects model


e)

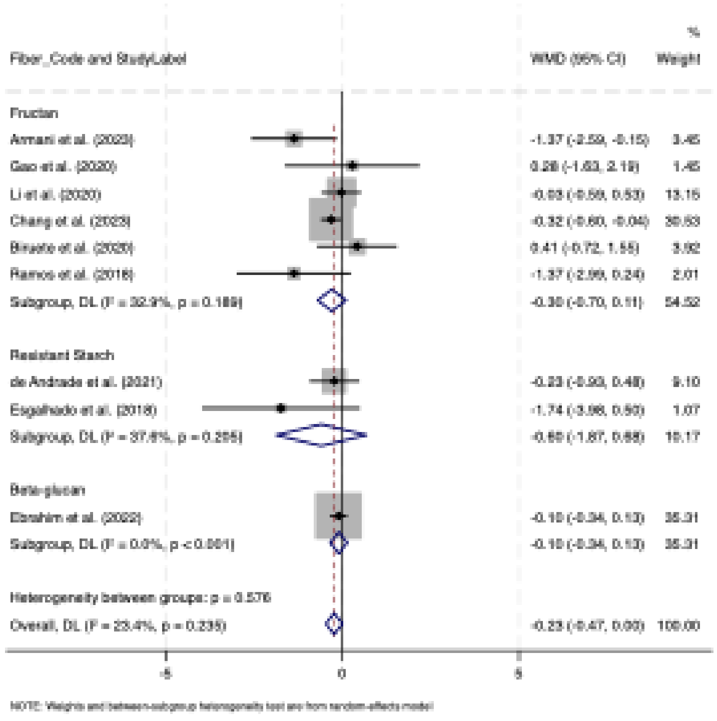
%
Fiber_Code and StudyLabel
WMD (95% CI)
Weight
Fructan
Armani et al. (2023)
-1.37 (-2.59, -0.15)
3.45
Gao et al. (2020)
0.28 (-1.63, 2.19)
1.45
Li et al. (2020)
-0.03 (-0.59, 0.53)
13.15
Chang et al. (2023)
-0.32 (-0.60, -0.04)
30.53
Biruete et al. (2020)
0.41 (-0.72, 1.55)
3.92
Ramos et al. (2018)
-1.37 (-2.99, 0.24)
2.01
Subgroup, DL (I² = 32.9%, p = 0.189)
-0.30 (-0.70, 0.11)
54.52
Resistant Starch
de Andrade et al. (2021)
-0.23 (-0.93, 0.48)
9.10
Esgalhado et al. (2018)
-1.74 (-3.98, 0.50)
1.07
Subgroup, DL (I² = 37.8%, p = 0.205)
-0.60 (-1.87, 0.68)
10.17
Beta-glucan
Ebrahim et al. (2022)
-0.10 (-0.34, 0.13)
35.31
Subgroup, DL (I² = 0.0%, p < 0.001)
-0.10 (-0.34, 0.13)
35.31
Heterogeneity between groups: p = 0.576
Overall, DL (I² = 23.4%, p = 0.235)
-0.23 (-0.47, 0.00)
100.00
-5
0
5
NOTE: Weights and between-subgroup heterogeneity test are from random-effects model

f)

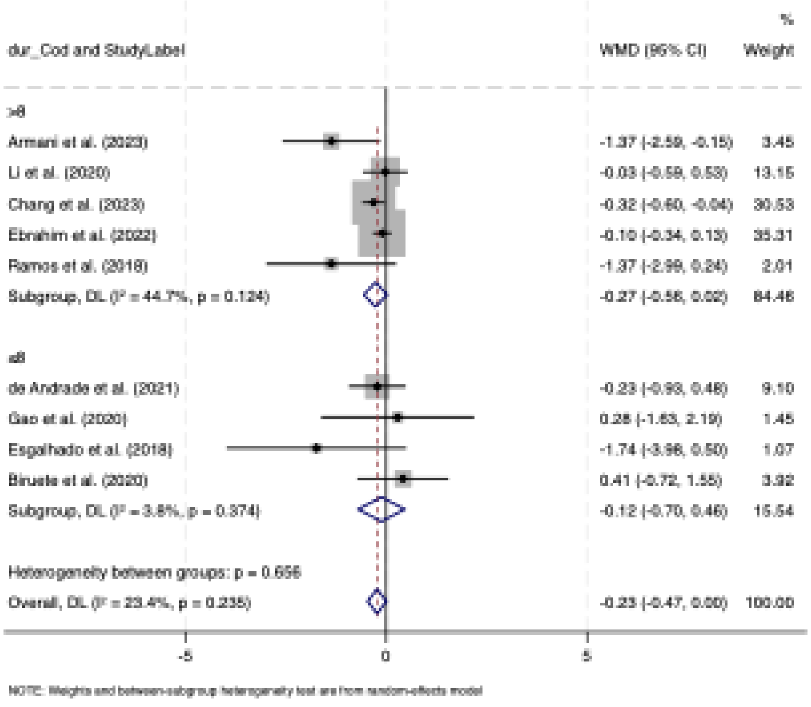
%
dur_Cod and StudyLabel
WMD (95% CI)
Weight
>8
Armani et al. (2023)
-1.37 (-2.59, -0.15)
3.45
Li et al. (2020)
-0.03 (-0.59, 0.53)
13.15
Chang et al. (2023)
-0.32 (-0.60, -0.04)
30.53
Ebrahim et al. (2022)
-0.10 (-0.34, 0.13)
35.31
Ramos et al. (2018)
-1.37 (-2.99, 0.24)
2.01
Subgroup, DL (I² = 44.7%, p = 0.124)
-0.27 (-0.56, 0.02)
84.46
≤8
de Andrade et al. (2021)
-0.23 (-0.93, 0.48)
9.10
Gao et al. (2020)
0.28 (-1.63, 2.19)
1.45
Esgalhado et al. (2018)
-1.74 (-3.98, 0.50)
1.07
Biruete et al. (2020)
0.41 (-0.72, 1.55)
3.92
Subgroup, DL (I² = 3.8%, p = 0.374)
-0.12 (-0.70, 0.46)
15.54
Heterogeneity between groups: p = 0.656
Overall, DL (I² = 23.4%, p = 0.235)
-0.23 (-0.47, 0.00)
100.00
-5
0
5
NOTE: Weights and between-subgroup heterogeneity test are from random-effects model

g)

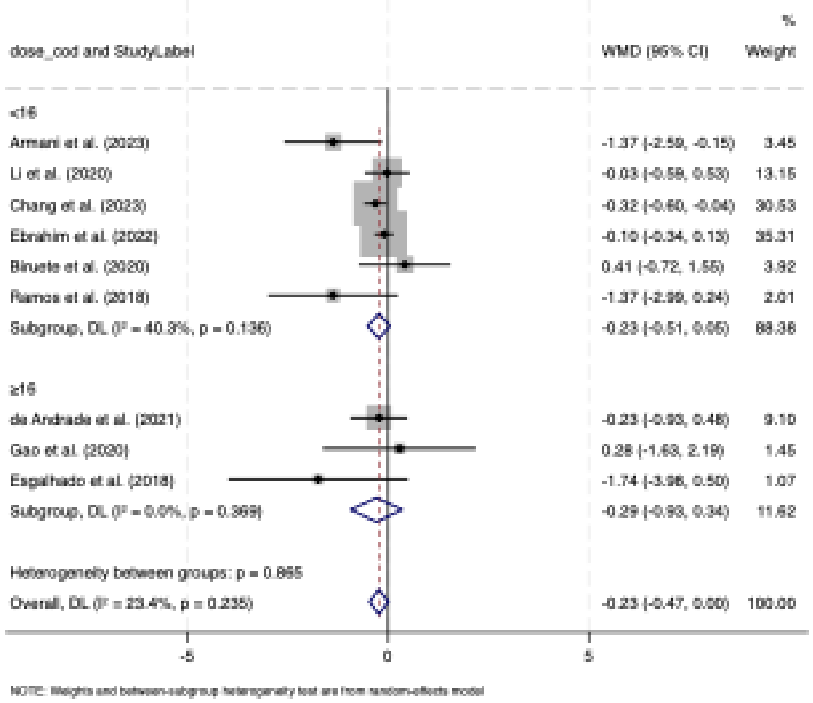
%
dose_cod and StudyLabel
WMD (95% CI)
Weight
<16
Armani et al. (2023)
-1.37 (-2.59, -0.15)
3.45
Li et al. (2020)
-0.03 (-0.59, 0.53)
13.15
Chang et al. (2023)
-0.32 (-0.60, -0.04)
30.53
Ebrahim et al. (2022)
-0.10 (-0.34, 0.13)
35.31
Biruete et al. (2020)
0.41 (-0.72, 1.55)
3.92
Ramos et al. (2018)
-1.37 (-2.99, 0.24)
2.01
Subgroup, DL (I² = 40.3%, p = 0.136)
-0.23 (-0.51, 0.05)
88.38
≥16
de Andrade et al. (2021)
-0.23 (-0.93, 0.46)
9.10
Gao et al. (2020)
0.28 (-1.63, 2.19)
1.45
Esgalhado et al. (2018)
-1.74 (-3.98, 0.50)
1.07
Subgroup, DL (I² = 0.0%, p = 0.369)
-0.29 (-0.93, 0.34)
11.62
Heterogeneity between groups: p = 0.865
Overall, DL (I² = 23.4%, p = 0.235)
-0.23 (-0.47, 0.00)
100.00
-5
0
5
NOTE: Weights and between-subgroup heterogeneity test are from random-effects model

h)

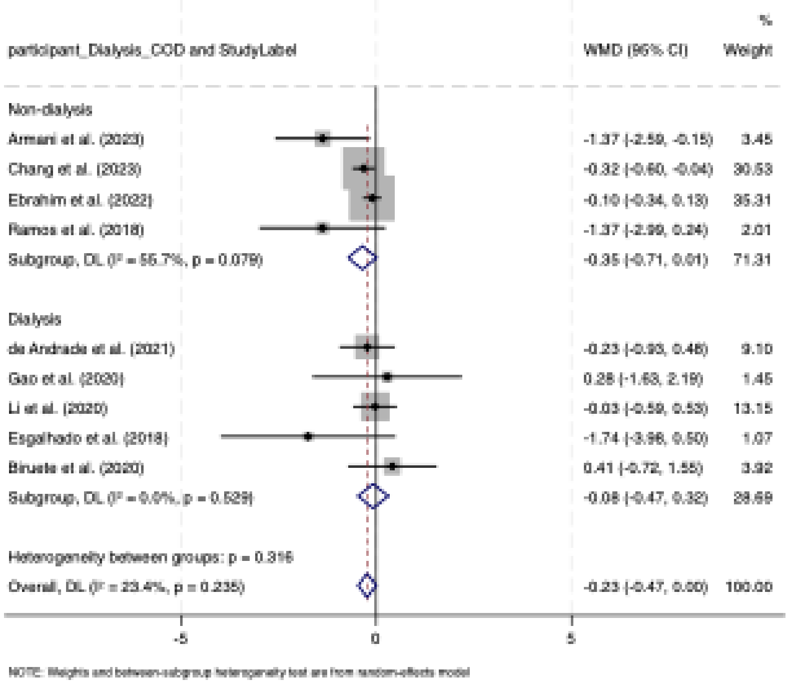
%
participant_Dialysis_COD and StudyLabel
WMD (95% CI)
Weight
Non-dialysis
Armani et al. (2023)
-1.37 (-2.59, -0.15)
3.45
Chang et al. (2023)
-0.32 (-0.60, -0.04)
30.53
Ebrahim et al. (2022)
-0.10 (-0.34, 0.13)
35.31
Ramos et al. (2018)
-1.37 (-2.99, 0.24)
2.01
Subgroup, DL (I² = 55.7%, p = 0.079)
-0.35 (-0.71, 0.01)
71.31
Dialysis
de Andrade et al. (2021)
-0.23 (-0.93, 0.46)
9.10
Gao et al. (2020)
0.28 (-1.63, 2.19)
1.45
Li et al. (2020)
-0.03 (-0.59, 0.53)
13.15
Esgalhado et al. (2018)
-1.74 (-3.98, 0.50)
1.07
Biruete et al. (2020)
0.41 (-0.72, 1.55)
3.92
Subgroup, DL (I² = 0.0%, p = 0.529)
-0.08 (-0.47, 0.32)
28.69
Heterogeneity between groups: p = 0.316
Overall, DL (I² = 23.4%, p = 0.235)
-0.23 (-0.47, 0.00)
100.00
-5
0
5
NOTE: Weights and between-subgroup heterogeneity test are from random-effects model

i)

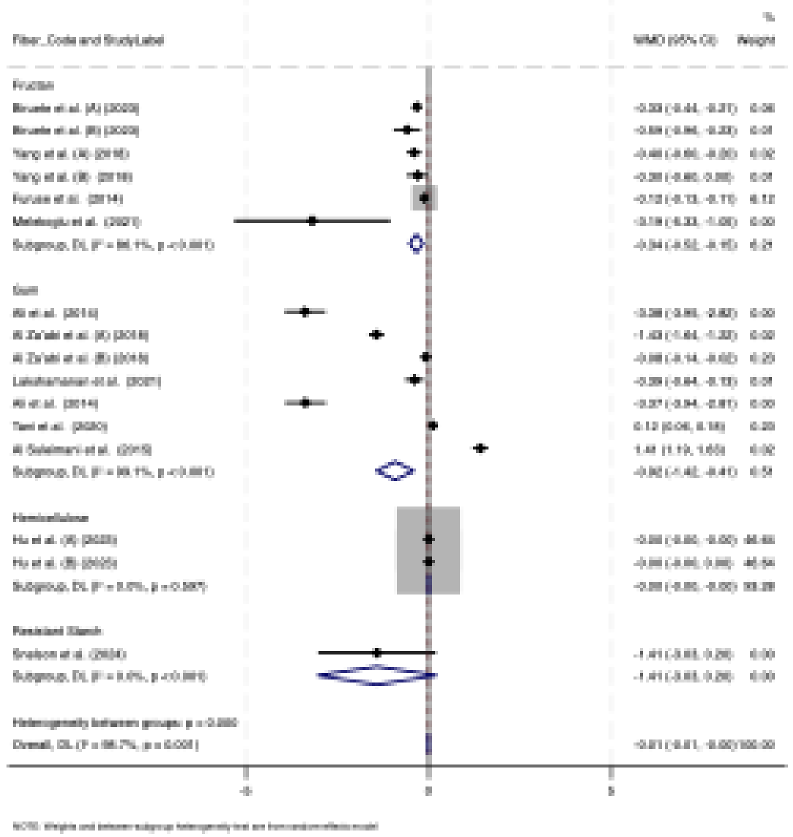
%
WMD (95% CI)
Weight
Fructan
Gum
Hemicellulose
Resistant Starch
-5
0
5

j)

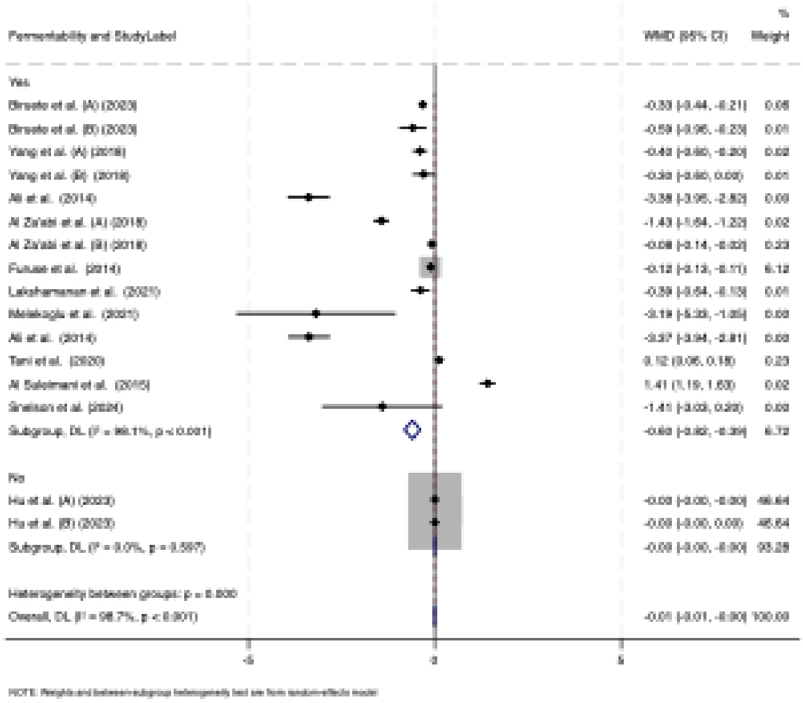
Fermentability and StudyLabel
%
WMD (95% CI)
Weight
Yes
No
-5
0
5

k)

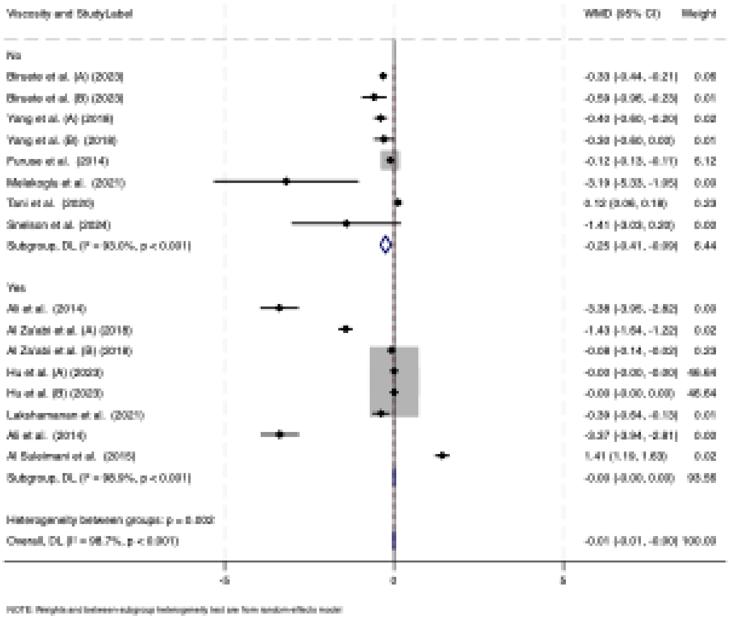

l)

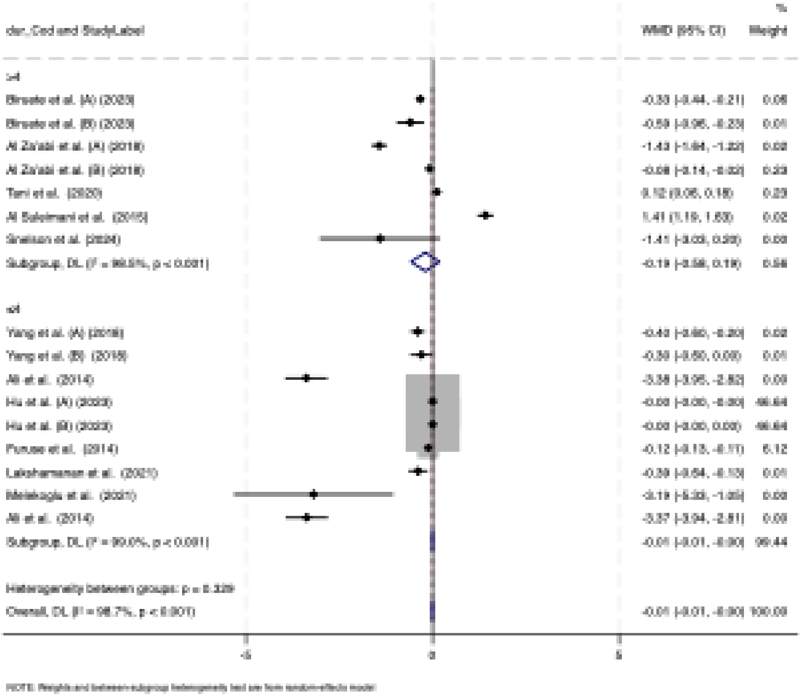

m)

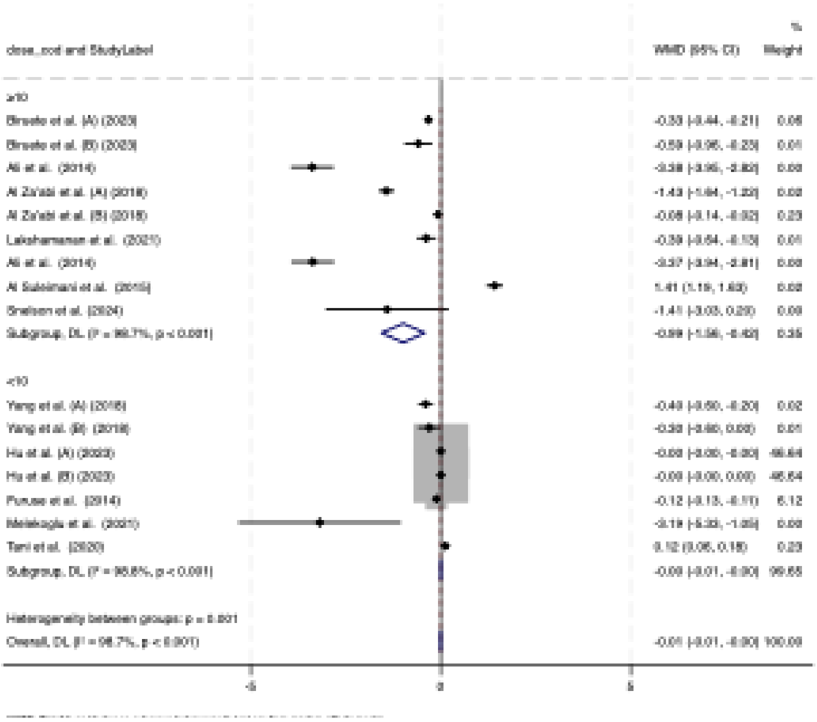
%
Weight
Ali et al. (2014)
Al Za'abi et al. (A) (2018)
Al Za'abi et al. (B) (2018)
Ali et al. (2014)
Yang et al. (A) (2018)
Yang et al. (B) (2018)
Hu et al. (A) (2023)
Hu et al. (B) (2023)
Furuse et al. (2014)
Tani et al. (2020)
-5
0
5

n)

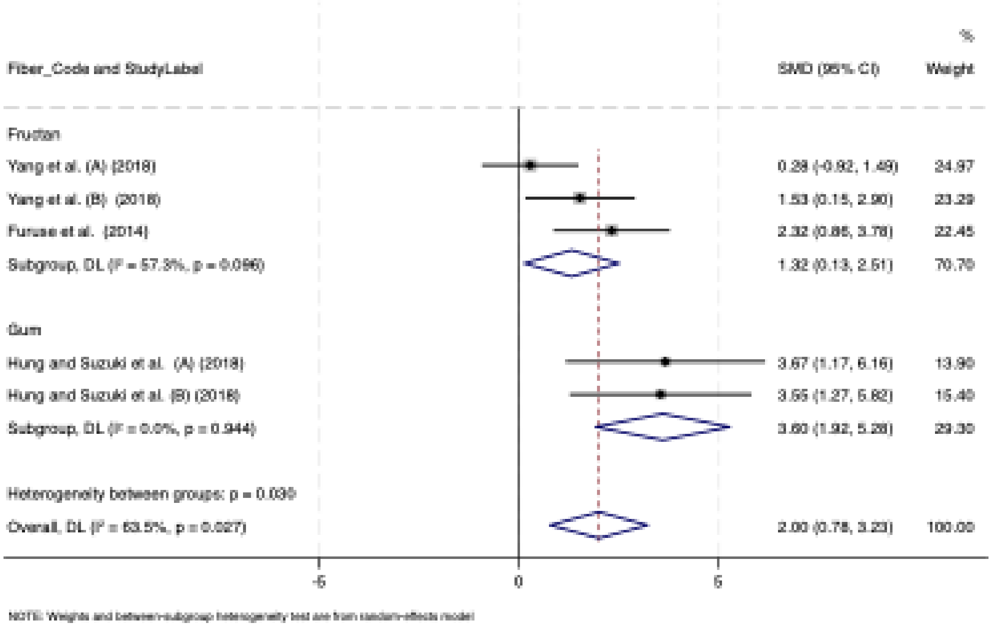
%
Fiber_Code and StudyLabel
SMD (95% CI)
Weight
Fructan
Yang et al. (A) (2018) 0.28 (-0.92, 1.49) 24.97
Yang et al. (B) (2018) 1.53 (0.15, 2.90) 23.29
Furuse et al. (2014) 2.32 (0.86, 3.78) 22.45
Subgroup, DL (I² = 57.3%, p = 0.096) 1.32 (0.13, 2.51) 70.70
Gum
Hung and Suzuki et al. (A) (2018) 3.67 (1.17, 6.16) 13.90
Hung and Suzuki et al. (B) (2018) 3.55 (1.27, 5.82) 15.40
Subgroup, DL (I² = 0.0%, p = 0.944) 3.60 (1.92, 5.28) 29.30
Heterogeneity between groups: p = 0.030
Overall, DL (I² = 63.5%, p = 0.027) 2.00 (0.78, 3.23) 100.00
-5
0
5
NOTE: Weights and between-subgroup heterogeneity test are from random-effects model

**Figure S1a-h.** Forest plot detailing the random-effects model of weighted mean difference (WMD) with 95% confidence intervals (CIs) for the subgroup analysis of the effect of dietary fiber supplementation on indoxyl sulfate (mg/dL) (a-d) and total serum/plasma p-cresyl sulfate (pCS) (e-h) in adults with chronic kidney disease (CKD) **by**: (**a**) dietary fiber type, (**b**) trial duration (week), (**c**) intervention dose (g/d), (**d**) dialysis status (**e**) dietary fiber type, (**f**) trial duration (week), (**g**) intervention dose (g/d), (**h**) dialysis status.

**Figure S1 i-m.** Forest plot detailing the random-effects model of standardized mean difference (SMD) with 95% confidence intervals (CIs) for the subgroup analysis of the effect of dietary fiber supplementation on Indoxyl sulfate (mg/dL) (i-m) in animal models with Chronic Kidney Disease (CKD) **by**: (**i**) dietary fiber type, (**j**) fermentability, (**k**) Viscosity, (**l**) Intervention dose, (**m**) intervention duration.

**Figure S1 n.** Forest plot detailing the random-effects model of standardized mean difference (SMD) with 95% confidence intervals (CIs) for the subgroup analysis of the effect of dietary fiber supplementation on cecal acetate (mg/dL) in animal models with chronic kidney disease (CKD) **by**: (**n**) dietary fiber type

a)

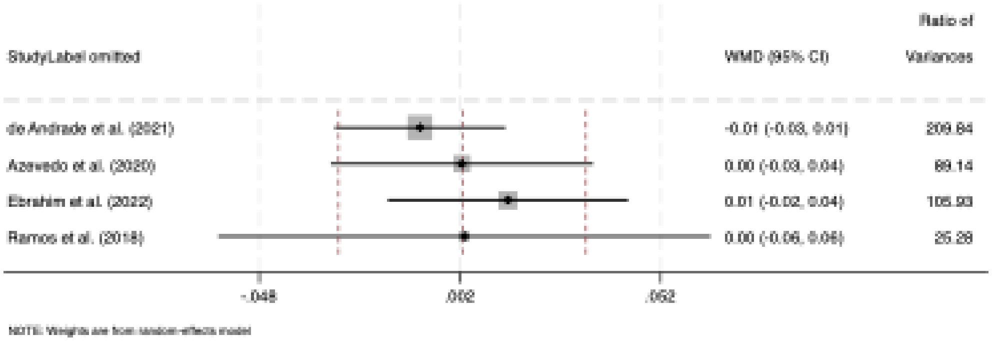


b)

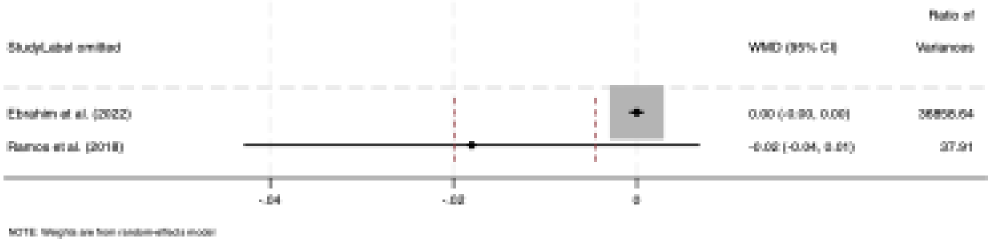


c)

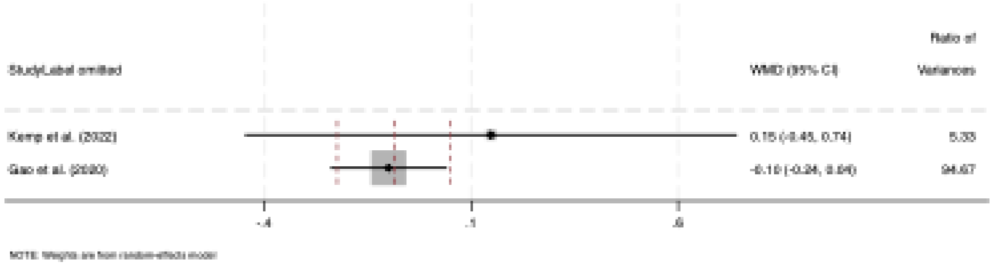

Ratio of
StudyLabel omitted
WMD (95% CI)
Variances
Kemp et al. (2022)
Gao et al. (2020)
-.4
.1
.6
NOTE: Weights are from random-effects model


d)

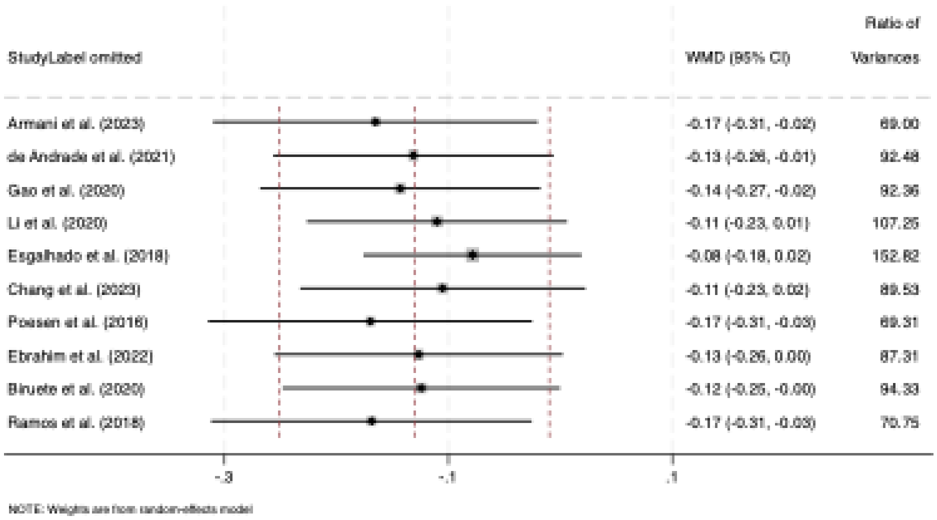

Ratio of
StudyLabel omitted
WMD (95% CI)
Variances
Armani et al. (2023)
-0.17 (-0.31, -0.02)
69.00
de Andrade et al. (2021)
-0.13 (-0.26, -0.01)
92.48
Gao et al. (2020)
-0.14 (-0.27, -0.02)
92.36
Li et al. (2020)
-0.11 (-0.23, 0.01)
107.25
Esgalhado et al. (2018)
-0.08 (-0.18, 0.02)
152.82
Chang et al. (2023)
-0.11 (-0.23, 0.02)
89.53
Poesen et al. (2016)
-0.17 (-0.31, -0.03)
69.31
Ebrahim et al. (2022)
-0.13 (-0.26, 0.00)
87.31
Biruete et al. (2020)
-0.12 (-0.25, -0.00)
94.33
Ramos et al. (2018)
-0.17 (-0.31, -0.03)
70.75
-.3
-.1
.1
NOTE: Weights are from random-effects model


e)

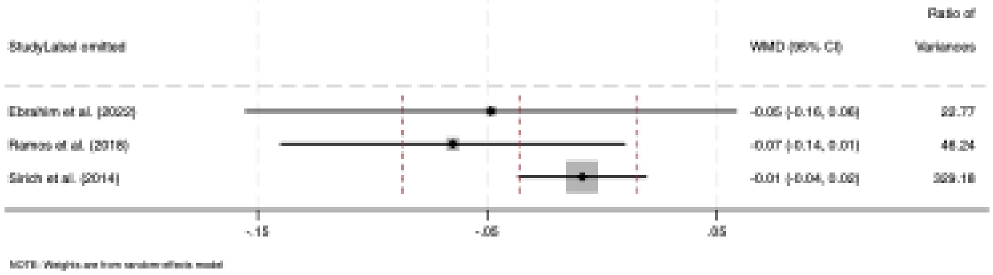

Ratio of
StudyLabel omitted
WMD (95% CI)
Variances
Ebrahim et al. (2022)
Ramos et al. (2018)
Sirich et al. (2014)
-.15
-.05
.05
NOTE: Weights are from random-effects model


f)

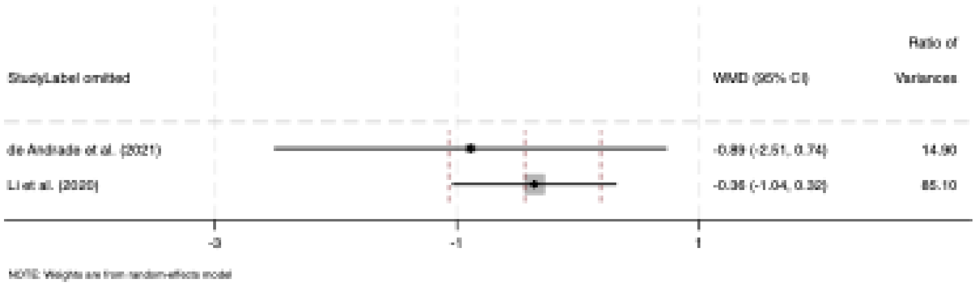

StudyLabel omitted
WMD (95% CI)
Ratio of Variances
de Andrade et al. (2021)
-0.89 (-2.51, 0.74)
14.90
Li et al. (2020)
-0.36 (-1.04, 0.32)
85.10
-3
-1
1
NOTE: Weights are from random-effects model


g)

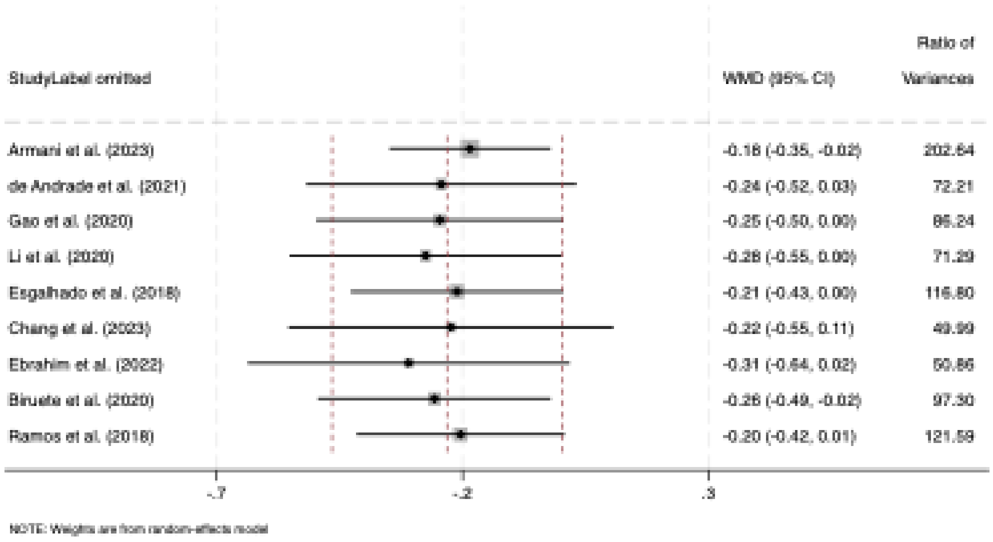

StudyLabel omitted
WMD (95% CI)
Ratio of Variances
Armani et al. (2023)
-0.18 (-0.35, -0.02)
202.64
de Andrade et al. (2021)
-0.24 (-0.52, 0.03)
72.21
Gao et al. (2020)
-0.25 (-0.50, 0.00)
86.24
Li et al. (2020)
-0.28 (-0.55, 0.00)
71.29
Esgalhado et al. (2018)
-0.21 (-0.43, 0.00)
116.80
Chang et al. (2023)
-0.22 (-0.55, 0.11)
49.99
Ebrahim et al. (2022)
-0.31 (-0.64, 0.02)
50.86
Biruete et al. (2020)
-0.26 (-0.49, -0.02)
97.30
Ramos et al. (2018)
-0.20 (-0.42, 0.01)
121.59
-.7
-.2
.3
NOTE: Weights are from random-effects model


h)

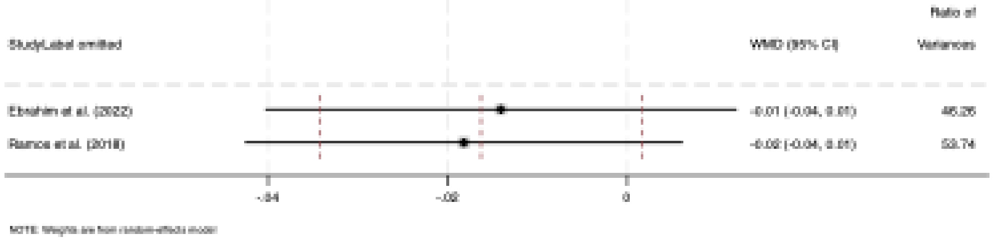

StudyLabel omitted
WMD (95% CI)
Ratio of Variances
Ebrahim et al. (2022)
-0.01 (-0.04, 0.01)
46.26
Ramos et al. (2018)
-0.02 (-0.04, 0.01)
53.74
-.04
-.02
0
NOTE: Weights are from random-effects model


i)

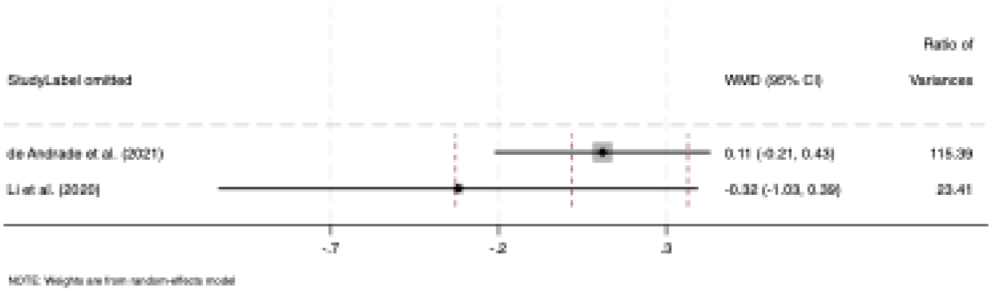
StudyLabel omitted
WMD (95% CI)
Ratio of Variances
de Andrade et al. (2021)
0.11 (-0.21, 0.43)
115.39
Li et al. (2020)
-0.32 (-1.03, 0.39)
23.41
-.7
-.2
.3
NOTE: Weights are from random-effects model

j)

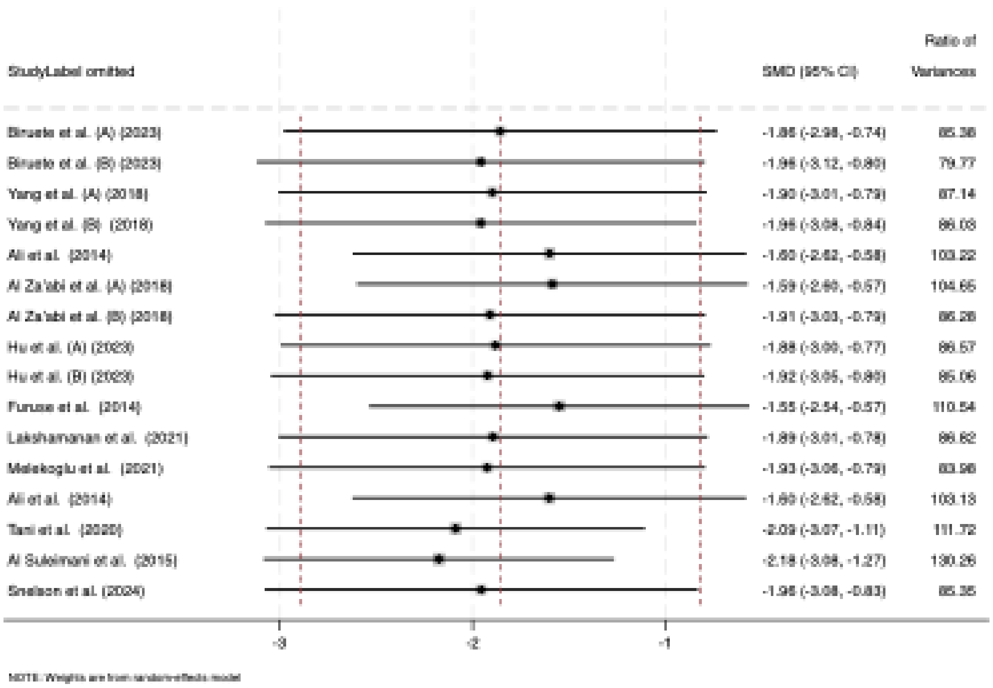
StudyLabel omitted
SMD (95% CI)
Ratio of Variances
Biruete et al. (A) (2023)
-1.86 (-2.98, -0.74)
85.36
Biruete et al. (B) (2023)
-1.96 (-3.12, -0.80)
79.77
Yang et al. (A) (2018)
-1.90 (-3.01, -0.79)
87.14
Yang et al. (B) (2018)
-1.96 (-3.08, -0.84)
86.03
Ali et al. (2014)
-1.60 (-2.62, -0.58)
103.22
Al Za'abi et al. (A) (2018)
-1.59 (-2.60, -0.57)
104.65
Al Za'abi et al. (B) (2018)
-1.91 (-3.03, -0.79)
86.28
Hu et al. (A) (2023)
-1.88 (-3.00, -0.77)
86.57
Hu et al. (B) (2023)
-1.92 (-3.05, -0.80)
85.06
Furuse et al. (2014)
-1.55 (-2.54, -0.57)
110.54
Lakshamanan et al. (2021)
-1.89 (-3.01, -0.78)
86.62
Melekoglu et al. (2021)
-1.93 (-3.06, -0.79)
83.96
Ali et al. (2014)
-1.60 (-2.62, -0.58)
103.13
Tani et al. (2020)
-2.09 (-3.07, -1.11)
111.72
Al Suleimani et al. (2015)
-2.18 (-3.08, -1.27)
130.26
Snelson et al. (2024)
-1.96 (-3.08, -0.83)
85.35
-3
-2
-1
NOTE: Weights are from random-effects model

k)

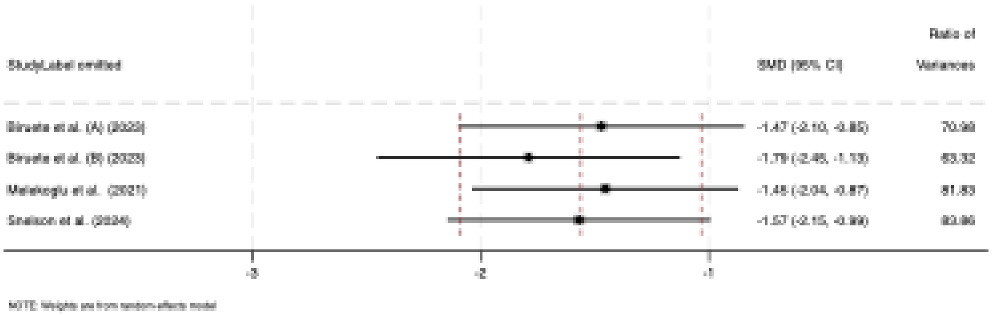
StudyLabel omitted
SMD (95% CI)
Ratio of
Variances

l)

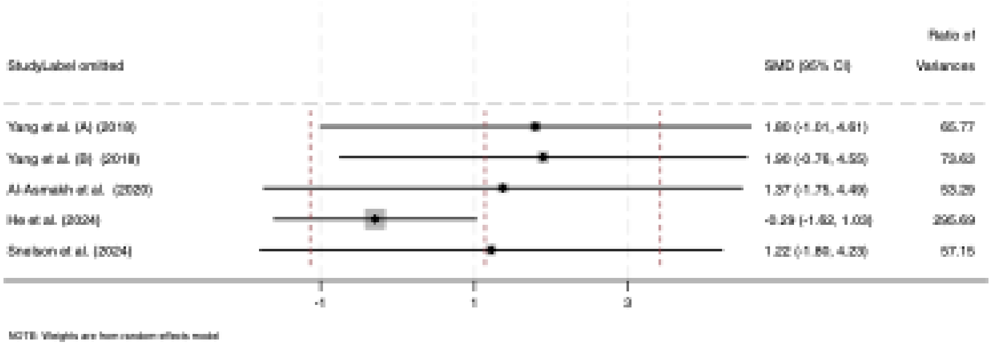
StudyLabel omitted
SMD (95% CI)
Ratio of
Variances

m)

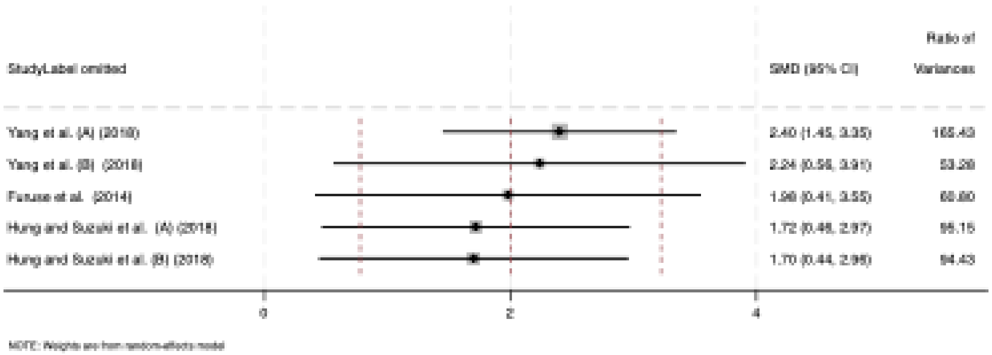
StudyLabel omitted
SMD (95% CI)
Ratio of
Variances

n)

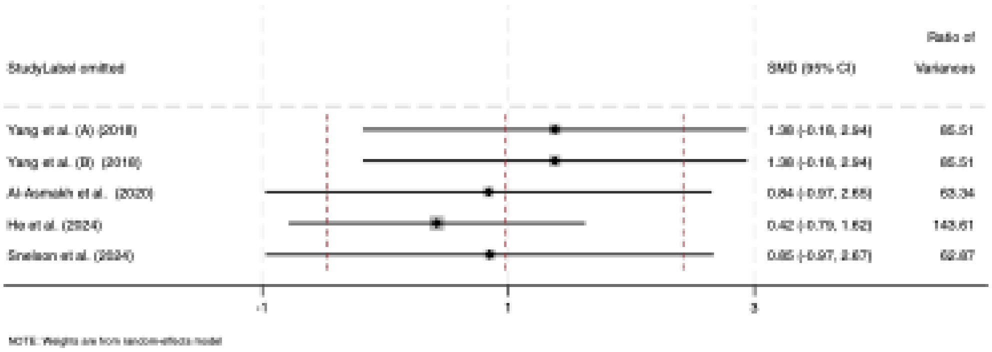
Ratio of
Variances
StudyLabel omitted
SMD (95% CI)
Yang et al. (A) (2018)
Yang et al. (B) (2018)
Al-Asmakh et al. (2020)
He et al. (2024)
Snelson et al. (2024)
-1
1
3
NOTE: Weights are from random-effects model

0)

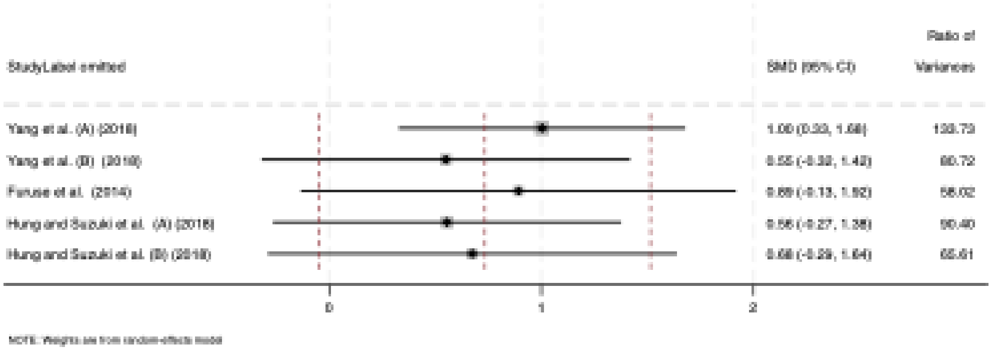
Ratio of
Variances
StudyLabel omitted
SMD (95% CI)
Yang et al. (A) (2018)
Yang et al. (B) (2018)
Furuse et al. (2014)
Hung and Suzuki et al. (A) (2018)
Hung and Suzuki et al. (B) (2018)
0
1
2
NOTE: Weights are from random-effects model

p)

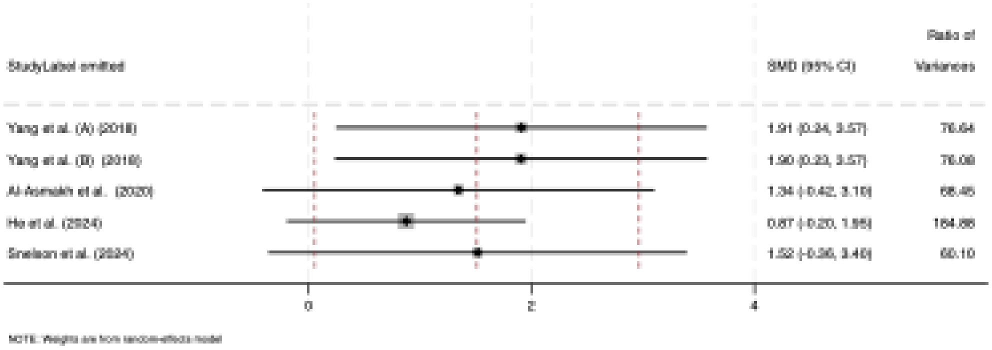
Ratio of
Variances
StudyLabel omitted
SMD (95% CI)
Yang et al. (A) (2018)
Yang et al. (B) (2018)
Al-Asmakh et al. (2020)
He et al. (2024)
Snelson et al. (2024)
0
2
4
NOTE: Weights are from random-effects model

q)

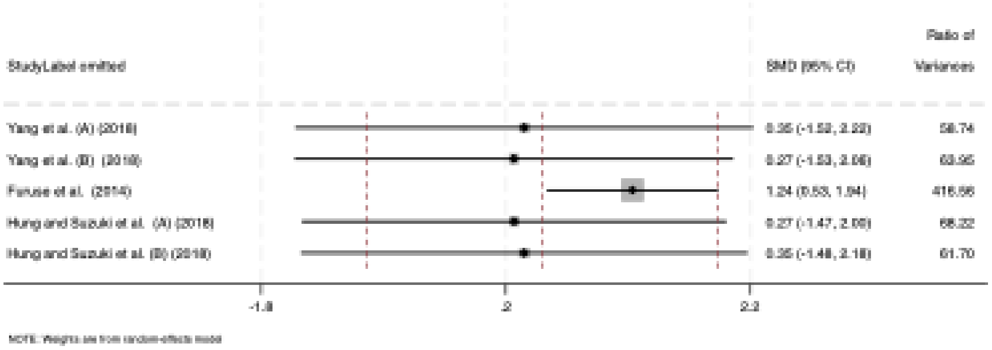


**Figure S2a-i.** Forest plot detailing the random-effects model of weighted mean difference (WMD) with 95% confidence intervals (CIs) for the sensitivity analysis of the effect of dietary fiber supplementation on gut microbiota metabolites (mg/dL) in adults with Chronic Kidney Disease (CKD) **(a-i),** including: (**a**) Total plasma/serum Indole-3-acetic acid (IAA), (**b**) Free plasma/serum Indole-3-acetic acid (IAA), (**c**) Plasma/serum trimethylamine-N-oxide (TMAO), (**d**) Total serum/plasma indoxyl sulfate (IS), (**e**) Free serum/plasma indoxyl sulphate (IS), (**f**) Urinary indoxyl sulfate (IS), (**g**) Total serum/plasma p-cresyl sulfate (pCS), (**h**) Free serum/plasma p-cresyl sulfate (pCS), (**i**) Urinary serum/plasma p-cresyl sulfate (pCS),

**Figure S2j-q.** Forest plot detailing the random-effects model of standardized mean difference (SMD) with 95% confidence intervals (CIs) for the sensitivity analysis of the effect of dietary fiber supplementation on gut microbiota metabolites (mg/dL) in animal models with Chronic Kidney Disease (CKD, (**j**) Total serum/plasma indoxyl sulfate (IS), (**k**) Total serum/plasma p-cresyl sulfate (pCS), (**l**) serum/plasma acetate, (**m**) cecal acetate, (**n**) serum/plasma butyrate, (**o**) cecal butyrate, (**p**) serum/plasma propionate, **(q**) cecal propionate.

a)

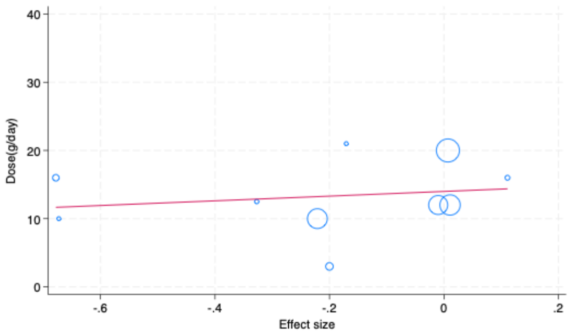


b)

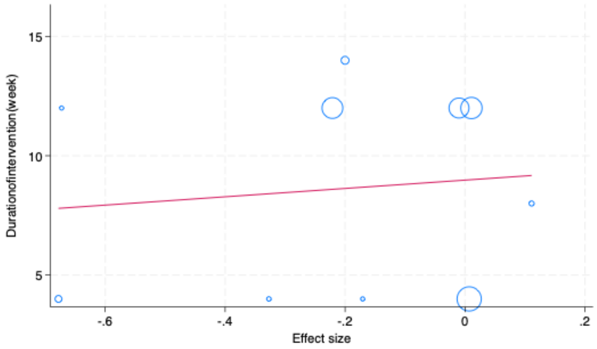


c)

d)

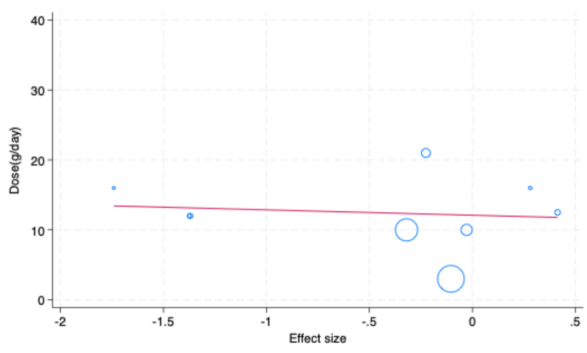


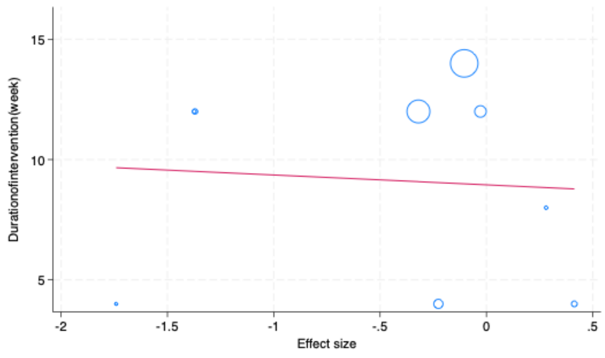


e)

f)

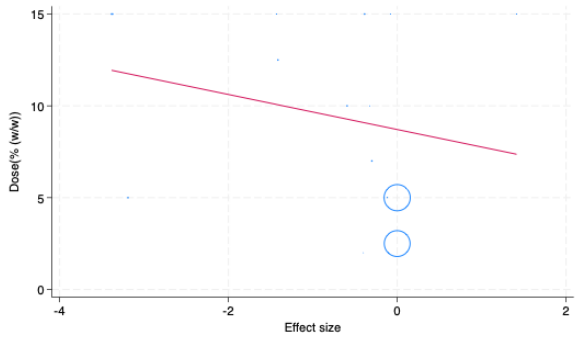


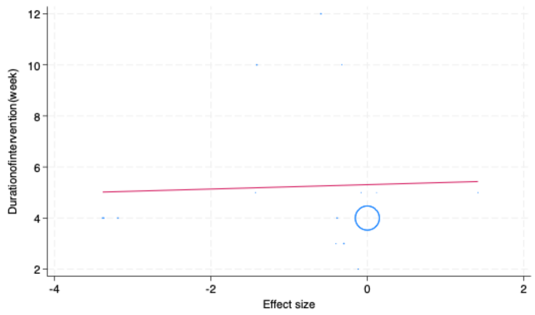


**Figure S3a-d.** Meta regression graph of linear effect of dietary fiber supplementation on Indoxyl sulfate (mg/dL) (a) Dose (g/day) b) Duration (week), and on serum/plasma p-cresyl sulfate (pCS) (c) Dose (g/day) (d) Duration (week) in adults with Chronic Kidney Disease (CKD). **Figure S3e-h.** Meta regression graph of linear effect of dietary fiber supplementation on Indoxyl sulfate (mg/dL) (a) Dose (g/day) b) Duration (week), and total serum/plasma p-cresyl sulfate (pCS) (mg/dL) (c) Dose (%) (d) Duration (week) in animal models with chronic kidney disease (CKD).
